# Expanding reproductive genetic screening through the inclusion of perinatal treatability

**DOI:** 10.64898/2026.08.24.26361139

**Authors:** Tse Yen Tan, Sarah Haas, Xiaoke Gao, Judy Li, Sarah Araji, Alex Liu, Courtney Wimberly, Nina Gold, Stefan Rentas, Michael Duyzend, Kyle M. Walsh, Jennifer L. Cohen

## Abstract

Various professional organizations recommend screening prospective parents for autosomal recessive (AR) and X-linked (XL) conditions, which is reflected in commercial screening panels. There is merit to developing a distinct reproductive gene-list and analytic framework inclusive of genes based on available perinatal intervention, defined as possible prenatal intervention (including investigational) for the fetus or necessary early initiation of approved postnatal treatments. We evaluated a reproductive genetic screening framework that incorporates perinatal actionability across AR, XL, and selected autosomal dominant (AD) genes. Using a curated list of genetic conditions with perinatal intervention, we evaluated five subset gene lists to determine the individual-level number-needed-to-screen (NNS) to identify one individual with at least one qualifying heterozygous variant, defined as a heterozygous pathogenic or likely pathogenic (P/LP) variant in a gene on the specified list. To conduct NNS analyses, we sourced carrier frequency and allele frequency data for each gene and their respective ClinVar-curated high-confidence (≥2 star) P/LP variants, from two population databases—gnomAD v4.1 and All of Us (AoU) v8. The analyses produced an individual-level NNS of 3.20 (CI: 3.193, 3.212) using gnomAD and 3.62 (CI: 3.606, 3.640) using AoU. These estimates do not represent couple-level reproductive risk, affected-pregnancy yield, clinical diagnostic yield, or validation of a clinical screening test. These findings support further evaluation of a perinatal-actionability framework, with clinical value dependent on which genes drive yield, and whether the relevant gene, variant, mechanism, and phenotype combinations are actionable in a reproductive or perinatal context for both the pregnant woman and her future offspring.

## Introduction

Carrier screening is a form of reproductive or preconception clinical genetic testing traditionally performed on ostensibly asymptomatic individuals to identify whether they possess a pathogenic or likely pathogenic (P/LP) variant in a gene associated with an autosomal recessive (AR) or X-linked (XL) genetic disease, with the key objective of informing procreative risk. The organizing principle of this traditional methodology does not account for whether a genetic condition has possible prenatal (including investigational) or imperative early neonatal intervention opportunities in the first week of life, referred to for the purposes of this study, as genes with “perinatal treatability” that have been described in our group’s prior work.(1) Furthermore, autosomal dominant (AD) conditions remain underrepresented in carrier screening, some of which may have maternal pregnancy health implications. Our study attempts to address these limitations by proposing a broader reproductive genetic screening framework centered on perinatal actionability, extending beyond the traditional AR and XL inheritance patterns used for carrier screening panels.

The American College of Obstetricians and Gynecologists (ACOG) recommends that information about the option for carrier screening be shared with every pregnant patient.(2) Along these lines, the American College of Medical Genetics and Genomics (ACMG) has produced a list of genes that should be offered to all individuals seeking carrier screening.(3) Ideally, screening occurs prior to conception. In the event that an individual is identified as a carrier—that is, heterozygous for a disease-causing, P/LP variant in an AR gene or a female carrying a P/LP variant in an XL gene—this screening process allows time for couples to understand their reproductive risks and consider a full range of reproductive options before they conceive. While it is recommended that all patients be informed of the option for carrier screening, participation in screening is voluntary, and individuals retain the autonomy to decline any or all components of testing. The carrier’s reproductive partner can also undergo sequential targeted screening of the affected gene if the partner did not initially opt into concurrent broad screening. If both partners are found to be carriers for the same AR condition, or if the mother is a carrier for an XL genetic condition, post-test genetic counseling is critical to inform the couple of their risk to transmit the corresponding disease-causing alleles to a current or future fetus.

Early identification of genetic conditions through reproductive genetic screening (including traditional carrier screening) and the subsequent opportunity for prenatal screening and diagnostic testing may be beneficial in a variety of instances. For a subset of genetic diseases, prenatal interventions are emerging that may be safe and plausibly efficacious for affected fetuses, such as investigational *in utero* enzyme replacement therapy for lysosomal storage diseases(4) or investigational intraamniotic protein replacement for XL hypohidrotic ectodermal dysplasia (*EDA* [MIM: 305100]).(5) Even when prenatal intervention is not available, early diagnosis can enable timely postnatal treatments, guide labor and delivery clinical management, or empower families to make informed reproductive decisions.(1) Delaying diagnosis until after birth may result in missed opportunities for treatment or preparation, underscoring the importance of expanding access to and awareness of reproductive genetic screening and follow-up prenatal screening and diagnostic testing.

Despite overall advances in genetic testing, prenatal diagnostic testing is often limited to instances with a known family history of disease or the presence of fetal anomalies on prenatal imaging, which can result in missed or delayed diagnoses. Expanding reproductive genetic screening and developing strategies for risk stratification could help identify at-risk pregnancies earlier, even in the absence of these traditional indicators. Earlier identification would not only enable access to emerging *in utero* therapies but also allow families to make informed reproductive decisions and prepare for the birth of a child with medical complexities after diagnostic testing. Research efforts are increasingly focused on accelerating diagnosis and improving prenatal risk assessment to optimize outcomes for affected individuals. There is growing recognition that phenotype-driven diagnostic approaches, especially in prenatal and neonatal settings, may miss affected individuals who do not present with early symptoms.(6) As genomic technologies advance, there is a need for more proactive, data-driven strategies that can improve early diagnosis and guide clinical decision-making and intervention.

There is wide variability in carrier screening panel composition and inconsistency in how screening is offered, with ongoing debate about whether screening goals should extend beyond AR/XL reproductive risk assessment or be more targeted to perinatal treatable conditions. In answer to this, our work reimagines the potential purpose and function of screening by not just expanding the number of genes screened for, but specifically by including genes because of their potential treatability and maternal health implications. In general, ACOG recommends genetic carrier screening for four disease categories: spinal muscular atrophy [MIM: 253300], *SMN1;* cystic fibrosis [MIM: 219700], *CFTR*; hemoglobinopathies (e.g. beta-thalassemia [MIM: 613985] *HBB*; sickle cell [MIM: 603903] *HBB;* alpha-thalassemia [MIM: 604131]*, HBA1;* Hemoglobin H disease [MIM: 613978], *HBA2);* and, in the presence of specific risk factors, Fragile X premutation testing ([MIM: 300624], *FMR1*).(2) ACOG also recommends that women of Ashkenazi Jewish, French-Canadian, or Cajun descent who are considering pregnancy be screened for Tay-Sachs disease (*HEXA* [MIM: 272800]).(2) Beyond the aforementioned four disease categories, ACOG states that expanded carrier screening is acceptable but does not specify which genes should be included. On the other hand, the ACMG has developed a tiered approach to screening, and the recommended tier 3 (T3) list consists of 113 AR and XL conditions determined based on carrier frequency thresholds and clinical utility.(7, 8) The lack of consensus between professional society guidelines contributes to the substantial variation in commercial panel composition (Table 1, Supplemental Table 1).(9) This variability not only reflects differences in gene selection, but also the unresolved question of whether reproductive genetic screening should remain focused on AR and XL risk or evolve into a broader framework that includes selected AD conditions with relevance to treatability and maternal health.

**Table 1.**
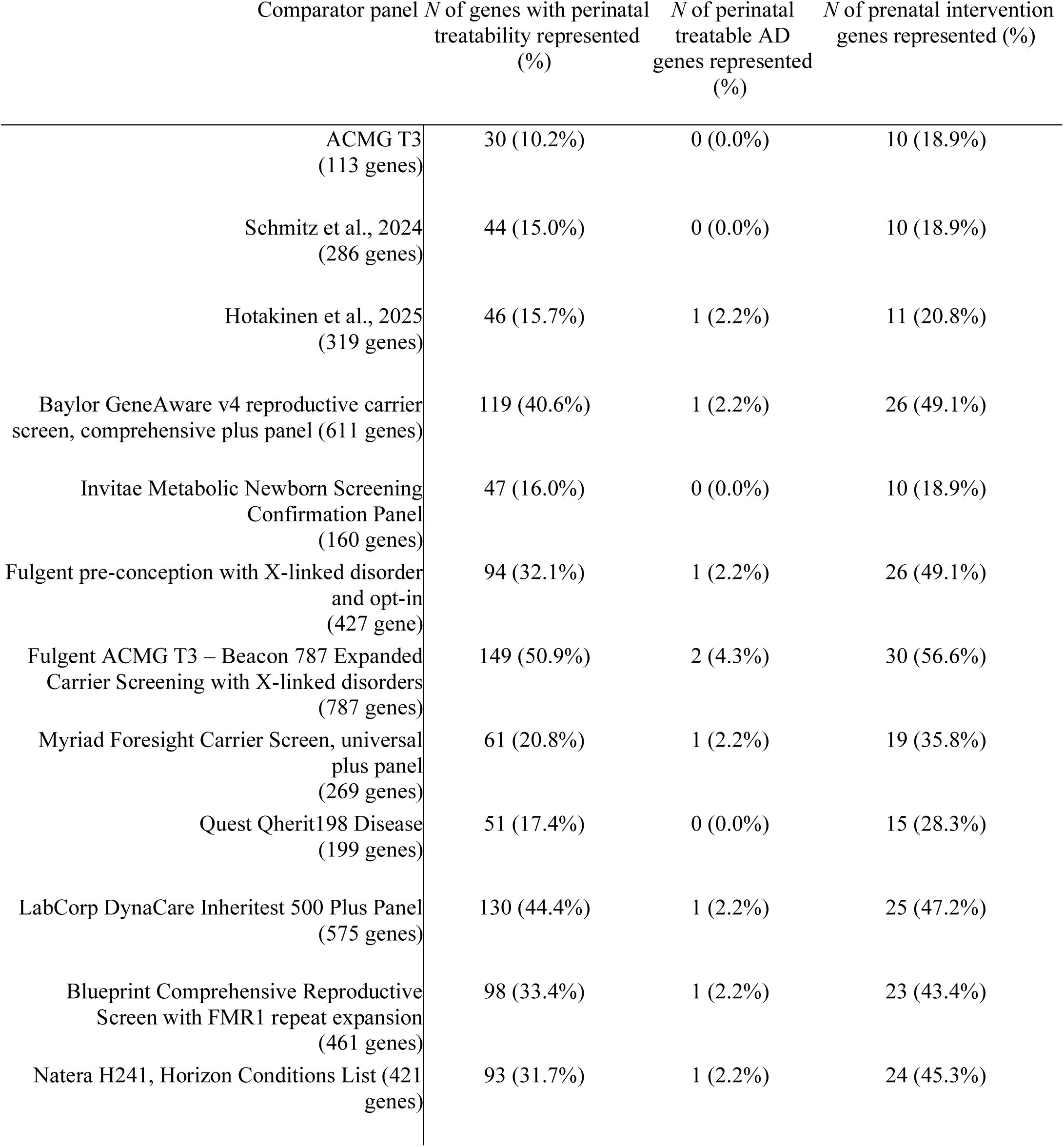
Summary table of the number and percentage of the following gene lists represented in each comparator panel: a 293 perinatal treatability gene list, subset list of 46 autosomal dominant genes, and a subset list of 52 prenatal intervention genes.

| Comparator panel | *N* of genes with perinatal treatability represented (%) | *N* of perinatal treatable AD genes represented (%) | *N* of prenatal intervention genes represented (%) |
| --- | --- | --- | --- |
| ACMG T3 (113 genes) | 30 (10.2%) | 0 (0.0%) | 10 (18.9%) |
| Schmitz et al., 2024 (286 genes) | 44 (15.0%) | 0 (0.0%) | 10 (18.9%) |
| Hotakinen et al., 2025 (319 genes) | 46 (15.7%) | 1 (2.2%) | 11 (20.8%) |
| Baylor GeneAware v4 reproductive carrier screen, comprehensive plus panel (611 genes) | 119 (40.6%) | 1 (2.2%) | 26 (49.1%) |
| Invitae Metabolic Newborn Screening Confirmation Panel (160 genes) | 47 (16.0%) | 0 (0.0%) | 10 (18.9%) |
| Fulgent pre-conception with X-linked disorder and opt-in (427 gene) | 94 (32.1%) | 1 (2.2%) | 26 (49.1%) |
| Fulgent ACMG T3 – Beacon 787 Expanded Carrier Screening with X-linked disorders (787 genes) | 149 (50.9%) | 2 (4.3%) | 30 (56.6%) |
| Myriad Foresight Carrier Screen, universal plus panel (269 genes) | 61 (20.8%) | 1 (2.2%) | 19 (35.8%) |
| Quest Qherit198 Disease (199 genes) | 51 (17.4%) | 0 (0.0%) | 15 (28.3%) |
| LabCorp DynaCare Inheritest 500 Plus Panel (575 genes) | 130 (44.4%) | 1 (2.2%) | 25 (47.2%) |
| Blueprint Comprehensive Reproductive Screen with FMR1 repeat expansion (461 genes) | 98 (33.4%) | 1 (2.2%) | 23 (43.4%) |
| Natera H241, Horizon Conditions List (421 genes) | 93 (31.7%) | 1 (2.2%) | 24 (45.3%) |

Although the ACMG T3 list provides an important framework for expanded carrier screening, its application depends on population-level estimates of carrier frequency and clinical utility that can differ in completeness across ancestral groups and disease areas.(3) Moreover, clinical utility in the carrier-screening context does not necessarily align with early, perinatal treatability. Many genes associated with perinatal treatability(1) are absent from both commercial carrier screening panels and the ACMG T3 gene list (Supplemental Table 1). NIH’s All of Us (AoU) program has attempted to increase ancestral diversity among its dataset, though the total sample size of individuals is still limited compared to gnomAD. Our study aims to refine the inclusion criteria for reproductive genetic screening panels using heterozygote frequencies derived from an evaluation of both gnomAD v4.1 and AoU, among a list of perinatal treatable conditions with a variety of inheritance patterns. We investigate, for instance, the potential utility of including AD genes with perinatal treatability, if they are incompletely penetrant and variably expressive, especially if these conditions have potential for maternal pregnancy health implications. If a gene is known to have incomplete penetrance and intrafamilial variability, it is plausible that a procreative-aged adult could be harboring a P/LP variant in an AD condition and have a fetus/child who inherits that variant with subsequent severe, early-onset disease in need of immediate management.

Our study’s departure from traditional carrier screening recommendations is in line with more recent research and interest in incorporating genes associated with prenatal lethality (miscarriage), fertility, and obstetric management into screening panels. For instance, a study by Aminbeidokhti and Qu (2023) identified 138 genes in which heterozygous variants are present at a carrier frequency of at least 0.5% in the general population.(10) These genes are hypothesized to be associated with embryonic or fetal lethality when biallelic P/LP variants are inherited. The authors estimate that screening for these genes could identify between 4.6% and 39.8% of couples at risk of miscarriage, depending on the population. This suggests that a substantial proportion of unexplained pregnancy losses may be attributable to recessive lethal variants.(10) Another recent study noted that 1.7% of expanded carrier screening results impacted fertility care and 3.9% of results impacted the personal health of the patient undergoing the carrier screening.(11) A study by Rosenfeld and colleagues (2026) further highlighted several genetic conditions with strong clinical implications in the carrier state that could also affect pregnancy, demonstrating how carrier screens may not only provide reproductive risk assessment but also provide invaluable insights into the carrier’s health status and influence obstetric management.(12)

The current study’s primary outcome is individual-level heterozygote yield. We aimed to define and curate a perinatal-actionability gene list across AR, XL, and selected AD genes, and to estimate the number of individuals needed to screen (NNS) to identify one individual with a qualifying heterozygous P/LP variant in one of the genes screened. These analyses approximate individual-level heterozygote yield and do not validate a clinical screening test, estimate couple-level reproductive risk, or estimate affected-pregnancy yield. The NNS analysis was designed to evaluate analytic yield for curated perinatal-actionability gene lists rather than to establish clinical utility for implementation. First, a “perinatal treatable findings” gene list, based on our previous publication^1^ was evaluated and further curated by the study team for relevance in a reproductive genetic screening context, and a finalized version of this list, consisting of AD, XL, and AR genes, was created. Subsets of this gene list were evaluated for NNS for combined ancestries and across separate ancestral subgroups as follows: 1) Full 293-gene list; 2) 46 AD genes; 3) all XL + AR genes from the list; 4) 289-gene list, excluding 4 AD genes; and 5) 42 AD genes, excluding 4 genes due to consistent high penetrance and early-onset phenotype.

## METHODS

### Gene Panel Selection and Composition

To determine which genes to include in our NNS analysis, we built upon a previously published “treatable fetal findings list” developed by our group.(1) This list was originally designed to guide the reporting of incidental findings from fetal genomic sequencing (GS), particularly in cases where early diagnosis could inform prenatal or immediate postnatal (first week of life) clinical management. The list was developed through an integrative review of genetic conditions with potential for prenatal or early-life intervention, prioritizing those with the greatest intervention and treatment opportunities. In total, the list included 296 genes.

For the purposes of creating subset gene lists in this study, gene inheritance patterns were classified based on the relevant treatable phenotypes noted in the aforementioned publication.^1^ For example, *NR5A1* (MIM: 184757) has a severe, early-onset form of disease when mutations are inherited in an AR pattern (e.g. homozygous for P/LP variant). *PDX1* (MIM: 600733) has a pancreatic agenesis phenotype associated with AR inheritance(13, 14), but the dominant form of monogenic diabetes (MIM: 606392)(15) is what was used for inheritance classification in this study and the previously published work.(1) Lastly, *GUCY2C* (MIM: 601330) has two separate phenotypes and the AD phenotype of congenital sodium diarrhea (MIM: 614616) was the inheritance pattern used for subset gene list classification. If a gene’s specific phenotype of interest was noted to have both AD and AR inheritance, for the purposes of downstream NNS subset gene panel analyses, these genes were classified as AR. In total, 293 genes (Supplemental Table 2) were included in the final analysis, classified by inheritance pattern: AR (n=222), AD (n=46), and XL (n=25). Three genes from the original gene list(1) were excluded from reproductive genetic screening analysis: *SMN1* (MIM: 600354), which requires specialized copy number variant (CNV) detection not available in standard single nucleotide variant (SNV) pipelines due to 99.9% sequence homology with *SMN2* (MIM: 601627)(16); *GATA1* (MIM: 305371), which represents a somatic variant not applicable to germline carrier screening; and *MT-RNR1* (MIM: 561000), a mitochondrial gene with distinct mitochondrial inheritance patterns that differ fundamentally from Mendelian inheritance.

All AR (n=222) and XL (n=25) genes from this list were automatically included in our study, as these inheritance patterns align with traditional carrier screening paradigms. AD genes (n=46), however, required further evaluation. Specifically, we carried out an investigation into the typical age of onset, penetrance, expressivity, and likelihood of primarily *de novo* occurrence for the conditions associated with these genes, to determine their relevance for a reproductive genetic screening context. The inclusion of any AD genes represents a novel framing beyond conventional reproductive carrier screening approaches. To evaluate these genes, we conducted a targeted review using Online Mendelian Inheritance in Man (OMIM) as our primary data source. OMIM was selected for its comprehensive and curated information on gene-disease relationships. Four members of the study team (TYT, SA, JL, SH) reviewed the genes’ associated OMIM entries to assess potential age(s) of onset, penetrance, variable expressivity, and reports of inherited cases (Supplemental Table 3), with supplemental PubMed searches conducted, as needed. A series of criteria were created to determine whether to exclude or include an AD gene in our study. The following inclusion criteria were used: age of onset not restricted to infancy, reduced penetrance, variable expressivity, and documented evidence of multiple affected generations. These combined features would suggest that an individual may be heterozygous for a P/LP variant without experiencing severe symptoms and remain with the possibility of having a severely affected child. The following exclusion criteria were used: reports of mainly or only *de novo* occurrence, severe onset in infancy in almost all cases, and complete penetrance and severe disease in almost all cases, with no phenotypic variability or mild symptoms reported. To adjudicate systematically, the reviewers entered data pertaining to the inclusion/exclusion criteria in a shared Excel document, producing individual judgements on whether to include or exclude the gene. In instances of discordant reviews, JLC resolved the conflict. This review and curation process occurred March to June 2025.

Based on this evaluation, four AD genes (*FAM111A* [MIM: 615292], *SNAP25* [MIM: 600322], *SCN8A* [MIM: 600702], *KCNQ3* [MIM: 602232]) were excluded from two of the subset gene panel analyses because reproductive-age ascertainment of an asymptomatic adult was determined unlikely for the target gene, due to high penetrance, minimal variable expressivity, consistent early age of onset, and classically *de novo* occurrence. These genes are typically associated with lifelong symptomatic disease beginning in infancy, even if diagnosis is delayed until later in life; therefore, it would be expected that an affected adult individual would be aware of their condition and procreative risk. This reasoning has traditionally been the rationale for AD gene exclusion from reproductive genetic screening.(3, 17, 18)

Among the retained AD genes, a subset (*n =* 33) may also have implications for maternal health or obstetric management when either the pregnant individual harbors a pathogenic variant with known personal-health relevance (12) or the fetus harbors the variant, which leads to a secondary obstetric complication. For instance, pregnancy and its associated physiologic stressors can unmask the symptoms of a genetic disease in a previously healthy individual (Supplemental Table 4). To evaluate potential pregnancy and maternal health implications, each gene was reviewed through a targeted literature search.(19–74) Genes were classified as having maternal pregnancy health implications when published evidence suggested a biologically plausible association with maternal health. These implications were categorized as (a) direct maternal effects of the disease/genotype, whereby a pathogenic variant in the mother may lead to maternal symptoms during pregnancy; (b) fetal-mediated effects, in which an affected fetus and its disease manifestations may lead to maternal complications (e.g., maternal mirror syndrome secondary to fetal hydrops); or (c) mechanistic or associative evidence, in which experimental, translational, or clinical data support a plausible link between the disease and adverse pregnancy outcomes.

The objective of the maternal pregnancy health literature review was to identify genes for which published evidence supports clinically relevant associations during pregnancy; it does not establish causality. Because evidence varied across conditions, no minimum level of evidence or study design was required for inclusion. The strength and actionability of the evidence vary substantially by gene, with many genes having minimal evidence through case reports or small case series; maternal health relevance, therefore, was treated as supportive context rather than as an inclusion criterion for reproductive genetic screening. The AD genes included in our study were included primarily for the purpose of ascertaining at-risk fetuses who have an opportunity for early disease intervention. A subsequent potential advantage may be the ascertainment of at-risk mothers with an opportunity for more precise antepartum management.

To assess the impact of including different subset gene lists on heterozygote screening performance (individual-level NNS), we evaluated five different gene panel configurations (see Supplemental Table 5): (a) 293 genes from the treatable fetal findings list described above, (b) only AR and XL genes from the list (n=247), (c) all AD genes from the list (n=46), (d) 293-gene list excluding four AD genes (n=289). and (e) AD genes excluding the four that did not meet inclusion criteria (*FAM111A, SNAP25, SCN8A, KCNQ3*; n=42). These configurations enabled assessment of the NNS to identify one heterozygote using data from: a) combined ancestries across two population datasets (gnomAD v4.1 and AoU), and then b) across various individual genomic ancestral subgroups. These analyses allowed comparison of traditional reproductive genetic screening approaches (AR and XL genes) with expanded panels that include AD genes, all with perinatal treatability as the core gene inclusion criterion.

### Data Sources and Variant Selection

P/LP variants and variants of uncertain significance (VUS) were extracted from ClinVar (VCF format v4.1, GRCh38 reference genome, release date: January 4, 2026) for the 293 genes of interest (Supplemental Table 2). We chose to estimate the frequency of high-confidence, ClinVar submitted variants, as a conservative proxy for all the variants in these genes. ClinVar variants were extracted using gene-symbol annotations in the ClinVar VCF GENEINFO field, using the first-listed gene in this field.

Extracted variants were then annotated in population databases using exact chromosome, position, reference allele, and alternate allele matching for all 293 genes in our proposed reproductive genetic screening gene list. This file was annotated with ClinVar star ratings for downstream analysis purposes. The ClinVar variant lists were then annotated with allele frequency (AF) data, as available, from gnomAD v4.1 and AoU Research Program Variant Annotation Table (VAT) version 8. For the AoU VAT, all transcripts were initially retrieved and then deduplicated to MANE Select or MANE Plus Clinical transcripts to ensure one canonical transcript representation per variant.

gnomAD v4.1 JOINT VCF files, which include 807,162 individuals, were queried chromosome-by-chromosome to determine variant-level allele count (AC) and allele number (AN) and calculate AF across the entire population dataset and across each genomic ancestral subgroup. For X chromosome genes, allele frequencies were determined using XX-specific AC and AN from the gnomAD v4.1 joint callset, restricting the denominator to female individuals, to obtain accurate female carrier AF.

The AoU Research Program BigQuery Genomic Variant Annotation Table (VAT) version 8 analysis included approximately 414,000 participants with short-read whole genome sequencing (srWGS) data, of which 250,071 females (based on self-report) were used for XL gene carrier frequency calculations. The AN for females (250,071 x 2) is an estimate because AoU does not publish female individual-specific variant-level AN. In addition to the P/LP and VUS lists from the ClinVar extraction, we identified computationally-predicted loss-of-function (LoF) variants using a Hail script to query the AoU VAT dataset for variants with a) one of the following high-impact Variant Effect Predictor (VEP) consequence terms (frameshift, stop gained/lost, start lost, splice acceptor/donor, transcript ablation); b) absent ClinVar classification (ClinVar classification empty/not_provided); and c) allele count > 0, meaning observed in the AoU population database. The extraction excluded 15 genes associated with AD gain-of-function P/LP mechanisms (Supplemental Table 6), where LoF variants are likely not P/LP, as well as *SMN1*, *GATA1*, and *MT-RNR1*, which were excluded from all analyses. LoF variants were identified using Hail by filtering the AoU VAT for high-impact consequence terms across all transcripts. Records were subsequently deduplicated to MANE Select or MANE Plus Clinical transcripts to ensure one canonical transcript representation per variant. Genetic ancestry assignments were based on AoU GVS (Genomic Variant Store) ancestry classifications. These GVS fields are generated using genetically inferred ancestry from the genome sequencing data and are therefore analogous to gnomAD ancestry classifications, though the two datasets vary slightly in the total number of groupings depicted.(75–77)

For all downstream NNS analyses, P/LP variants with a 2 star or higher (≥ 2 star) rating were included, and for VUS, only variants with ≥ 2 star rating and rarity, define as variant AF <0.001% (<0.00001), were included. These VUS’ AF were determined based on their AC and AN within each population database (gnomAD JOINT or AoU GVS, respectively). P/LP inclusion required ≥ 2 star ClinVar review status and P/LP clinical significance; variants with conflicting or benign/likely benign assertions were not counted as P/LP. VUS were defined as ClinVar records with an aggregate clinical significance of ‘Uncertain_significance’ and review status ≥2 stars. Records classified as ‘Conflicting_interpretations_of_pathogenicity’ were excluded, as these do not carry the ‘Uncertain_significance’ aggregate designation and additionally fail the ≥2 star criterion which requires multiple submitters with no conflicts. A 5% reclassification rate was applied as part of an additional analysis, to estimate the contribution of potentially pathogenic VUS to carrier frequency.

ClinVar P/LP status was used as an analytic input for estimating heterozygote yield, but variant inclusion is not equivalent to full clinical assessment of phenotype specificity, molecular mechanism, penetrance, or perinatal actionability. Because ClinVar assertions are often gene- or condition-associated rather than tailored to the specific phenotype motivating inclusion of a gene in this framework, estimates should be interpreted as qualifying heterozygote-yield estimates rather than clinically actionable result estimates. To model real-world genetic screening panels, all qualifying P/LP variants, including those outside the intended early-onset phenotype or inheritance pattern, were reviewed and captured by our methods.

### Gene Level Aggregation (AoU and gnomAD) and Statistical Analysis for Number Needed to Screen

Gene-level carrier frequencies were calculated among AoU and gnomAD datasets using Hardy-Weinberg equilibrium calculations. Qualifying variants were treated as independent rare events for purposes of gene-level and panel-level aggregation. This approximation does not model linkage disequilibrium, phase, multiple qualifying variants in the same individual, compound heterozygosity, population-specific structure, or gene-specific clinical actionability. Wilson confidence intervals (CI) were used for carrier frequency and Bootstrap resampling (n=10,000 iterations) was used for NNS CI across 5 dataset scenarios, for 5 subset gene panels, across 8 ancestries, leading to 200 NNS calculations. These CIs represent statistical uncertainty under the implemented model only; they do not capture uncertainty from ClinVar ascertainment, variant reinterpretation, missing variant classes, ancestry structure, phenotype-mechanism matching, or downstream clinical actionability.

Gene-level qualifying allele burden was approximated by summing AC across qualifying variants and dividing by the mean variant-level AN. This calculation is an approximation, as it does not precisely account for the variability of AN across each individual variant, which can affect estimates.

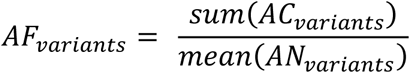

For autosomal variants, heterozygote carrier frequency was then approximated under Hardy-Weinberg equilibrium as:

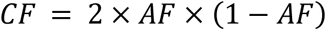

For X-linked genes, analyses were restricted to female participants. Carrier frequency reflects the proportion of females carrying one pathogenic allele on one X chromosome (i.e., heterozygotes), estimated using Hardy-Weinberg equilibrium as CF = 2q(1−q), where q is the aggregate qualifying allele frequency among females. For AoU, female AN was defined as n_females × 2 (500,142 alleles from 250,071 female participants), consistent with diploid X-chromosome counting. For gnomAD, XX-specific AC and AN were used directly from the gnomAD joint callset. These estimates were incorporated into panel-level NNS calculations alongside autosomal carrier frequencies.

Number needed to screen (NNS) was calculated as:

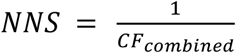

where combined carrier frequency (*CF_combin_*_ed_) was calculated using the following formula:

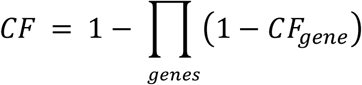

Panel-level NNS estimates incorporate both AR and XL carrier frequencies into a single combined carrier frequency using the product rule: CF_combined = 1 − ∏(1 − CF_gene), where the product is taken across all genes in the panel. This formulation calculates the probability of being a carrier for at least one qualifying gene by subtracting from 1 the probability of being a carrier for none of them, thereby avoiding the overestimation that would result from simply summing individual gene carrier frequencies. AR and XL carrier status carry distinct reproductive implications, with XL carrier females having a 50% risk of an affected son among male offspring (25% of all pregnancies assuming equal sex distribution), compared to a 25% risk of an affected child per pregnancy when both partners are autosomal carriers. Combining them into a single panel-level NNS, however, reflects the practical reality of population-level carrier screening panels, which typically include both AR and XL inheritance types.

### Carrier frequency including sensitivity analysis estimate

To determine how VUS classification may alter NNS results, we conducted an analysis using a conservative, illustrative reclassification rate. We estimated what the gene-level carrier frequencies would be if 5% of the high confidence and rare VUS (≥ 2 stars and AF<0.001%) were to be reclassified as P/LP.

### Analysis Scenarios

The following 5 variant datasets were used to compare results across ancestral groups between AoU and gnomAD: P/LP only (AoU); P/LP + LoF (AoU); P/LP + 5% VUS (AoU); P/LP only (gnomAD); P/LP + 5% VUS (gnomAD). The following 5 subset gene panels were investigated: 1) 293 analyzable perinatal treatable genes; 2) AR + XL Panel (247 genes); 3) AD Panel (46 genes); 4) 42 AD genes (46 AD genes minus 4 complete penetrance genes); and 5) 289 gene panel (all analyzable perinatal treatable genes minus 4 complete penetrance AD genes) (Supplemental Table 5).

We also compared the top 20 genes (by carrier frequency) in each dataset (Supplemental Figures 1 and 2) and cross-referenced whether they appeared on various commercially available carrier screening panels, the ACMG recommended T3 gene list (3) or the more recently published suggested updates to the T3 gene list (18, 78, 79) (Supplemental Tables 7 and 8). Furthermore, we conducted a cumulative contribution plot, showing the share of total qualifying heterozygote yield explained by the top 1, 3, 5, 10 and 20 genes for each of the two datasets (Supplemental Figure 3).

### Number needed to screen analyses

NNS was calculated as an individual-level heterozygote-yield metric for the five subset gene panels (Supplemental Table 5). NNS denotes the estimated number of individuals needed to screen to identify one individual with at least one qualifying heterozygous variant in a gene on the specified list. These estimates do not represent couple-level reproductive risk, affected-pregnancy yield, clinical diagnostic yield, or validation of a clinical screening test. The primary cross-dataset comparison emphasized the shared ClinVar ≥2-star P/LP analysis. The AoU P/LP plus predicted LoF scenario was evaluated separately as an exploratory sensitivity analysis.

### Statistical analysis

All analyses were performed using Python 3.13 (Python Software Foundation) with pandas 2.3.3, NumPy 2.4.1, and SciPy 1.17.0 for carrier frequency calculations and statistical analyses. Data visualization and additional analyses were conducted using R version 4.5.1 (R Core Team, 2025) with tidyverse 2.0.0, ggplot2 3.5.2, dplyr 1.1.4, and data.table 1.17.8.

## RESULTS

### Variant coverage across databases

Among the 293 genes investigated in ClinVar, there were 11,431 P/LP variants with ≥ 2 stars across 266/293 genes with a breakdown as follows: 2 stars (n=9,578 variants); 3 stars (n=1,830 variants); 4 stars (n=23 variants).

Of the 11,431 P/LP variants with ≥ 2 stars identified across 266/293 genes in ClinVar (excluding *MT-RNR1*, *GATA1*, and *SMN1*), 5,351 (46.8%) were present in the AoU VAT and 6,080 (53.2%) were absent. In gnomAD, 6,643/11,431 (58.0%) had non-null AN data and 4,799 (42.0%) were absent (no coverage at the locus); of the 11,431 total, 412 (3.6%) had AC=0 with non-zero AN (covered but not observed) and 6,220 (54.4%) were observed with AC>0. For VUS, 7,311 variants met both the ≥ 2 star and AoU AF<0.001% thresholds in AoU across 271/293 genes (6,946 autosomal + 365 XL); in gnomAD, 12,817 VUS met these criteria using gnomAD AF <0.001% across 282/293 genes (12,387 autosomal and 430 XL).

After flagging the 293 genes for ClinGen haploinsufficiency (HI=3) and triplosensitivity (TS=3), 42/293 (14.3%) flagged true for potential undercounting from dosage sensitivity variation being a likely risk (Supplemental Table 9). Among these 42 flagged genes, 5 are AR, 18 are XL, and 19 are AD.

### Carrier frequency results cross-referenced to commercially available gene panels

The top 20 genes by carrier frequency among our perinatal treatable list for each database—AoU and gnomAD—are notably missing to varying degrees when compared to commercially available carrier screening panels, the ACMG recommended T3 gene list,(3) and more recent publications building upon the ACMG T3 gene list (Supplemental Tables 7 and 8).(78, 79)

To further assess the concentration of carrier frequency yield within the panel, we examined the cumulative contribution of genes ranked by carrier frequency to the total 293-gene panel carrier frequency (Supplemental Figure 3). In both AoU and gnomAD, a small number of genes accounted for a disproportionate share of total yield. In All of Us, *CFTR* alone explained 19.0% of the total panel combined carrier frequency, with the top 3, 5, 10, and 20 genes accounting for 31.8%, 40.7%, 52.3%, and 65.5% respectively. In gnomAD, *CFTR* similarly explained 18.1%, with the top 3, 5, 10, and 20 genes accounting for 32.6%, 42.9%, 56.3%, and 69.5%. The top 5 genes were largely concordant between datasets (AoU: *CFTR*, *F2*, *DHCR7*, *GAA*, *ACADM*; gnomAD: *CFTR*, *CYP21A2*, *F2*, *DHCR7*, *GAA*), with *CYP21A2* ranking second in gnomAD but outside the top 5 in All of Us, likely reflecting differences in ancestry composition between the two cohorts. Notably, even the top 20 genes, which together explain approximately two-thirds of panel-level carrier yield, represent fewer than 7% of the 293-gene panel, highlighting that the remaining genes contribute individually rare, but collectively meaningful, detection of less common conditions.

### Carrier frequency results for genes with possible prenatal intervention

Evaluation of gene-level carrier frequency for a list of 52 genes with prenatal intervention opportunity specifically,(1) demonstrated high carrier frequency for three lysosomal storage disease genes: *GAA* (MIM: 606800), *IDUA* (MIM: 252800), and *GBA1* (MIM: 606463) (Supplemental Table 10).

### Autosomal Dominant Genes

Among the 46 AD genes on the 293 gene list, 42 were considered to meet the following criterion: 1) Perinatal treatability (defined as available/investigational *in utero* interventions or imperative early postnatal treatment), and 2) evidence of either a) variable expressivity, b) incomplete penetrance, or c) plausible asymptomatic adults of procreative age with a risk to have offspring with neonatal symptom onset. The last scenario (c) was defined as an unrestricted age of onset for disease symptoms and demonstrated intrafamilial variability. The remaining four genes not meeting criteria: *FAM111A, SNAP25, SCN8A, KCNQ3*, were conditions with early intervention that also include one or more of the following features: age-of-onset restricted to infancy, complete penetrance, and/or typically occurring de novo. Of the 42 genes meeting criteria, 33 are considered to additionally have potential maternal implications in pregnancy, labor and postpartum, if the mother herself is affected (Supplemental Table 4) and 31 had available data within AoU and gnomAD to determine their carrier frequencies (Supplemental Figure 5). Of note, there are additional AD genes that may have maternal health implications but were not included in our analysis if they did not meet the first criterion of perinatal treatability (e.g. *FBN1* [MIM: 134797], the gene responsible for Marfan syndrome).

The AD-only panel yield was driven by a small number of genes. In AoU, the top three genes (*CYP11B1*, *TSHR*, and *KCNQ1*) had a combined carrier frequency of 0.38%, representing 55.1% of the total AD panel carrier frequency (0.69%). In gnomAD, the top three genes (*PRRT2*, *TSHR*, and *KCNQ1*) had a combined carrier frequency of 0.80%, representing 63.9% of the total AD panel carrier frequency (1.24%). Of note, the top gene in gnomAD (*PRRT2*, CF = 0.46%) ranked fifth in AoU (CF = 0.05%).

Further investigation of variant-level data demonstrated the discrepancy is driven by the known *PRRT2* c.649dup founder variant, which appears as both an insertion (insC, chr16:29813694 G>GC; gnomAD AF = 0.00174 vs AoU AF = 0.00024, 7.3× higher) and a deletion (del, chr16:29813694 GC>G; gnomAD AF = 0.00066 vs AoU AF = 0.00002, 31.6× higher). Together these two representations account for 98.8% of gnomAD’s *PRRT2* allele count (3,562 of 3,606). *PRRT2* c.649dup is a well-documented founder variant with substantially higher prevalence in European-ancestry populations. The gnomAD v4.1 joint dataset is enriched for European-ancestry individuals, whereas the AoU cohort reflects a more ancestrally diverse population, likely explaining the lower observed carrier frequency. This finding illustrates the importance of population-matched reference databases for carrier frequency estimation.

### Number needed to screen analyses

The NNS was highest for the panel that consisted solely of retained AD genes (n = 42; NNS 80-145), especially compared to that of the retained 289-gene list and the AR+XL gene list, which yielded an aggregate NNS between approximately 3 and 4 individuals in the shared ClinVar ≥2-star P/LP analyses across both gnomAD and AoU (Figure 1). When examining the largest proposed panels, including the 293-gene list and the retained 289-gene list excluding *FAM111A, SNAP25, SCN8A,* and *KCNQ3*, ancestry-stratified NNS estimates were approximately 3-6 individuals across different ancestral groups (Figure 2, Supplemental Figure 4). When ancestry was aggregated, the NNS to identify one heterozygote with a qualifying variant across the 289-gene panel remained between 3 and 4 across sensitivity analyses including the P/LP plus LoF and the 5% VUS reclassification scenarios (Figure 3). The AR+XL panel showed similar aggregate NNS estimates of approximately 3-4 individuals (Figure 4). These findings support high individual-level heterozygote yield for the curated list, but they should not be interpreted as couple-level reproductive-risk yield or as evidence of clinical screening-test validation.

**Figure 1.**
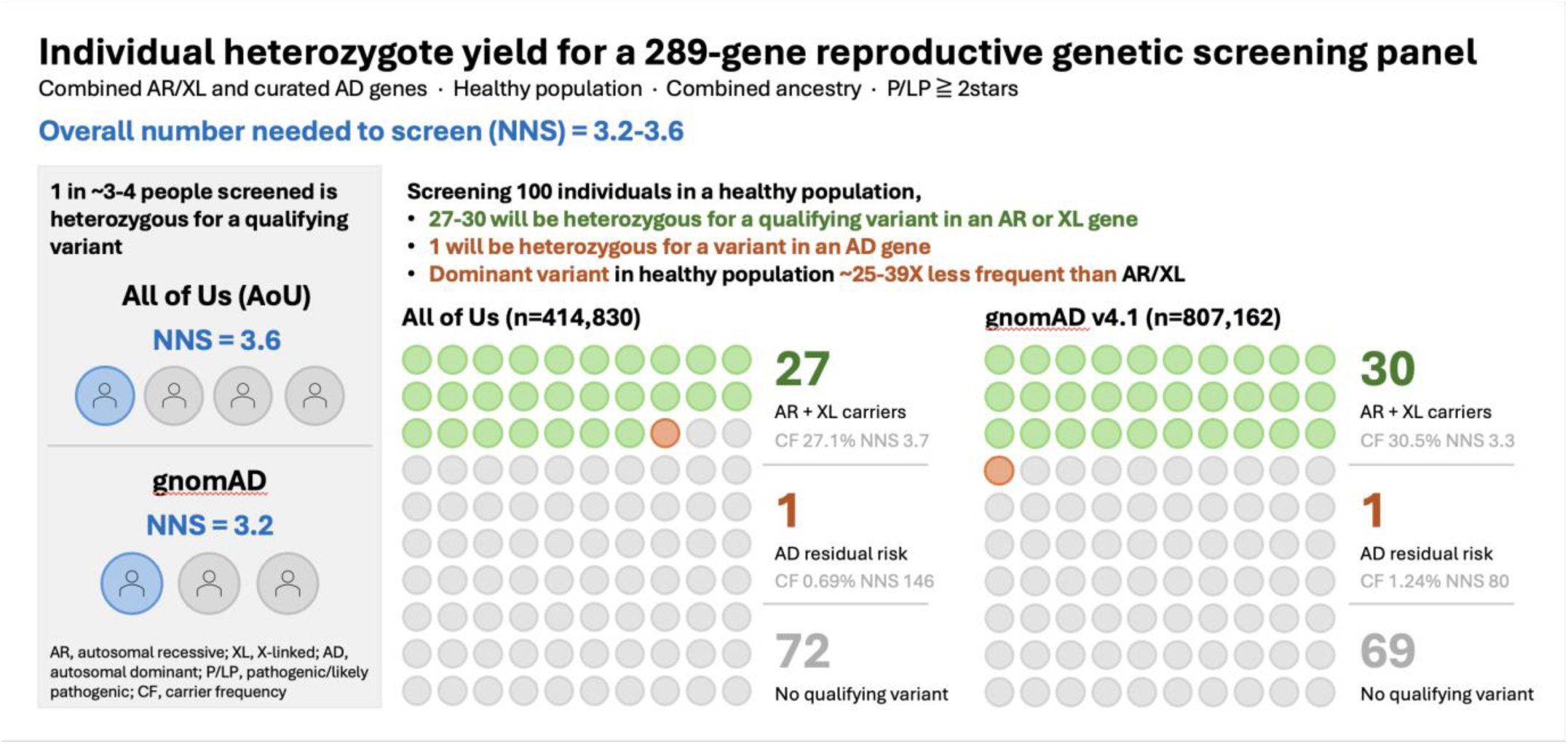
Individual level heterozygote yield (NNS) for a 289-gene reproductive genetic screening panel. Carrier frequency results and NNS were stratified by population database (AoU vs gnomAD). Notably, if 100 healthy individuals in the population were screened, our study found that approximately 1 individual would be heterozygous for a qualifying AD variant, while 27 individuals (AoU) or 30 individuals (gnomAD) would be heterozygous for an AR or XL variant. NNS does not represent couple-level reproductive risk, affected-pregnancy yield, or diagnostic yield. *NNS, number needed to screen; AR, autosomal recessive; XL, X-linked; P/LP, pathogenic/likely pathogenic; AoU, all of us (v8)*.

**Figure 2.**
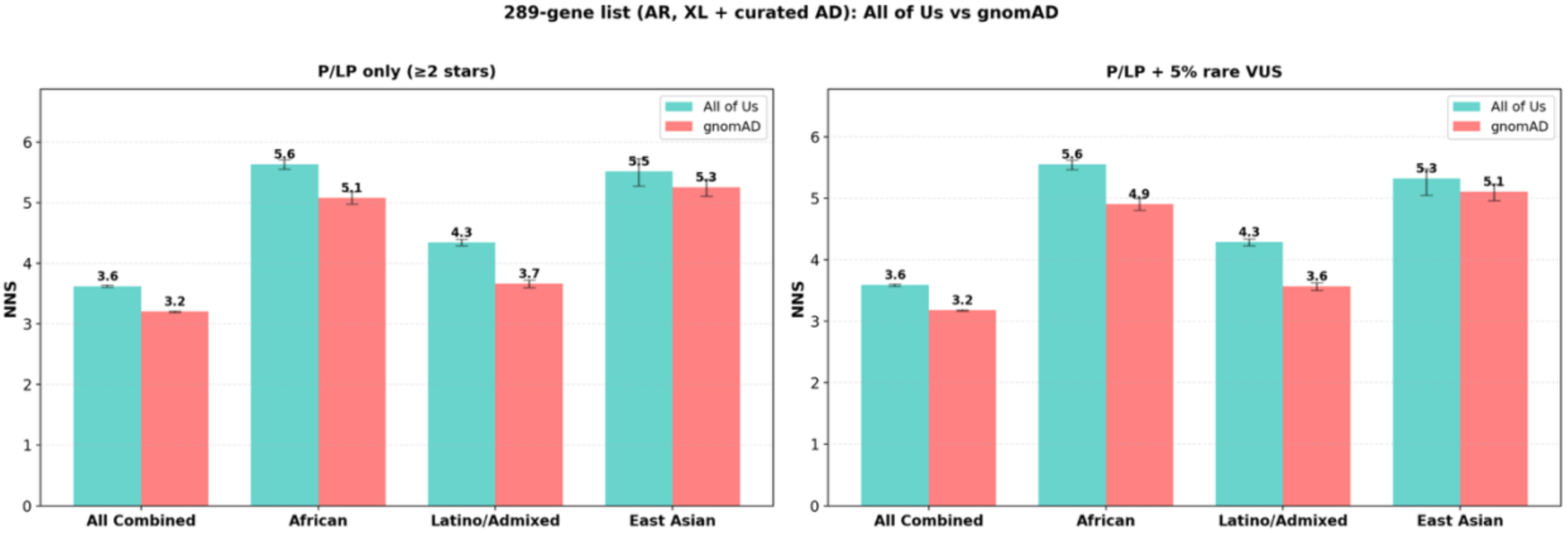
Individual NNS for a 289-gene panel across various ancestral groups and across two variant inclusion datasets. The NNS refers to the individual level heterozygote yield (i.e., NNS to ascertain a single individual with a heterozygous pathogenic or likely pathogenic (P/LP) variant in a gene on the list). Error bars represent the 95% confidence intervals for the NNS. NNS does not represent couple-level reproductive risk, affected-pregnancy yield, or diagnostic yield. *NNS, number needed to screen; AR, autosomal recessive; XL, X-linked; P/LP, pathogenic/likely pathogenic; VUS, variants of uncertain significance*.

**Figure 3.**
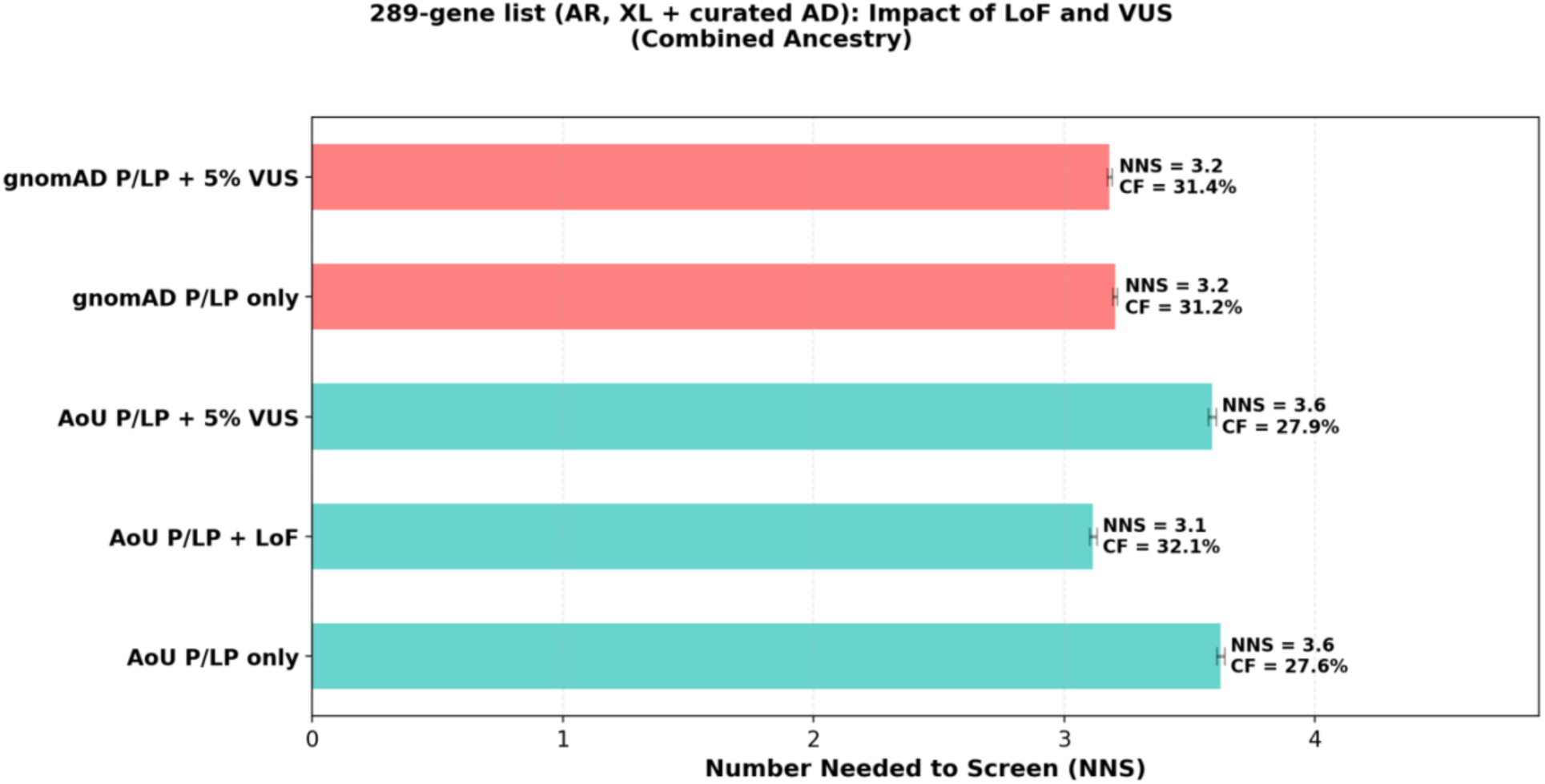
Individual NNS analyses for a 289-gene panel across two biobanks and multiple variant inclusion datasets. P/LP + 5% VUS refers to a hypothetical, illustrative scenario where a 5% conversion rate of rare of high-confidence VUS to P/LP variants in the future is applied to the dataset. LoF (loss of function) refers to the inclusion of non-ClinVar-curated LoF variants that were added after querying the AoU database. Error bars are representative of 95% confidence intervals. NNS does not represent couple-level reproductive risk, affected-pregnancy yield, or diagnostic yield. *CF, carrier frequency; NNS, number needed to screen; AR, autosomal recessive; XL, X-linked; P/LP, pathogenic/likely pathogenic; VUS, variants of uncertain significance; LoF, loss of function*.

**Figure 4.**
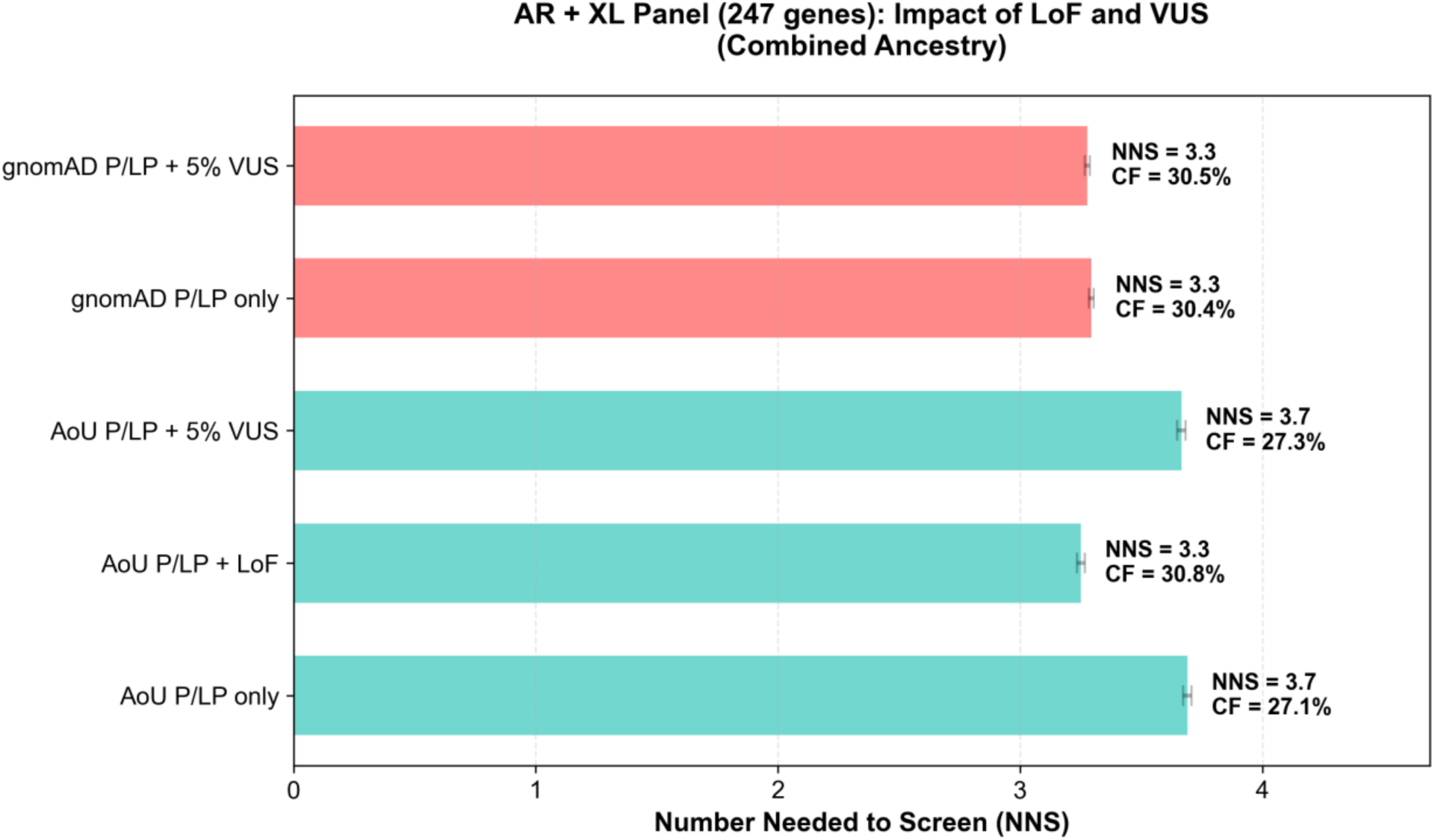
Individual NNS analyses for 247-gene (AR+XL) panel across two biobanks and multiple variant inclusion datasets. P/LP + 5% VUS refers to a hypothetical, illustrative scenario where a 5% conversion rate of rare of high-confidence VUS to P/LP variants in the future is applied to the dataset. LoF (loss of function) refers to the inclusion of non-ClinVar-curated LoF variants that were added after querying the AoU database. Error bars are representative of 95% confidence intervals. NNS does not represent couple-level reproductive risk, affected-pregnancy yield, or diagnostic yield. *CF, carrier frequency; NNS, number needed to screen; AR, autosomal recessive; XL, X-linked; P/LP, pathogenic/likely pathogenic; VUS, variants of uncertain significance; LoF, loss of function*.

The low aggregate NNS should not be interpreted as evidence that every gene contributes equally or that every qualifying heterozygous variant would be clinically actionable. Rather, the framework’s potential value depends on separating yield driven by common genes already present on many panels from incremental yield attributable to genes selected because of perinatal actionability. Future clinical evaluation should assess gene-level contribution, panel gaps, phenotype specificity, and the downstream clinical pathways required for a result to be actionable.

## DISCUSSION

Our prior work demonstrated that a perinatal-actionability gene list can be defined and curated across AR, XL, and selected AD genes when gene inclusion is anchored to potential prenatal intervention or the need for early postnatal treatment. Through this study, we demonstrated that in two large population datasets, this curated list produces high individual-level heterozygote yield, with aggregate NNS estimates of approximately 3–4 for the retained full panel in the shared ClinVar ≥2-star P/LP analyses. The potential value of this framework depends not only on aggregate yield, but also on which genes drive that yield, how many are absent from existing reproductive genetic screening panels, and whether the relevant gene, variant, molecular mechanism, and phenotype combinations are actionable in a reproductive or perinatal context. Accordingly, these analyses support further evaluation of a perinatal-actionability framework for reproductive genetic screening and do not present a validation of a clinical screening test.

Based on our P/LP ≥2-star rating ClinVar variants within gnomAD v4.1 and AoU v8 datasets, 17 genes have both a carrier frequency ≥1/200 (0.5% CF)(3) and have perinatal treatability.(1) 11 genes were shown to have CF ≥1/200 in both datasets; 2 in AoU only; 4 in gnomAD only (Supplemental Tables 11 and 12). Five of the eleven genes remain absent from at least one commercially available gene panel and/or the ACMG recommended T3 gene list(3) (Supplemental Table 11). Therefore, considerable improvements can be made to universal reproductive genetic screening guidelines and recommendations.

When higher-frequency conditions are aggregated with the remainder of the genes on the 293-gene perinatal-actionability list, the NNS to identify one individual with a qualifying heterozygous P/LP variant is 3.20 (CI: 3.193, 3.212) using gnomAD and 3.62 (CI: 3.606, 3.640) using AoU v8 in the shared ClinVar ≥2-star P/LP analyses (Figure 1, Supplemental Table 13). These low individual-level NNS values suggest that the framework may be worth prospective evaluation because it could identify individuals for whom follow-up counseling and testing may clarify reproductive or perinatal risk. However, heterozygote yield is not equivalent to reproductive-risk yield, affected-pregnancy yield, or clinical actionability yield. The low aggregate NNS also implies substantial implementation burden, including potential downstream needs for genetic counseling, partner testing, prenatal diagnostic pathways, confirmatory testing, and clinical follow-up, all of which would require robust clinical infrastructure.

Notably, our observed Ashkenazi Jewish (ASJ) gene carrier frequency for *CFTR* skews higher than reported literature values (typically 0.03-0.04).(80–82) This discrepancy is likely due to our inclusion of the c.1210-11T>G variant in our carrier frequency calculations, a variant that appeared frequently in the gnomAD ASJ cohort (AN=1103). This variant has traditionally been excluded due to its CFTR2 classification as having “varying clinical consequence” as opposed to being strictly “CF-causing.”(83) However, as our study advocates for the screening of genes and variants associated with treatable clinical outcomes, we included this variant given its implications in meconium ileus and availability of effective treatment with *CFTR* modulators.(84, 85) As such, our aggregate NNS estimates may be disproportionately influenced by a small number of high-frequency genes (such as *CFTR*), including genes already included in many conventional carrier-screening panels. Future work should quantify the incremental yield attributable to genes added specifically because of perinatal actionability.

Based on missing variants, the joint VCFs within gnomAD (a combination of exome and genome sequencing) captures the majority of clinically relevant variants used in this study’s carrier frequency calculations. Genes flagged as having a likely underestimation of carrier frequency due to haploinsufficiency scores may benefit from targeted structural variant analysis or long-read sequencing applications to determine their true carrier frequency. Based on the results in this study, both AoU (Unites States-based, ancestrally diverse) and gnomAD (global and European-centric) produce broadly concordant aggregate estimates for our analytic framework.

### Limitations

Several limitations should be considered when evaluating these analytic yield estimates. A notable limitation is that individual-level NNS reflect individual-level positive heterozygote yield estimates and do not directly estimate couple-level reproductive-risk yield, affected pregnancy yield, diagnostic yield, clinical actionability yield, or health-economic utility. The reported NNS values estimate the number of individuals needed to screen to identify one individual with a heterozygous qualifying variant in at least one gene on the panel. These analyses therefore should not be interpreted as validation of a clinical screening test. Prospective clinical evaluation would be needed to determine whether this framework improves reproductive counseling, partner testing, prenatal diagnostic pathways, perinatal management, or clinical outcomes.

A further clinical limitation is that maternal-health implications among retained AD genes are variable and should not be assumed to be uniformly actionable. Maternal-health relevance was treated as supportive context for some genes, not as a universal property of all retained AD genes or as the primary criterion for inclusion. Prospective evaluation will be required to determine which AD gene, variant, phenotype, and pregnancy-context combinations yield clinically meaningful maternal or perinatal management opportunities.

This study has several technical limitations as well, related to the underlying datasets. Since the AoU autosomal variant-level results include only variants observed at least once in the cohort, the exclusion of absent variants may underestimate the contribution of extremely rare or unobserved P/LP alleles. In contrast, gnomAD explicitly reports AN and AC for sites evaluated in joint VCFs, allowing true observed zero frequencies (AC=0 and AN=nonzero) to be distinguished from unobserved variants. Although downstream aggregation excluded variants lacking AN information, differences in database structure, ascertainment, sequencing approach, and cohort composition limit comparability of frequency estimates. For genes with multiple associated phenotypes or molecular mechanisms, our analysis used gene-level ClinVar P/LP classifications and may include variants whose asserted disease mechanism or phenotype is not fully concordant with the perinatal-treatable phenotype used for original gene inclusion. This limitation is especially important because ClinVar P/LP status is not sufficient to establish reproductive or perinatal actionability. Clinical interpretation would require matching the specific variant, molecular mechanism, phenotype, penetrance, and reproductive or perinatal management context. Without this level of variant- and phenotype-specific adjudication, heterozygote-yield estimates may overestimate the proportion of results that would ultimately be clinically actionable.

The exploratory AoU P/LP plus predicted LoF sensitivity analysis used computational high-impact LoF calls, which are not equivalent to clinically curated P/LP variants. Predicted LoF variants are most interpretable for genes and phenotypes in which LoF is an established disease mechanism. Although 15 known gain-of-function genes were excluded from this analysis, the approach still lacks gene-specific mechanism and transcript-level pathogenicity adjudication. The AoU P/LP plus predicted LoF scenario should therefore be interpreted as an exploratory sensitivity analysis rather than as directly analogous to the ClinVar-curated analyses. Additionally, because the primary analyses for this study were based on high-confidence ClinVar P/LP variants, they exclude many VUS that may later be reclassified and may be more prevalent in understudied genes or underrepresented ancestral populations.(86) To evaluate the potential impact of evolving variant classification, we conducted sensitivity analyses modeling partial reclassification of rare VUS. Future work will require periodic reanalysis of gene-level carrier frequency and panel-level NNS as variant curation improves and additional functional and clinical data become available.

Another limitation is the methodology used to calculate gene-level carrier frequencies from variant-level data. Autosomal gene carrier frequency calculations used Hardy-Weinberg equilibrium assumptions with aggregated AF, which does not account for linkage disequilibrium between variants within a gene, compound heterozygosity, population-specific deviations from Hardy-Weinberg equilibrium, phase, or multiple qualifying variants in the same individual. Similarly, subset-panel carrier frequencies and NNS estimates assume independence across genes, which may not hold true in the presence of shared ancestry, consanguinity or population structure. Therefore, all results should be interpreted as analytic approximations rather than observed individual-level carrier probabilities. For XL gene analysis, X-linked carrier frequency estimates assume heterozygosity; homozygous females would have their allele count underestimated by one allele per individual.

Our analysis was weighted toward SNVs and small indels; inclusion of CNVs, repeat expansions, and other complex structural variation was limited by data availability in the two biobanks. This is especially important for genes in which P/LP structural variants account for a substantial proportion of disease-causing alleles and may therefore lead to an underestimation of heterozygote frequency. Detection of *SMN1* variants within AoU and gnomAD, for example, is limited by short-read and whole-exome sequencing technologies given segmental duplication and presence of the homologous paralog *SMN2*.

Omission of such variants can produce clinically meaningful underestimation for some genes(87) and is the reason we excluded *SMN1* from this study’s analyses. Future analyses should integrate structural-variant callsets across population biobanks to more comprehensively capture P/LP variation.

The confidence intervals reported in this study describe statistical uncertainty under the implemented model and are likely narrow because they reflect sampling precision in large reference datasets and the mechanics of bootstrapping. They do not capture broader sources of uncertainty, including ClinVar ascertainment, variant reinterpretation, missing variant classes, unmodeled variant dependence, ancestry misclassification, gene mechanism, phenotype matching, or clinical actionability.

Finally, population biobank cohorts are not fully representative of all global populations. Despite improved ancestral diversity relative to earlier reference datasets, both AoU v8 and gnomAD v4.1 likely still underrepresent certain populations. Carrier frequency estimates for underrepresented groups should therefore be interpreted cautiously, and ongoing efforts to expand diversity in genomic reference resources remain essential. The Goldilocks approach, as developed and outlined by Gruzin and colleagues (2025) (18), offers one strategy for improving equity in carrier-screening gene selections by applying yield thresholds across genetic ancestries and addressing the historical overrepresentation of European genomes in genomic databases.

## Conclusion

While the inclusion of selected AD genes in a reproductive screening framework represents a departure from traditional paradigms, our approach prioritizes identification of perinatal conditions with potential actionability over strict adherence to inheritance pattern alone or individual gene-level carrier frequency. The consistently low individual-level NNS values observed across aggregate analyses and ancestral subgroups support high analytic heterozygote yield and motivate prospective evaluation of whether a perinatal-actionability framework can improve reproductive and perinatal decision-making when paired with appropriate variant interpretation, counseling, partner testing, diagnostic follow-up, and clinical management pathways. In the context of variable guidelines around universal preconception carrier screening, particularly regarding gene selection and panel size, this study provides a clinically informed and data-driven framework for future evaluation. Importantly, these findings should be interpreted as evidence of analytic yield, not as validation of a clinical screening test or as proof that all qualifying heterozygous variants are clinically actionable. This work offers a new perspective for the field of universal preconception screening, with emphasis on perinatal actionability to inform future guideline development and prospective clinical studies.

## Data Availability

All data produced in the present study are available upon reasonable request to the authors.
All analysis code is available for reviewer inspection at https://github.com/JenniferLCohen/Reproductive_Genetic_Screening_for_Perinatal_Treatability.

https://github.com/JenniferLCohen/Reproductive_Genetic_Screening_for_Perinatal_Treatability

## Declaration of generative AI and AI-assisted technologies in the writing process

During the preparation of this work the authors used Anthropic’s ClaudeAI to create and modify Python version 3.13 and R version 4.5.1 scripts that were used for both data analysis and figure creation. Microsoft CoPilot and ClaudeAI were used to improve the clarity of language and to check accuracy/concordance of the language used to describe the methodology and results. After using these tools/services, the authors reviewed and edited the content as needed and take full responsibility for the content of the publication.

## Acknowledgements

The authors acknowledge the following funding sources: Y.T. and Alice Chen Pediatric Genetics and Genomics Research Center; K23HD113824 (JLC); 1K08HD121809-01 (MD). The authors also acknowledge the following students for their early participation in this work: Elle Guillame and Evan Marlowe-Cohen. We gratefully acknowledge All of Us Participants for their contributions, without whom this research would not have been possible. We also thank the National Institutes of Health’s All of Us Research Program for making available the participant data examined in this study.

## Author Contributions

**TYT*:** Writing – original draft (supporting), investigation (supporting), visualization (supporting), writing – review & editing (equal)

**SH*:** Writing – original draft (supporting), investigation (supporting), writing – review & editing (equal)

**XG:** Data curation (equal), software (equal)

**JL:** Investigation (supporting), writing – review and editing (equal)

**SA:** Investigation (supporting), visualization (supporting)

**AL:** Data curation (equal), software (equal)

**CW:** Writing – review and editing (equal)

**NG:** Writing – review and editing (equal)

**SR:** Formal analysis (supporting), writing – review and editing (equal)

**MD:** Formal analysis (supporting), writing – review and editing (equal)

**KW:** Conceptualization (supporting), supervision (supporting), writing – review and editing (equal)

**JLC:** Conceptualization (lead); investigation (lead); writing – original draft (lead); formal analysis (lead), visualization (lead), supervision (lead), writing – review and editing (equal)

## Data and code availability

All analysis code is available for reviewer inspection at https://github.com/JenniferLCohen/Reproductive_Genetic_Screening_for_Perinatal_Treatability.

