## Supplemental tables (1-13) and figures (1-5) for "Expanding reproductive genetic screening through the inclusion of perinatal treatability"

**Table S1.** List of 293 genes with perinatal treatability cross-referenced against the ACMG T3 list,(1) list of genes meeting criteria for carrier screening as determined by Schmitz et al. (286 genes) (2), list of genes meeting criteria for carrier screening given GCF > 1/200 recommendation by Hotakainen (319 genes)(3), and commercially available carrier screening panels: a) Ba – Baylor GeneAware v4 reproductive carrier screen, comprehensive plus panel (611 genes)(4); b) In – Invitae Metabolic Newborn Screening Confirmation Panel (160 genes)(5); c) FuP - Fulgent pre-conception with X-linked disorder and opt-in (427 gene)(6); d) Fulgent ACMG T3 – Beacon 787 Expanded Carrier Screening with X-linked disorders (787 genes)(7); e) My - Myriad Foresight Carrier Screen, universal plus panel (269 genes(8); f) Qu– Quest Qherit198 Disease (199 genes)(9); g) La - LabCorp DynaCare Inheritest 500 Plus Panel (575 genes)(10); h) Bl - Blueprint Comprehensive Reproductive Screen with FMR1 repeat expansion (461 genes)(11); i) Na - Natera H241, Horizon Conditions List (421 genes)(12).

**Table S2.** Full perinatal treatable gene list with inheritance classifications used for the purposes of creating subset gene lists and performing number needed to screen analyses. *AD, autosomal dominant; AR, autosomal recessive; XL, X-linked.*

**Table S3.** List of AD genes curated for our proposed reproductive genetic screening panel. Genes were evaluated and ultimately excluded or included after evaluating penetrance, expressivity, age of onset, whether they were primarily *de novo*, and treatability. *AD, autosomal dominant; GDM, gestational diabetes mellitus; FBG, fetal blood glucose; US, ultrasound; T2DM, Type 2 diabetes mellitus; HI/HA, Hyperinsulinism/hyperammonemia; PPROM, preterm, premature rupture of membranes; PPH, post-partum hemorrhage.*

**Table S4** (see separate word document). Table showing list of 33 autosomal dominant genes with maternal implications antepartum and postpartum if mother is affected, or if mother carries a relevant pathogenic/likely pathogenic variant. This table supports but does not define inclusion criteria for autosomal dominant genes among reproductive genetic screening. *PPROM, preterm premature rupture of membranes.*

**Table S5.** Five subset gene panels created for the number-needed-to-screen (NNS) analysis. *AD, autosomal dominant; AR, autosomal recessive; XL, X-linked.*

**Table S6.** The 15 autosomal dominant genes with a primary mechanism of pathogenicity of gain-of-function. These genes were filtered out of the list of 293 genes of interest for the computational loss-of-function variant analysis conducted within the All of Us database. *AD, autosomal dominant; AR, autosomal recessive; XL, X-linked.*

**Table S7.** Top 20 genes by carrier frequency within the gnomAD database, cross-referenced against the ACMG T3 list,(1) list of genes meeting criteria for carrier screening as determined by Schmitz et al. (286 genes) (2), list of genes meeting criteria for carrier screening given GCF > 1/200 recommendation by Hotakainen (319 genes)(3), and commercially available carrier screening panels: a) Ba – Baylor GeneAware v4 reproductive carrier screen, comprehensive plus panel (611 genes)(4); b) In – Invitae Metabolic Newborn Screening Confirmation Panel (160 genes)(5); c) FuP - Fulgent pre-conception with X-linked disorder and opt-in (427 gene)(6); d) Fulgent ACMG T3 – Beacon 787 Expanded Carrier Screening with X-linked disorders (787 genes)(7); e) My - Myriad Foresight Carrier Screen, universal plus panel (269 genes(8); f) Qu– Quest Qherit198 Disease (199 genes)(9); g) La - LabCorp DynaCare Inheritest 500 Plus Panel (575 genes)(10); h) Bl - Blueprint Comprehensive Reproductive Screen with FMR1 repeat expansion (461 genes)(11); i) Na - Natera H241, Horizon Conditions List (421 genes)(12). *AD, autosomal dominant; AR, autosomal recessive; XL, X-linked.*

**Table S8.** Table showing top 20 genes by carrier frequency within the All of Us (AoU) database, cross-referenced against the ACMG T3 list,(1) list of genes meeting criteria for carrier screening as determined by Schmitz et al. (286 genes) (2), list of genes meeting criteria for carrier screening given GCF > 1/200 recommendation by Hotakainen (319 genes)(3), and commercially available carrier screening panels: a) Ba – Baylor GeneAware v4 reproductive carrier screen, comprehensive plus panel (611 genes)(4); b) In – Invitae Metabolic Newborn Screening Confirmation Panel (160 genes)(5); c) FuP - Fulgent pre-conception with X-linked disorder and opt-in (427 gene)(6); d) Fulgent ACMG T3 – Beacon 787 Expanded Carrier Screening with X-linked disorders (787 genes)(7); e) My - Myriad Foresight Carrier Screen, universal plus panel (269 genes(8); f) Qu– Quest Qherit198 Disease (199 genes)(9); g) La - LabCorp DynaCare Inheritest 500 Plus Panel (575 genes)(10); h) Bl - Blueprint Comprehensive Reproductive Screen with FMR1 repeat expansion (461 genes)(11); i) Na - Natera H241, Horizon Conditions List (421 genes)(12). *AD, autosomal dominant; AR, autosomal recessive; XL, X-linked.*

**Table S9.** Table showing genes out of original 293-gene list that were flagged as likely undercounted due to classification of haploinsufficiency (HI = 3) or triplosensitivity (TS = 3) as assessed in ClinGen. *HI, haploinsufficiency; TS, triplosensitivity.*

**Table S10**. Table showing the gene level carrier frequency for a list of 52 genes with possible prenatal intervention, including emerging interventions and those in clinical trials. “No qualifying variants” refers to cases in which a gene had no ClinVar pathogenic/likely pathogenic variants meeting the ≥2 star threshold in their respective dataset (AoU or gnomAD) and was therefore excluded from gene-level carrier frequency calculation. The NNS for the possible prenatal intervention gene list (n = 52) in the AoU dataset was 6.9, 95% CI: [6.8, 7.0]; the NNS for the gnomAD dataset was 6.6, 95% CI [6.6, 6.7]. NNS refers to the individual level heterozygote yield (i.e., NNS to ascertain a single individual with a heterozygous pathogenic or likely pathogenic (P/LP) variant in a gene on the list). *NNS, number needed to screen; AoU, All of Us (v8); CI, confidence interval.*

**Table S11**. Table showing genes with carrier frequency >1/200 across both AoU and gnomAD, cross-referenced against the ACMG T3 list,(1) list of genes meeting criteria for carrier screening as determined by Schmitz et al. (286 genes) (2), list of genes meeting criteria for carrier screening given GCF > 1/200 recommendation by Hotakainen (319 genes)(3), and commercially available carrier screening panels: a) Ba – Baylor GeneAware v4 reproductive carrier screen, comprehensive plus panel (611 genes)(4); b) In – Invitae Metabolic Newborn Screening Confirmation Panel (160 genes)(5); c) FuP - Fulgent pre-conception with X-linked disorder and opt-in (427 gene)(6); d) Fulgent ACMG T3 – Beacon 787 Expanded Carrier Screening with X-linked disorders (787 genes)(7); e) My - Myriad Foresight Carrier Screen, universal plus panel (269 genes(8); f) Qu– Quest Qherit198 Disease (199 genes)(9); g) La - LabCorp DynaCare Inheritest 500 Plus Panel (575 genes)(10); h) Bl - Blueprint Comprehensive Reproductive Screen with FMR1 repeat expansion (461 genes)(11); i) Na - Natera H241, Horizon Conditions List (421 genes)(12). *AD, autosomal dominant; AR, autosomal recessive; XL, X-linked*

**Table S12**. List of genes in gnomAD and AoU that were found to have a carrier frequency of >1/200 in either one or both databases. *T, true for having carrier frequency > 1/200; F, false for having carrier frequency > 1/200; AoU, All of Us (v8).*

**Table S13.** Table showing NNS, or the individual level heterozygote yield (i.e., NNS to ascertain a single individual with a heterozygous pathogenic or likely pathogenic (P/LP) variant in a gene on the list), depending on the variant scenario for different variant dataset scenarios (P/LP only, P/LP + LoF, P/LP + 5% rare VUS) across the 5 subset gene panels and ancestry groups. gnomAD and AoU used different classifications of ancestry groups. For AoU: ALL (aggregated ancestral groups), AFR (African), AMR (admixed American), EAS (East Asian), MID (Middle Eastern), SAS (South Asian), OTH (individuals that do not fall into any of the other pre-determined groups). For gnomAD: Total (aggregated ancestral groups), AFR (African/African American), ASJ (Ashkenazi Jewish), EAS (East Asian), FIN (Finnish), NFE (non-Finnish European), SAS (South Asian), MID (Middle Eastern), Remaining (individuals that do not fall into any of the other pre-determined groups). 'Genes with data' refers to genes with available allele frequency data among the high-confidence ClinVar variants queried within the All of Us v8 and/or gnomAD v4.1 datasets. *NNS, number needed to screen; P/LP, pathogenic/likely pathogenic; LoF, loss of function; VUS, variant of uncertain significance; AR, autosomal recessive; XL, X-linked; AoU, All of Us (v8); CF, carrier frequency; CI, confidence interval.*

### Supplemental Table 1

| Perinatal treatability gene list | ACMG T3 | Schmitz 2024 | Hotakainen 2025 | Ba | In | FuP | FuT3 | My | Qu | La | Bl | Na |
|---|---|---|---|---|---|---|---|---|---|---|---|---|
| *COL1A1* | | | | | | | | | | | | |
| *COL1A2* | | | | | | | | | | | | |
| *GAA* | ● | ● | ● | ● | ● | ● | ● | ● | ● | ● | ● | ● |
| *LIPA* | | | | ● | | ● | ● | ● | | ● | ● | ● |
| *IDUA* | ● | | ● | ● | ● | ● | ● | ● | ● | ● | ● | ● |
| *IDS* | | | | ● | ● | ● | ● | ● | ● | ● | ● | ● |
| *GALNS* | | | | ● | | ● | ● | | | ● | ● | ● |
| *ARSB* | | | | ● | | ● | ● | | ● | | ● | ● |
| *GUSB* | | | | ● | | ● | ● | | | ● | | ● |
| *GBA1* | ● | ● | ● | ● | | ● | ● | ● | ● | ● | ● | ● |
| *EDA* | | | | ● | | ● | ● | | | ● | ● | ● |
| *KCNH2* | | | | | | | | | | | | |
| *SCN5A* | | | | | | | | | | | | |
| *CACNA1C* | | | | | | | | | | | | |
| *TPO* | | ● | ● | ● | | ● | ● | | | ● | ● | |
| *SLC16A2* | | | | | | ● | ● | | | ● | | |
| *RPS19* | | | | | | | | | | | | |
| *RPL5* | | | | | | | | | | | | |
| *RPS10* | | | | | | | | | | | | |
| *RPS24* | | | | | | | | | | | | |
| *FGB* | | | | | | | | | | | | |
| *FGA* | | ● | ● | | | | | | | | | |
| *FGG* | | | ● | | | | | | | | | |

| | | | | | | | | | | | | |
|---|---|---|---|---|---|---|---|---|---|---|---|---|
| *F2* | | | | • | | | • | • | | | | |
| *F5* | | | | • | | | • | • | | • | | |
| *F7* | | | • | | | | • | | | | | |
| *F8* | • | | | • | | • | • | • | • | | | |
| *F9* | • | | | • | | • | • | • | • | • | • | • |
| *F10* | | | | | | | | | | | | |
| *F11* | | | | • | | • | • | • | • | | • | • |
| *F12* | | | | | | | | | | | | |
| *F13A1* | | | | | | | | | | | | |
| *F13B* | | | | | | | | | | | | |
| *HBA1* | • | • | | | | • | • | | • | • | • | • |
| *HBA2* | • | • | | | | • | • | | • | • | • | • |
| *IL2RG* | | | | • | | • | • | • | | • | • | • |
| *MMACHC* | • | • | • | • | • | • | • | • | • | • | • | • |
| *HLCS* | | | | • | • | • | • | • | • | • | • | • |
| *MMAA* | | | | • | • | • | • | • | • | • | • | • |
| *MMAB* | | | | • | • | • | • | • | • | • | • | • |
| *MMADHC* | | | | • | • | • | • | | | • | • | • |
| *NANS* | | | | | | | | | | | | |
| *PHGDH* | | | | • | | • | • | • | | • | • | • |
| *DHCR7* | • | • | • | • | | • | • | • | | • | • | • |
| *TRMU* | | | | • | | • | • | • | | • | | • |
| *ALDH7A1* | | | | • | • | | • | | | • | • | • |
| *SCN8A* | | | | | | | | | | | | |
| *TSC1* | | | | | | | | | | | | |
| *TSC2* | | | | | | | | | | | | |
| *MAGED2* | | | | | | | | | | | | |

| | | | | | | | | | | | | |
|---|---|---|---|---|---|---|---|---|---|---|---|---|
| *ACE* | | • | • | | | | | | | | | |
| *CFTR* | • | • | • | • | • | • | • | • | • | • | • | • |
| *MMUT* | • | • | • | • | | | | • | • | • | | |
| *KCNQ1* | | | | | | | | | | | | |
| *PDX1* | | | | | | | | | | | | |
| *TBX19* | | | | | | | • | | | | | |
| *POMC* | | | | | | | | | | | | |
| *MRAP* | | | | | | | | | | | | |
| *PCSK1* | | | | | | | | | | | | |
| *CYP21A2* | • | • | • | • | | • | • | • | • | • | • | • |
| *HSD3B2* | | | | • | | • | • | | | • | • | • |
| *STAR* | | | | • | | • | • | • | | • | • | • |
| *CYP11A1* | • | | | • | | • | • | • | • | • | | |
| *NR0B1* | • | | | | • | • | • | | • | • | • | |
| *NR5A1* | | | | | | | | | | | | |
| *AVPR2* | | | | • | | | • | | | • | • | |
| *AQP2* | | | | • | | • | • | | | | | • |
| *TSHB* | | | | • | | • | • | | | | | |
| *TRHR* | | | | | | | • | | | | | |
| *IGSF1* | | | | | | | • | | | | | |
| *SOX3* | | | | | | | | | | | | |
| *POU1F1* | | | | • | | | • | | | | | |
| *HESX1* | | | | | | | | | | | | |
| *PROP1* | | | | • | | • | • | • | • | • | • | • |
| *LHX3* | | | | • | | • | • | | | • | • | • |
| *LHX4* | | | | | | | | | | | | |
| *OTX2* | | | | | | | | | | | | |

| | | | | | | | | | | | |
|---|---|---|---|---|---|---|---|---|---|---|---|
| *KCNJ11* | | | ● | | ● | ● | ● | ● | | | ● |
| *ABCC8* | ● | ● | ● | | ● | ● | ● | ● | ● | ● | ● |
| *GCK* | | | | | | | | | | | |
| *GLUD1* | | | | | | | | | | | |
| *HNF4A* | | | | | | | | | | | |
| *HNF1A* | | | | | | | | | | | |
| *FAM111A* | | | | | | | | | | | |
| *CYP11B1* | | | | | ● | ● | ● | | ● | ● | ● |
| *CLCN2* | | | | | | | | | | | |
| *KCNJ5* | | | | | | | | | | | |
| *CYP11B2* | | | ● | | ● | ● | ● | | | ● | |
| *INS* | | | | | | | | | | | |
| *GATA4* | | | | | | | | | | | |
| *GATA6* | | | | | | | | | | | |
| *RFX6* | | | | | | | | | | | |
| *SLC25A15* | | | ● | ● | ● | ● | | | ● | | ● |
| *TRPM6* | | | | | | ● | | | | | |
| *PTF1A* | | | | | | | | | | | |
| *ZFP57* | | | | | | | | | | | |
| *EIF2AK3* | | | ● | | ● | ● | | | | | ● |
| *NEUROG3* | | | | | | | | | | | |
| *IER3IP1* | | | | | | | | | | | |
| *GLIS3* | | | | | | | | | | | |
| *PAX6* | | | | | | | | | | | |
| *SLC2A2* | | | | | ● | ● | | | | | |
| *NKX2-2* | | | | | | | | | | | |
| *MNX1* | | | | | | | | | | | |

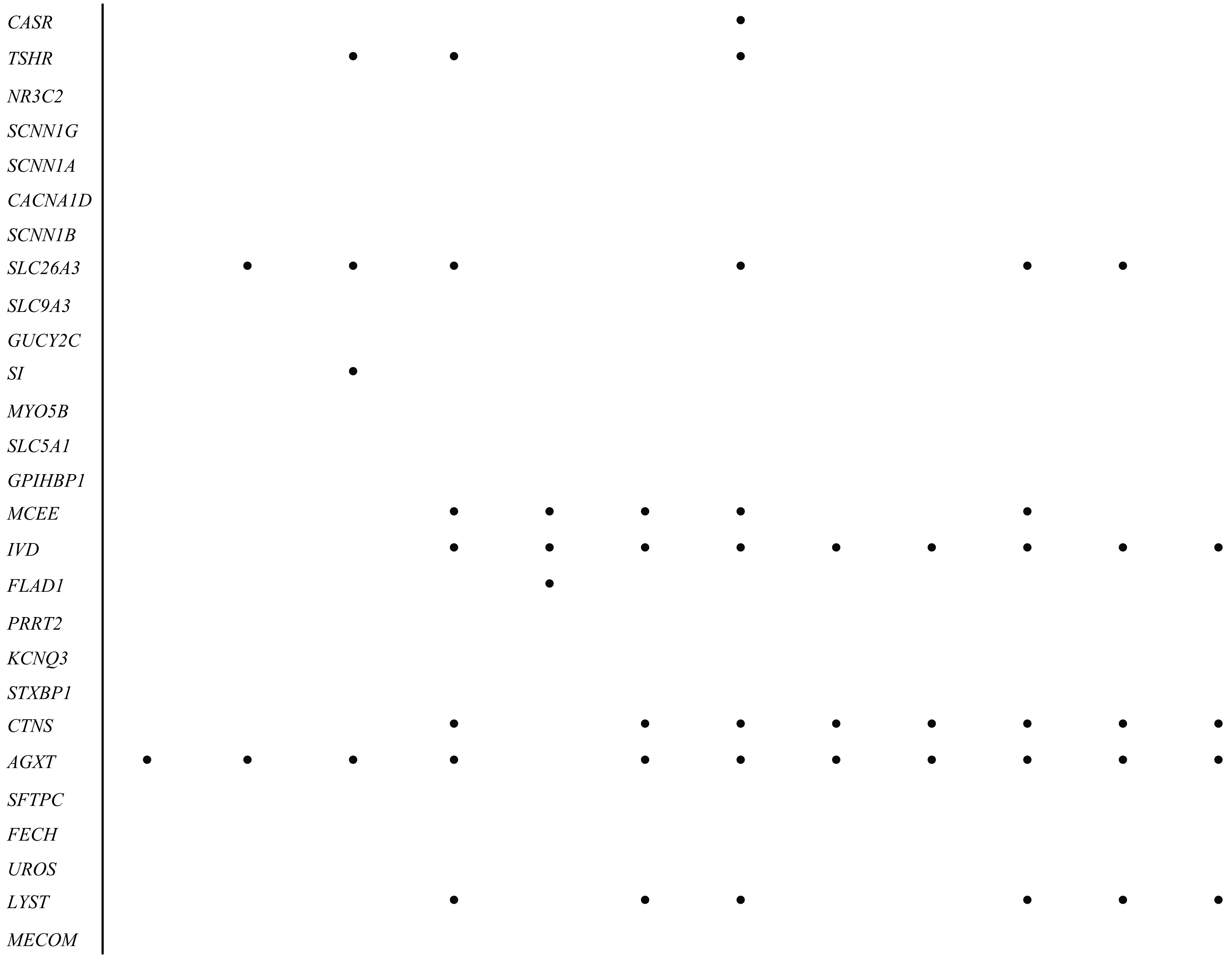
CASR
TSHR
NR3C2
SCNN1G
SCNN1A
CACNA1D
SCNN1B
SLC26A3
SLC9A3
GUCY2C
SI
MYO5B
SLC5A1
GPIHBP1
MCEE
IVD
FLAD1
PRRT2
KCNQ3
STXBP1
CTNS
AGXT
SFTPC
FECH
UROS
LYST
MECOM

| | | | | | | | | | | |
|---|---|---|---|---|---|---|---|---|---|---|
| *MPL* | • | • | • | • | • | • | • | • | • | • |
| *THPO* | | | | | | | | | | |
| *RBM8A* | • | | | | | | | | | |
| *BLNK* | | | | | | | | | | |
| *BTK* | | | • | | • | | | • | | • |
| *CD79A* | | | | | | | | | | |
| *IGHM* | | | | | | | | | | |
| *TCF3* | | | | | | | | | | |
| *FNIP1* | | | | | | | | | | |
| *NLRC4* | | | | | | | | | | |
| *IL1RN* | | | | | | | | | | |
| *NLRP3* | | | | | | | | | | |
| *USP18* | | | | | | | | | | |
| *CD40* | | | • | | | | | • | • | |
| *CD40LG* | | | • | • | • | | | • | • | • |
| *CIITA* | | | • | • | • | | | • | • | • |
| *DOCK8* | | | | | • | | | • | | |
| *IKZF1* | | | | | | | | | | |
| *IKBKG* | | | | | | | | | | |
| *RFXANK* | | | | | • | | | • | | |
| *RFX5* | | | | | • | | | • | | |
| *RFXAP* | | | | | • | | | • | | |
| *RMRP* | • | • | • | • | • | | • | • | • | • |
| *STAT3* | | | | | | | | | | |
| *WAS* | | | • | • | • | | | • | | • |
| *ZAP70* | | | | | • | | | • | | |
| *LCK* | | | | | • | | | • | | |

| | | | | | | | | | | | | |
|---|---|---|---|---|---|---|---|---|---|---|---|---|
| *PGM3* | | | | • | | | • | | | • | | |
| *CYBA* | | | | • | | • | • | • | | • | • | • |
| *CYBC1* | | | | | | | | | | | | |
| *NCF1* | | • | • | | | | | | | | | |
| *NCF2* | | | | • | | | • | | | • | • | |
| *NCF4* | | | | | | | • | | | • | | |
| *CYBB* | | | | • | | • | • | | | • | • | • |
| *ITGB2* | | | | | | | | | | | | |
| *IRAK4* | | | | | | | | | | | | |
| *MYD88* | | | | | | | | | | | | |
| *LRBA* | | | | | | | | | | | | |
| *FASLG* | | | | | | | | | | | | |
| *FOXP3* | | | | • | | | • | | | • | | • |
| *PRF1* | • | • | • | • | | • | • | • | • | • | • | • |
| *RAB27A* | | • | | | | | | | | | | |
| *STX11* | | | | • | | | • | | | • | | • |
| *STXBP2* | | | | • | | | • | | | • | • | • |
| *UNC13D* | | | | | | | • | | | • | • | |
| *FAS* | | | | | | | | | | | | |
| *IL10RA* | | | | | | | | | | | | |
| *IL10RB* | | | | | | | | | | | | |
| *SH2D1A* | | | | | | | | | | | | |
| *XIAP* | | | | | | | | | | | | |
| *ADA* | | | | • | | • | • | • | | • | • | • |
| *AK2* | | | | | | | • | | | • | | |
| *CD3D* | | | | • | | | • | | | • | | |
| *CD3E* | | | | • | | | • | | | • | | |

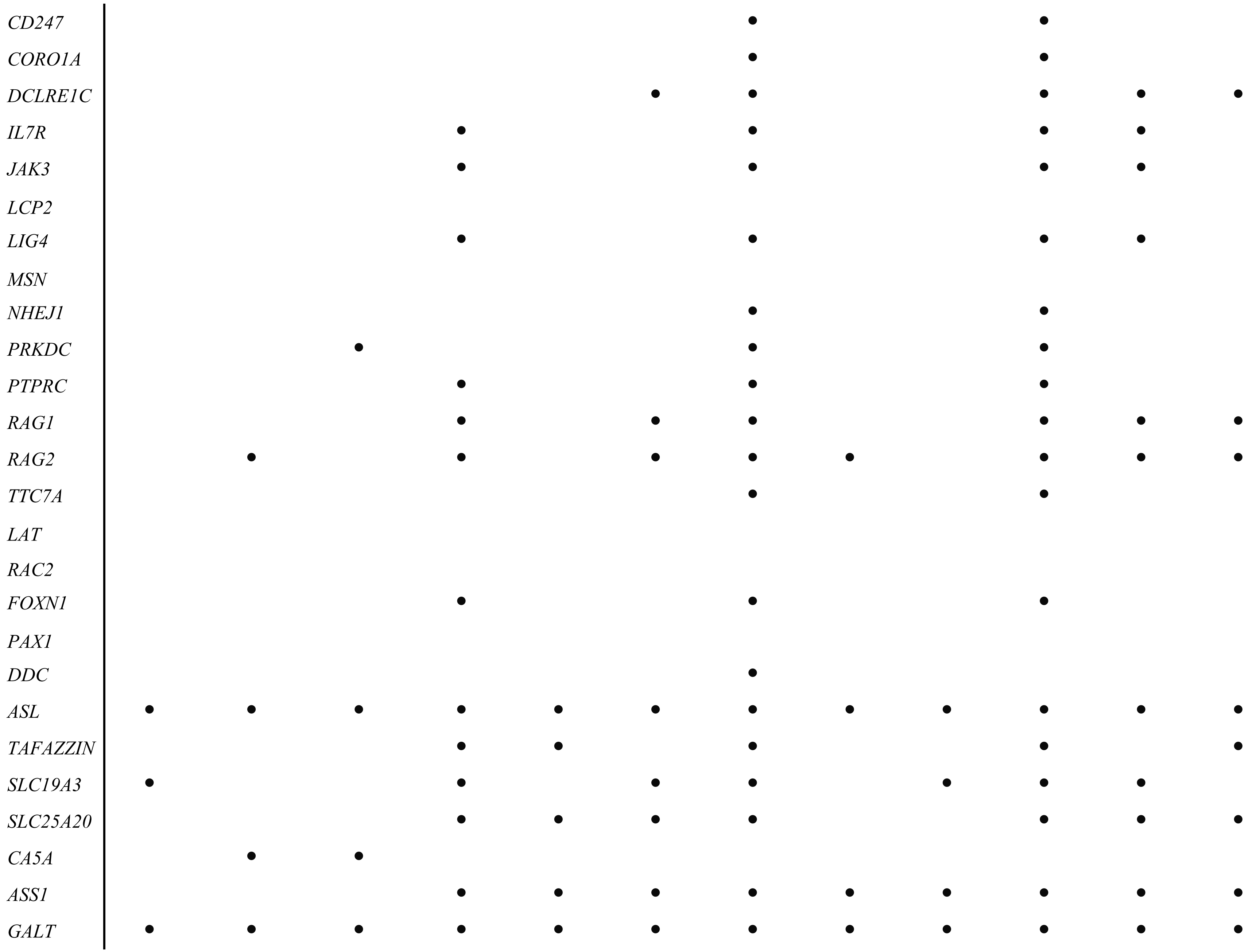
CD247
CORO1A
DCLRE1C
IL7R
JAK3
LCP2
LIG4
MSN
NHEJ1
PRKDC
PTPRC
RAG1
RAG2
TTC7A
LAT
RAC2
FOXN1
PAX1
DDC
ASL
TAFAZZIN
SLC19A3
SLC25A20
CA5A
ASS1
GALT

| | | | | | | | | | | | | |
|---|---|---|---|---|---|---|---|---|---|---|---|---|
| *CPS1* | | | | ● | ● | ● | ● | ● | | ● | ● | ● |
| *CPT2* | ● | ● | ● | ● | ● | ● | ● | ● | | ● | ● | ● |
| *DLD* | ● | ● | ● | ● | ● | ● | ● | ● | ● | ● | ● | ● |
| *FBP1* | | | | ● | | | ● | | | ● | | |
| *GOT2* | | | | | | | | | | | | |
| *ETFA* | | | | ● | ● | ● | ● | | | ● | ● | ● |
| *ETFB* | | | | ● | ● | ● | ● | | | ● | ● | ● |
| *ETFDH* | | ● | ● | ● | ● | ● | ● | | | ● | ● | ● |
| *AMT* | | | | ● | ● | ● | ● | ● | ● | ● | ● | ● |
| *GLDC* | | ● | ● | ● | ● | ● | ● | ● | ● | ● | ● | ● |
| *ALDOB* | ● | ● | ● | ● | | ● | ● | | ● | ● | ● | ● |
| *FAH* | ● | ● | ● | ● | ● | ● | ● | ● | ● | ● | ● | ● |
| *HMGCL* | | | | ● | ● | ● | ● | ● | ● | ● | ● | ● |
| *HADHA* | | ● | ● | ● | ● | ● | ● | ● | ● | ● | ● | ● |
| *HADHB* | | | | ● | ● | | ● | | | ● | ● | ● |
| *BCKDHA* | | | | ● | ● | ● | ● | ● | ● | ● | ● | ● |
| *BCKDHB* | ● | ● | ● | ● | ● | ● | ● | ● | ● | ● | ● | ● |
| *DBT* | | | | ● | ● | ● | ● | ● | ● | ● | ● | ● |
| *ACADM* | ● | ● | ● | ● | ● | ● | ● | ● | ● | ● | ● | ● |
| *AP1S1* | | | | ● | | | ● | | | | ● | ● |
| *ATP7A* | | | | ● | | ● | ● | ● | ● | ● | ● | ● |
| *ACAD9* | | | | ● | | ● | ● | | | ● | ● | |
| *ECHS1* | | | | | ● | | | | | | | |
| *MOCS1* | | | | ● | | | | | | ● | | ● |
| *MPI* | | | | ● | | ● | ● | ● | ● | ● | ● | ● |
| *NAGS* | | | | ● | ● | ● | ● | | | ● | ● | ● |

| | | | | | | | | | | | | |
|---|---|---|---|---|---|---|---|---|---|---|---|---|
| *OAT* | | • | • | • | • | • | • | • | • | • | • | • |
| *OTC* | • | | | • | • | • | • | • | • | • | • | • |
| *PGM1* | | | | | • | | | | | | | |
| *COQ2* | | | | | | | | | | | | |
| *COQ4* | | • | • | | | | • | | | | | |
| *COQ9* | | | | | | | | | | | | |
| *PDSS2* | | | | | | | | | | | | |
| *PDSS1* | | | | | | | | | | | | |
| *COQ8A* | | | • | | | | | | | | | |
| *COQ6* | | | | | | | | | | | | |
| *COQ7* | | | | | | | | | | | | |
| *COQ5* | | | | | | | | | | | | |
| *PDHA1* | | | | | | • | • | | | • | • | • |
| *PDHX* | | | | | | | • | | | • | | |
| *DLAT* | | | | | | | • | | | | | |
| *PDHB* | | • | | • | | • | • | | | • | • | • |
| *PDP1* | | | | | | | • | | | • | | |
| *PCCA* | | | | • | • | • | • | • | • | • | • | • |
| *PCCB* | | • | • | • | • | • | • | • | • | • | | • |
| *PNPO* | | | | • | • | | • | | | • | • | |
| *ALPL* | • | • | • | • | | • | • | • | • | • | • | • |
| *PLPBP* | | | | | | | | | | | | |
| *SLC35A2* | | | | | | | | | | | • | |
| *OXCT1* | | • | • | | | | | | | | | |
| *ACADVL* | • | • | • | • | • | • | • | • | | • | • | • |
| *FOLR1* | | | | | | | • | | | | | |
| *DPAGT1* | | | | | | | | | | | | |

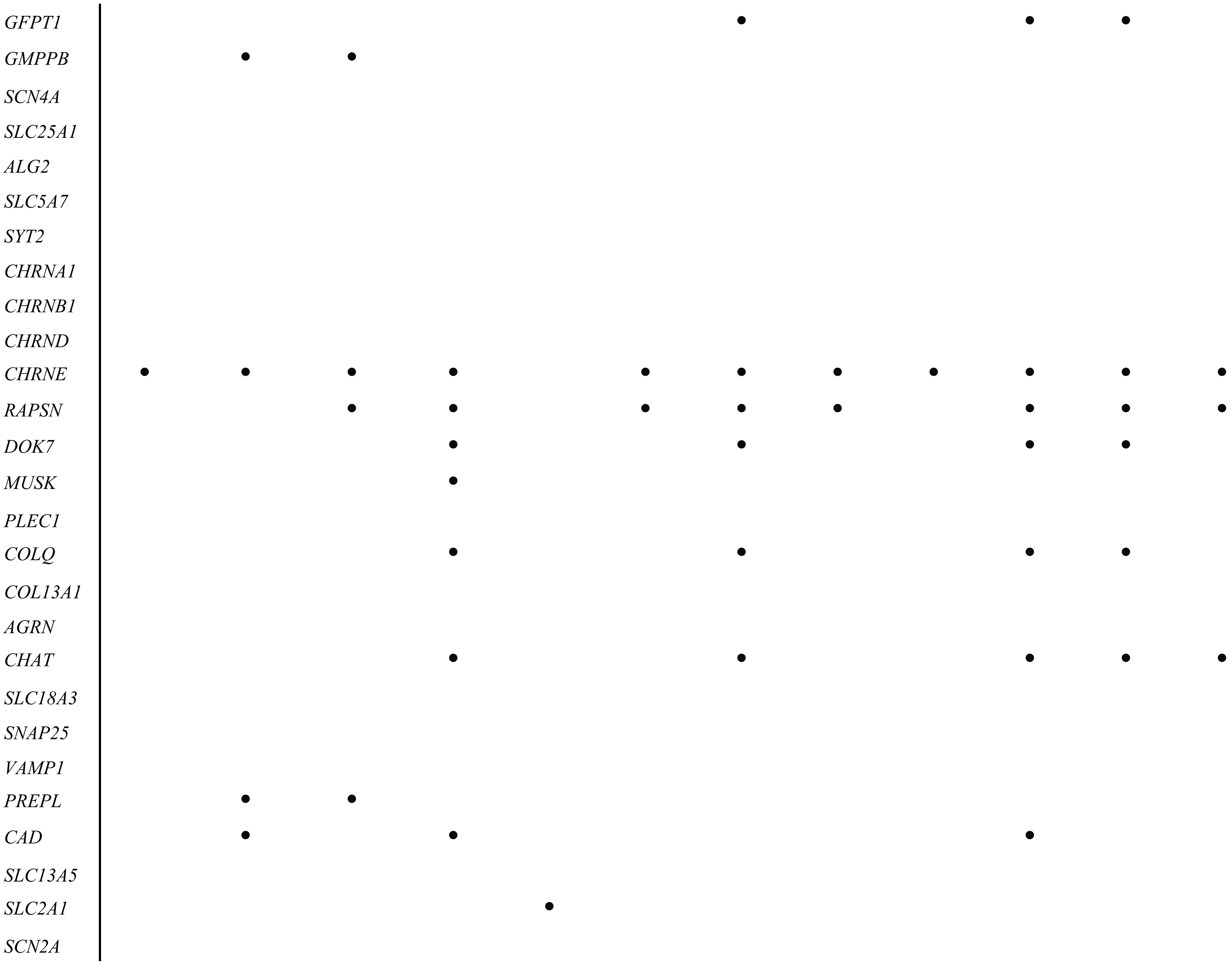
GFPT1
GMPPB
SCN4A
SLC25A1
ALG2
SLC5A7
SYT2
CHRNA1
CHRNB1
CHRND
CHRNE
RAPSN
DOK7
MUSK
PLEC1
COLQ
COL13A1
AGRN
CHAT
SLC18A3
SNAP25
VAMP1
PREPL
CAD
SLC13A5
SLC2A1
SCN2A

| | |
|---|---|
| *KCNQ2* | |
| *HIBCH* | • |

Supplemental Table 2

| Gene symbol | Inheritance classification for panel analyses | Known inheritance patterns for phenotype of interest |
|---|---|---|
| *ABCC8* | AR | AD;AR |
| *ACAD9* | AR | AR |
| *ACADM* | AR | AR |
| *ACADVL* | AR | AR |
| *ACE* | AR | AR |
| *ADA* | AR | AR |
| *AGRN* | AR | AR |
| *AGXT* | AR | AR |
| *AK2* | AR | AR |
| *ALDH7A1* | AR | AR |
| *ALDOB* | AR | AR |
| *ALG2* | AR | AR |
| *ALPL* | AR | AR |
| *AMT* | AR | AR |
| *AP1S1* | AR | AR |
| *AQP2* | AR | AD;AR |
| *ARSB* | AR | AR |
| *ASL* | AR | AR |
| *ASS1* | AR | AR |
| *ATP7A* | XL | XL |
| *AVPR2* | XL | XL |
| *BCKDHA* | AR | AR |
| *BCKDHB* | AR | AR |
| *BLNK* | AR | AR |
| *BTK* | XL | XL |
| *CA5A* | AR | AR |
| *CACNA1C* | AD | AD |
| *CACNA1D* | AD | AD |
| *CAD* | AR | AR |
| *CASR* | AR | AD;AR |
| *CD247* | AR | AR |
| *CD3D* | AR | AR |
| *CD3E* | AR | AR |
| *CD40* | AR | AR |
| *CD40LG* | XL | XL |
| *CD79A* | AR | AR |
| *CFTR* | AR | AR |
| *CHAT* | AR | AR |
| *CHRNA1* | AD | AD |
| *CHRNB1* | AD | AD |
| *CHRND* | AR | AR |
| *CHRNE* | AR | AR |
| *CIITA* | AR | AR |
| *CLCN2* | AD | AD |

| | | |
|---|---|---|
| *COL13A1* | AR | AR |
| *COL1A1* | AD | AD |
| *COL1A2* | AD | AD |
| *COLQ* | AR | AR |
| *COQ2* | AR | AR |
| *COQ4* | AR | AR |
| *COQ5* | AR | AR |
| *COQ6* | AR | AR |
| *COQ7* | AR | AR |
| *COQ8A* | AR | AR |
| *COQ9* | AR | AR |
| *CORO1A* | AR | AR |
| *CPS1* | AR | AR |
| *CPT2* | AR | AD;AR |
| *CTNS* | AR | AR |
| *CYBA* | AR | AR |
| *CYBB* | XL | XL |
| *CYBC1* | AR | AR |
| *CYP11A1* | AR | AR |
| *CYP11B1* | AD | AD |
| *CYP11B2* | AR | AR |
| *CYP21A2* | AR | AR |
| *DBT* | AR | AR |
| *DCLRE1C* | AR | AR |
| *DDC* | AR | AR |
| *DHCR7* | AR | AR |
| *DLAT* | AR | AR |
| *DLD* | AR | AR |
| *DOCK8* | AR | AR |
| *DOK7* | AR | AR |
| *DPAGT1* | AR | AR |
| *ECHS1* | AR | AR |
| *EDA* | XL | XL |
| *EIF2AK3* | AR | AR |
| *ETFA* | AR | AR |
| *ETFB* | AR | AR |
| *ETFDH* | AR | AR |
| *F10* | AR | AR |
| *F11* | AR | AD;AR |
| *F12* | AR | AR |
| *F13A1* | AR | AR |
| *F13B* | AR | AR |
| *F2* | AR | AR |
| *F5* | AR | AR |
| *F7* | AR | AR |
| *F8* | XL | XL |
| *F9* | XL | XL |

| | | |
|---|---|---|
| *FAH* | AR | AR |
| *FAM111A* | AD | AD |
| *FAS* | AD | AD |
| *FASLG* | AR | AR |
| *FBP1* | AR | AR |
| *FECH* | AR | AR |
| *FGA* | AR | AR |
| *FGB* | AR | AR |
| *FGG* | AR | AR |
| *FLAD1* | AR | AR |
| *FNIP1* | AR | AR |
| *FOLR1* | AR | AR |
| *FOXN1* | AR | AR |
| *FOXP3* | XL | XL |
| *GAA* | AR | AR |
| *GALNS* | AR | AR |
| *GALT* | AR | AR |
| *GATA4* | AD | AD |
| *GATA6* | AD | AD |
| *GBA1* | AR | AR |
| *GCK* | AR | AD;AR |
| *GFPT1* | AR | AR |
| *GLDC* | AR | AR |
| *GLIS3* | AR | AR |
| *GLUD1* | AD | AD |
| *GMPPB* | AR | AR |
| *GOT2* | AR | AR |
| *GPIHBP1* | AR | AR |
| *GUCY2C* | AD | AD |
| *GUSB* | AR | AR |
| *HADHA* | AR | AR |
| *HADHB* | AR | AR |
| *HBA1* | AR | AR |
| *HBA2* | AR | AR |
| *HESX1* | AR | AD;AR |
| *HIBCH* | AR | AR |
| *HLCS* | AR | AR |
| *HMGCL* | AR | AR |
| *HNF1A* | AD | AD |
| *HNF4A* | AD | AD |
| *HSD3B2* | AR | AR |
| *IDS* | XL | XL |
| *IDUA* | AR | AR |
| *IER3IP1* | AR | AR |
| *IGHM* | AR | AR |
| *IGSF1* | XL | XL |
| *IKBKG* | XL | XL |

| | | |
|---|---|---|
| *IKZF1* | AD | AD |
| *IL10RA* | AR | AR |
| *IL10RB* | AR | AR |
| *IL1RN* | AR | AR |
| *IL2RG* | XL | XL |
| *IL7R* | AR | AR |
| *INS* | AR | AD; AR |
| *IRAK4* | AR | AR |
| *ITGB2* | AR | AR |
| *IVD* | AR | AR |
| *JAK3* | AR | AR |
| *KCNH2* | AD | AD |
| *KCNJ11* | AR | AD; AR |
| *KCNJ5* | AD | AD |
| *KCNQ1* | AD | AD |
| *KCNQ2* | AD | AD |
| *KCNQ3* | AD | AD |
| *LAT* | AR | AR |
| *LCK* | AR | AR |
| *LCP2* | AR | AR |
| *LHX3* | AR | AR |
| *LHX4* | AD | AD |
| *LIG4* | AR | AR |
| *LIPA* | AR | AR |
| *LRBA* | AR | AR |
| *LYST* | AR | AR |
| *MAGED2* | XL | XL |
| *MCEE* | AR | AR |
| *MECOM* | AD | AD |
| *MMAA* | AR | AR |
| *MMAB* | AR | AR |
| *MMACHC* | AR | AR |
| *MMADHC* | AR | AR |
| *MMUT* | AR | AR |
| *MNX1* | AR | AR |
| *MOCS1* | AR | AR |
| *MPI* | AR | AR |
| *MPL* | AR | AR |
| *MRAP* | AR | AR |
| *MSN* | XL | XL |
| *MUSK* | AR | AR |
| *MYD88* | AR | AR |
| *MYO5B* | AR | AR |
| *NAGS* | AR | AR |
| *NANS* | AR | AR |
| *NCF1* | AR | AR |
| *NCF2* | AR | AR |

| | | |
|---|---|---|
| *NCF4* | AR | AR |
| *NEUROG3* | AR | AR |
| *NHEJ1* | AR | AR |
| *NKX2-2* | AR | AR |
| *NLRC4* | AD | AD |
| *NLRP3* | AD | AD |
| *NR0B1* | XL | XL |
| *NR3C2* | AD | AD |
| *NR5A1* | AR | AR |
| *OAT* | AR | AR |
| *OTC* | XL | XL |
| *OTX2* | AD | AD |
| *OXCT1* | AR | AR |
| *PAX1* | AR | AR |
| *PAX6* | AD | AD |
| *PCCA* | AR | AR |
| *PCCB* | AR | AR |
| *PCSK1* | AR | AR |
| *PDHA1* | XL | XL |
| *PDHB* | AR | AR |
| *PDHX* | AR | AR |
| *PDP1* | AR | AR |
| *PDSS1* | AR | AR |
| *PDSS2* | AR | AR |
| *PDX1* | AD | AD |
| *PGM1* | AR | AR |
| *PGM3* | AR | AR |
| *PHGDH* | AR | AR |
| *PLEC1* | AR | AR |
| *PLPBP* | AR | AR |
| *PNPO* | AR | AR |
| *POMC* | AR | AR |
| *POU1F1* | AR | AD; AR |
| *PREPL* | AR | AR |
| *PRF1* | AR | AR |
| *PRKDC* | AR | AR |
| *PROP1* | AR | AR |
| *PRRT2* | AD | AD |
| *PTF1A* | AR | AR |
| *PTPRC* | AR | AR |
| *RAB27A* | AR | AR |
| *RAC2* | AD | AD |
| *RAG1* | AR | AR |
| *RAG2* | AR | AR |
| *RAPSN* | AR | AR |
| *RBM8A* | AR | AR |
| *RFX5* | AR | AR |

| | | |
|---|---|---|
| *RFX6* | AR | AR |
| *RFXANK* | AR | AR |
| *RFXAP* | AR | AR |
| *RMRP* | AR | AR |
| *RPL5* | AD | AD |
| *RPS10* | AD | AD |
| *RPS19* | AD | AD |
| *RPS24* | AD | AD |
| *SCN2A* | AD | AD |
| *SCN4A* | AR | AR |
| *SCN5A* | AD | AD |
| *SCN8A* | AD | AD |
| *SCNN1A* | AR | AR |
| *SCNN1B* | AR | AR |
| *SCNN1G* | AR | AR |
| *SFTPC* | AD | AD |
| *SH2D1A* | XL | XL |
| *SI* | AR | AR |
| *SLC13A5* | AR | AR |
| *SLC16A2* | XL | XL |
| *SLC18A3* | AR | AR |
| *SLC19A3* | AR | AR |
| *SLC25A1* | AR | AR |
| *SLC25A15* | AR | AR |
| *SLC25A20* | AR | AR |
| *SLC26A3* | AR | AR |
| *SLC2A1* | AR | AD; AR |
| *SLC2A2* | AR | AR |
| *SLC35A2* | XL | XL |
| *SLC5A1* | AR | AR |
| *SLC5A7* | AR | AR |
| *SLC9A3* | AR | AR |
| *SNAP25* | AD | AD |
| *SOX3* | XL | XL |
| *STAR* | AR | AR |
| *STAT3* | AD | AD |
| *STX11* | AR | AR |
| *STXBP1* | AR | AD; AR |
| *STXBP2* | AR | AR |
| *SYT2* | AD | AD |
| *TAFAZZIN* | XL | XL |
| *TBX19* | AR | AR |
| *TCF3* | AR | AD; AR |
| *THPO* | AR | AR |
| *TPO* | AR | AR |
| *TRHR* | AR | AR |
| *TRMU* | AR | AR |

| | | |
|---|---|---|
| *TRPM6* | AR | AR |
| *TSC1* | AD | AD |
| *TSC2* | AD | AD |
| *TSHB* | AR | AR |
| *TSHR* | AD | AD |
| *TTC7A* | AR | AR |
| *UNC13D* | AR | AR |
| *UROS* | AR | AR |
| *USP18* | AR | AR |
| *VAMP1* | AR | AR |
| *WAS* | XL | XL |
| *XIAP* | XL | XL |
| *ZAP70* | AR | AR |
| *ZFP57* | AR | AD; AR |

Supplemental Table 3

| Phenotype | Gene symbol | Penetrance | Variable expressivity | Age of onset restricted to infancy? | Primarily de novo | Opportunity for intervention | Established versus investigational interventions | Maternal or pregnancy implications | Include/ exclude |
|---|---|---|---|---|---|---|---|---|---|
| Long QT Syndrome | *KCNH2* | Incomplete penetrance | Yes | No | No | Prenatal & postnatal | Established and investigational | Maternal cardiovascular events | Include |
| | *KCNQ1* | Incomplete penetrance | Yes | No | No | Early postnatal | Established | Maternal cardiovascular events | Include |
| | *SCN5A* | Incomplete penetrance | Yes | No | No | Prenatal & postnatal | Established and investigational | Maternal cardiovascular events | Include |
| Combined pituitary hormone deficiency | *LHX4* | Incomplete penetrance | Yes | Not restricted to infancy in cases of growth hormone deficiency | Only ~10 cases, 4 were from a family, the others were separate /de novo | Early postnatal | Established | Fetal growth restriction - close monitoring of growth needed in pregnancy | Include |
| | *OTX2* | Incomplete penetrance | Yes | No | No | Early postnatal | Established | | Include |
| Congenital hyperinsulinism | *GCK* | Incomplete penetrance | Yes | No | No | Early postnatal | Established | GDM screening differently (FBG), can lead macrosomia--growth US, low risk of T2DM on Mothers compared to GDM | Include |
| | *GLUD1* | Incomplete penetrance | Yes | No | No | Early postnatal | Established | HI/HA, diet changes | Include |
| | *HNF4A* | Incomplete penetrance | Yes | No (onset from adolescent to early adult) | No | Early postnatal | Established | Tighter glycemic control | Include |
| | *HNF1A* | Incomplete penetrance | Yes | No (onset from adolescent to early adult) | No | Early postnatal | Established | Tighter glycemic control | Include |
| FAM111A-related skeletal dysplasia | *FAM111A* | Complete penetrance | Yes | Yes | Yes | Early postnatal | Established | Maternal calcium levels evaluation | Exclude |
| Hyperaldosteronism | *CYP11B1/CYP11 B2* | Incomplete penetrance | Yes | No | No | Early postnatal | Established | Hypertension, virilization (mother) | Include |
| | *CLCN2* | Incomplete penetrance | Yes | No | No | Early postnatal | Established | Hypertension | Include |
| | *KCNJ5* | High penetrance but can be incomplete depending on the variant for this phenotype | Yes | No | No | Early postnatal | Established | Hypertension | Include |
| Infantile Diabetes Mellitus | *KCNJ11* | Incomplete penetrance | Yes | No | Most cases | Early postnatal | Established | Glycemic control | Include |
| | *GATA6* | Incomplete penetrance | Yes | No | Yes | Early postnatal | Established | | Include |
| | *PDX1 (OMIM#: 606392)* | Incomplete penetrance depending on the variant | Yes | No | Yes | Early postnatal | Established | Increased risk of GDM | Include |
| | *GATA4* | Incomplete penetrance | Yes | No | Yes | Early postnatal | Established | | Include |
| | *PAX6* | Incomplete penetrance – high penetrance | Yes | No | No | Early postnatal | Established | | Include |
| | *STAT3* | Incomplete penetrance | Yes | No | Yes | Early postnatal | Established | | Include |
| Permanent neonatal hyperthyroidism | *TSHR* | Incomplete penetrance | Yes | No | Yes | Early postnatal | Established | Gestational hyperthyroidism | Include |
| Primary aldosteronism | *NR3C2* | Incomplete penetrance | Yes | Yes | Yes | Early postnatal | Established | Hypertension | Include |
| | *CACNA1D* | Incomplete penetrance | Yes | No | Yes - only 2 separate cases reported | Early postnatal | Established | Preeclampsia and preterm birth | Include |
| Congenital sodium diarrhea | *GUCY2C* | Incomplete penetrance | Yes | No | No | Early postnatal | Established | | Include |
| Severe congenital thrombocytopenias | *MECOM* | Incomplete penetrance | Yes | No | Yes - only 3 separate cases | Early postnatal | Established | *MECOM* -AS- bone marrow failure leading to hydrops fetalis heart failure which can lead to | Include |
| Autoinflammatory | *NLRC4* | Incomplete penetrance | Yes | Yes | Yes | Early postnatal | Established | Autoinflammation and uncontrolled immune response => PPROM | Include |

| | | | | | | | | | |
|---|---|---|---|---|---|---|---|---|---|
| disorders | *NLRP3* | Incomplete penetrance | Yes | Yes | Yes | Early postnatal | Established | Preeclampsia (overactive *NLRP3* seen in placenta), preterm birth and spontaneous abortions | Include |
| Combined immunodeficiencies | *IKZF1* | Incomplete penetrance | Yes | No | | Early postnatal | Established | Increased susceptibility to infection, preterm birth | Include |
| | *STAT3* | Incomplete penetrance | Yes | No | No | Early postnatal | Established | Increased susceptibility to infection (may require antibiotics), spontaneous abortions, preeclampsia | Include |
| Diseases of immune dysregulation | *FAS* | Incomplete penetrance | Yes | No | Yes | Early postnatal | Established | Spontaneous abortions, preeclampsia | Include |
| Severe combined immunodeficiencies | *RAC2* | Incomplete penetrance | No | No | Yes | Early postnatal | Established | Risk of infection | Include |
| Congenital myasthenia syndromes | *SYT2* | Incomplete-high penetrance | Yes | No | Yes (some) | Early postnatal | Established | If mother undiagnosed- Magnesium sulfate can trigger crisis | Include |
| | *CHRNA1* | Incomplete penetrance | Yes | No | Yes | Early postnatal | Established | If mother undiagnosed- Magnesium sulfate can trigger crisis | Include |
| | *CHRNB1* | Incomplete-high penetrance | No | No | Yes | Early postnatal | Established | If mother undiagnosed- Magnesium sulfate can trigger crisis | Include |
| | *SNAP25* | No (only 1 patient) | No | Yes | Yes | Early postnatal | Established | If mother undiagnosed- Magnesium sulfate can trigger crisis | Exclude |
| Developmental and epileptic encephalopathy | *SCN2A* | Incomplete penetrance | Yes | Yes | Yes | Early postnatal | Established | Prenatal movement changes (fetal seizures if inherited) | Include |
| Self-limited familial neonatal epilepsy, neonatal-onset developmental and epileptic encephalopathy | *KCNQ2* | Incomplete penetrance | Yes | Yes | Yes | Early postnatal | Established | | Include |
| | *SCN8A* | Complete penetrance | Yes | Yes | Yes | Prenatal & postnatal | Established and investigational | Prenatal movement changes (fetal seizures if inherited) | Exclude |
| | *PRRT2* | Incomplete penetrance | Yes | Yes | Potentially yes? | Early postnatal | Established | | Include |
| | *KCNQ3* | Complete penetrance | No | Yes | NA | Early postnatal | Established | | Exclude |
| Surfactant deficiency | *SFTPC* | Incomplete penetrance | Yes | No | | Early postnatal | Established | | Include |
| Osteogenesis Imperfecta type III or severe type IV | *COL1A1* | Incomplete penetrance | Yes | No | | Prenatal | Investigational | OI in pregnancy guidelines: PPH, fractures, GDM. C-section risk | Include |
| | *COL1A2* | Incomplete penetrance | Yes | No | | Prenatal | Investigational | Osteogenesis imperfecta in pregnancy guidelines: PPH, fractures, GDM. C-section | Include |
| Long QT syndrome | *CACNA1C* | Incomplete penetrance | Yes | No | | Prenatal | Investigational | Maternal cardiovascular events (arrhythmias) | Include |
| Diamond-Blackfan anemia | *RPS19* | High penetrance but can be incomplete | Yes | Yes | Most cases de-novo | Prenatal | Investigational | If inherited by the fetus risk of Hydrops Fetalis => mirror syndrome | Include |
| | *RPL5* | Incomplete penetrance | Yes | Yes | No | Prenatal | Investigational | If inherited by the fetus risk of Hydrops Fetalis => mirror syndrome | Include |
| | *RPS10* | Incomplete penetrance | Yes | No | No | Prenatal | Investigational | If inherited by the fetus risk of Hydrops Fetalis => mirror syndrome | Include |
| | *RPS24* | Incomplete penetrance | Yes | Yes | No | Prenatal | Investigational | If inherited by the fetus risk of Hydrops Fetalis => mirror syndrome | Include |

| | | | | | | | | | |
|---|---|---|---|---|---|---|---|---|---|
| Tuberous sclerosis complex | *TSC1* | Complete penetrance | Yes | No | | Prenatal | Investigational | Tuberous sclerosis in pregnancy: increased risk of preeclampsia, worsening existing conditions (angiomyolipoma) | Include |
| | *TSC2* | Complete penetrance | Yes | No | Yes | Prenatal | Investigational | Tuberous sclerosis in pregnancy: increased risk of preeclampsia, worsening existing conditions (angiomyolipoma) | Include |

Supplemental Table 4.

| Condition | Genes | Direct maternal impact | Fetal-mediated obstetric impact | Mechanistic evidence |
|---|---|---|---|---|
| Long QT syndrome (1-3) | *KCNH2* | Cardiac arrythmias | | *KCNH2* encodes for rapidly activating delayed-rectifier potassium channels. |
| | *KCNQ1* (4) | | | Impairment in the $I_{ks}$ (adrenergic-sensitive, slowly activating delayed rectifier potassium channels) current disrupts the heart's normal ability to shorten ventricular repolarization during increase in sympathetic activity and heart rate. |
| | *CACNA1C* (5) | | | L-type calcium channel is comprised of 3 subunits; *CACNA1C* encodes one of these sub-units (a1c alpha-subunit) so mutations in *CACNA1C* can cause alterations in this channel leading to impaired channel activation, thus QT prolongation |
| | *SCN5A* | Increased risk of adverse cardiac events during rest or sleep | | Mutation in the sodium channels, therefore impaired fast inactivation of cardiac sodium channels during early cardiac repolarization |
| Congenital hyperinsulinism (6, 7) | *GLUD1* (8) | Maternal hypoglycemia, HI/HA | | Gain of function mutation in *GLUD1* è unrestrained insulin production by beta-cells in pancreas & increased ammonia levels |
| | *HNF4A* (9) | GDM | Increased risk of infant macrosomia; implications for delivery | Loss of *HNF4A* function leads to beta-cell dysfunction and impaired insulin secretion |
| | *HNF1A* (10, 11) | | | *HNF1A* acts as a regulator of insulin secretion partially through *GLUT2*; impaired insulin secretion in beta-cells with decreased insulin response to high glucose |
| Infantile diabetes | *PDX1* (12) | GDM | | PDX1 is a nuclear transcription factor involved with pancreatic development, beta-cell differentiation, and mature beta-cell function. Activates insulin gene expression promoter, |

| | | | | |
|---|---|---|---|---|
| | | | | therefore can lead to abnormalities in blood glucose regulation. |
| Hyperaldosteronism (13) | *CYP11B1 (14)* | Pre-eclampsia, HTN | | Mutation in the *CYP11B1* gene causes 11 beta-OH deficiency in the adrenal cortex, leading to increased concentration of deoxycorticosterone, 11-deoxycortisol, and overproduction of adrenal androgens. |
| | *CLCN2* (15) | Pre-eclampsia; HTN; hypokalemia | | The *CLCN2* encodes a voltage-gated chloride channel that is expressed in the adrenal glomerulosa which opens at hyperpolarized membrane potentials. With *CLCN2* gain-of-function mutations, channels have a higher probability of opening at the glomerulosa resting potential, which means that there is higher expression of aldosterone synthase and thus hyperaldosteronism. |
| | *KCNJ5* (16) | Pre-eclampsia | | *KCNJ5* encodes the G-protein-gated inwardly rectifying $K^+$ channel Kir3.4/GIRK4 in zona glomerulosa cells. The hot-spot mutations (G151R, L168R in APAs; T158A in FH-III) cluster in or near the selectivity filter, causing loss of $K^+$ selectivity and pathological $Na^+$ conductance → chronic membrane depolarization → opening of voltage-gated $Ca^{2+}$ channels → increased intracellular $Ca^{2+}$ → constitutive aldosterone synthesis and possibly cell proliferation |
| Pseudo-hypoaldosteronism | *NR3C2* (17) | HTN | | The *NR3C2* gene encodes the mineralocorticoid receptor. Variants of *NR3C2* can alter receptor distribution or expression, so can affect renal tubular sensitivity to aldosterone. |
| Primary aldosteronism | *CACNA1D* (18) | Pre-eclampsia | | *CACNA1D* is involved in encoding voltage-gated calcium channel resulting in channel activation at less depolarized potentials; Gly403 alterations also impair channel inactivation. |

| | | | | |
|---|---|---|---|---|
| | | | | These effects are inferred to cause increased Ca(2+) influx, which is a sufficient stimulus for aldosterone production and cell proliferation in adrenal glomerulosa. |
| Tuberous sclerosis complex (19, 20) | *TSC1*<br>*TSC2* | Lymphangio-leiomyomatosis (LAM) exacerbation, seizures | | *TSC1* and *TSC2* encode hamartin and tuberin, respectively, which form components of a GTPase activating protein for Rheb, which normally suppresses mTOR. Mutations in *TSC1* and *TSC2* therefore can cause hyperactivation of mTOR. LAM exacerbation can worsen during pregnancy, likely due to estrogen-mediated augmentation of mTOR-dependent pathways. mTOR hyperactivation in vascular endothelial cells can lead to disorganized vascular networks, which can explain the preeclampsia, FGR, and placental abruption in TSC pregnancies |
| Immune dysregulation | *FAS (*21-23) | Spontaneous abortion; preeclampsia risk | | Fas-Fas-L system is an important inducer of apoptosis. Therefore, mutations affecting this system could lead to abnormal trophoblastic development and invasion. |
| Autoinflammatory disorders | *NLRC4* (24) | PPROM | | *NLRC4* encodes the pattern recognition receptor in the inflammasome complex that is mainly expressed in macrophages, neutrophils, dendritic cells, and glial cells. Mechanisms responsible for PPROM are thought to involve inflammation. |
| | *NLRP3* (25, 26) | Pre-eclampsia, HTN | | *NLPR3* makes up an inflammasome (consisting of NLPR4, ASC, and cysteine protease precursor procaspase-1). Activation of NLRP3 inflammasome induces inflammation in the hypothalamus, inducing a sympathetic response that results in RAAS activation. The NLRP3 inflammasome can further be activated by |

| | | | | |
|---|---|---|---|---|
| | | | | DAMPs and PAMPs, causing activation of platelets and leukocytes downstream, thus playing a role in pre-eclampsia associated coagulopathy. |
| Combined immunodeficiency | *IKZF1* (27, 28) | Risk of infection | | The *IKZF1* gene encodes a member of the IKAROS family of the zinc transcription factor. This has a role in lymphocyte differentiation and development and also plays a role in the ontogeny of B cells. Mutations in this gene therefore can result in inborn error of immunity. The clinical phenotype of IKZF1 mutations is primary immunodeficiency with b-cell deficiency, hypogammaglobulinemia, and autoimmunity which can theoretically affect pregnancy. Plausible that it could cause increased risk of infection due to poor immune system. |
| | *STAT3* (29-32) | Risk of infection, spontaneous abortions, pre-eclampsia | | *STAT3* is a signal transducer protein; it serves an important role in regulating immune response by modulating Th1/Th17/Th2 and Treg gene expression. STAT3 is required for embryonic development, as deficient embryos die between 6.5-7.5 dpc. Hypothesized that *STAT3* has a PR-dependent role; STAT3 and PR crosstalk needed for successful implantation in uterus.<br>There is also evidence of the role of the *STAT3*/EMT pathway in mediating the effect of RPS4Y1 on trophoblast cell migration/invasion. |
| Severe Combined Immunodeficiency | *RAC2* (33, 34) | Risk of infection | | *RAC2* is confined to the hematopoietic system; *in vivo* studies have found *RAC2* coordinates distinct cellular processes in cells of the hematopoietic system. It is a GTPase that acts as a regulator of cell-behavior and plays a key role in F-actin assembly, chemotaxis, and superoxide production. Therefore, in gain of |

| | | | | |
|---|---|---|---|---|
| | | | | function and loss of function mutations, these processes are impaired. Gain of function *RAC2* mutations also appear to drive *NLRP3* dependent inflammasome activation leading to hyperinflammatory features. |
| Neonatal hyperthyroidism (35) | *TSHR* (36-39) | Gestational hyperthyroidism; spontaneous abortion; hyperemesis gravidarum | Non-immune fetal hydrops | *TSHR* mutation can lead to a TSH receptor with an increased sensitivity to hCG. In patients with this gain of function mutation, normal hCG levels can lead to severe, early, and prolonged gestational thyrotoxicosis with suppressed TSH, and absence of thyroid antibody. |
| Severe congenital thrombocytopenia | *MECOM (40, 41)* | | Fetal bone marrow failure è hydrops fetalis è maternal mirror syndrome; intraventricular hemorrhage can impact delivery mode | The *MEOCM* gene encodes many protein isoforms that regulate hematopoietic stem cell self-renewal and maintenance. MECOM also encodes a zinc finger transcription factor. |
| Diamond-Blackfan Anemia (42-44) | *RPS19 (45)* | Recurrent spontaneous abortions | Hydrops fetalis è maternal mirror syndrome | *RPS19* haploinsufficiency gives rise to hematopoietic stem and progenitor cell depletion and impaired erythroid expansion through P53-dependent apoptosis. |
| | *RPL5* | | | *RPL5* haploinsufficiency has a distinct and more severe prenatal consequence than *RPS19*. In in vivo models, *RPL5* haploinsufficiency causes hematopoietic stem/progenitor accumulation and prenatal lethality via p53-mediated ferroptosis of mature erythroid progenitors (nature article linked above) |
| | *RPS10* | | | |
| | *RPS24 (46)* | | | *RPS24* haploinsufficiency impairs erythropoiesis via a STAT6–SATB1 pathway, reducing hemoglobin—consistent with an erythroid-specific defect |

| | | | | |
|---|---|---|---|---|
| Congenital Myasthenic Syndromes (47-49) | *SYT2*<br>*CHRNA1*<br>*CHRNB1* | Magnesium sulfate (seizure prophylaxis in the setting of pre-eclampsia) can trigger myasthenic crisis (i.e., bulbar weakness, limb weakness) | | Minimal mechanistic evidence |
| Epileptic encephalopathy (50-52) | *SCN2A* | Mother at risk of seizures; pregnancy can lower seizure threshold increasing risk | | *SCN2A* is located on the transmembrane S5 of DI of NaV1.2, which is an evolutionarily conserved region and is the principal VGSC in excitatory pyramidal neurons, mainly expressed in axons and nerve terminals. Therefore, a variant in this domain can have detrimental effects on the channel function, stability, and trafficking, resulting in severe phenotypes and/or lethality. |
| Osteogenesis imperfecta (53-56) | *COL1A1*<br>*COL1A2* | Post-partum hemorrhage; GDM | Risk of fetal fractures during delivery | Collagen defects with these mutations will affect bone, vascular structures, fetal membrane structures, and cervix, thus impacting fracture risk, breastfeeding, bleeding, and preterm birth. |

#### Abbreviations

| *Acronym* | Definition |
|---|---|
| ***11β-OH*** | 11-beta-hydroxylase |
| ***APA(s)*** | aldosterone-producing adenoma(s) |
| ***DAMPs*** | Damage-associated molecular patterns |
| ***DI*** | domain I |
| ***DM*** | diabetes mellitus |
| ***dpc*** | days post conception |
| ***EMT*** | epithelial–mesenchymal transition |
| ***F-actin*** | filamentous actin |
| ***Fas-L*** | Fas ligand |
| ***FGR*** | fetal growth restriction |
| ***FH-III*** | familial hyperaldosteronism type III |
| ***GDH*** | glutamate dehydrogenase |
| ***GIRK4*** | G-protein-gated inwardly rectifying potassium channel 4 |
| ***GLUT2*** | glucose transporter 2 |
| ***GoF*** | gain of function |
| ***GTP*** | guanosine triphosphate |
| ***GTPase*** | guanosine triphosphatase |
| ***hCG*** | human chorionic gonadotropin |
| ***HI/HA*** | hyperinsulinism/hyperammonemia syndrome |
| ***HTN*** | hypertension |
| $I_{ks}$ | slowly activating delayed-rectifier potassium current |
| ***LAM*** | lymphangioleiomyomatosis |
| ***LoF*** | loss of function |
| ***LQT/LQT3*** | long QT syndrome/long QT syndrome type 3 |
| ***MODY*** | maturity-onset diabetes of the young |
| ***mTOR*** | mechanistic target of rapamycin |
| ***NaV1.2*** | voltage-gated sodium channel NaV1.2 |
| ***p53/P53*** | tumor protein p53 |
| ***PAMPs*** | Pathogen-associated molecular patterns |
| ***PPROM*** | preterm prelabor rupture of membranes |
| ***PR*** | progesterone receptor |

| | |
|---|---|
| ***QT*** | QT interval (the interval from the start of the Q wave to the end of the T wave on an electrocardiogram) |
| ***Rheb*** | Ras homolog enriched in brain |
| ***S5*** | fifth transmembrane segment |
| ***Th1*** | T-helper type 1 cell |
| ***Th2*** | T-helper type 2 cell |
| ***Th17*** | T-helper type 17 cell |
| ***Treg*** | regulatory T cell |
| ***TSC*** | tuberous sclerosis complex |
| ***TSH*** | thyroid-stimulating hormone |
| ***VGSC*** | voltage-gated sodium channel |

### Supplemental Table 5

| Panel | Description | Genes (N) | Includes | Excludes |
|---|---|---|---|---|
| **Panel 1: 293 gene list** | All perinatal treatable genes | 293 | AR (222), XL (25), AD (46) | None |
| **Panel 2: AR + XL** | Autosomal recessive and X-linked genes | 247 | AR (222), XL (25) | AD genes |
| **Panel 3: 46 AD gene list** | Autosomal dominant genes | 46 | AD (46) | AR/XL genes |
| **Panel 4: 289 gene list** | All genes except 4 excluded AD | 289 | AR (222), XL (25), AD (42) | *FAM111A, SNAP25, SCN8A, KCNQ3* |
| **Panel 5: 42 AD gene list** | AD genes except 4 excluded | 42 | AD (42) | *FAM111A, SNAP25, SCN8A, KCNQ3* , AR/XL genes |

Supplemental Table 6

| Gene symbol | Inheritance | Primary mechanism of pathogenicity |
|---|---|---|
| *GLUD1* | AD | Gain of function |
| *FAM111A* | AD | Gain of function |
| *CYP11B1* | AD | Gain of function |
| *CLCN2* | AD | Gain of function |
| *KCNJ5* | AD | Gain of function |
| *KCNJ11* | AD | Gain of function |
| *STAT3* | AD | Gain of function |
| *TSHR* | AD | Gain of function |
| *CACNA1D* | AD | Gain of function |
| *GUCY2C* | AD | Gain of function |
| *NLRC4* | AD | Gain of function |
| *NLRP3* | AD | Gain of function |
| *RAC2* | AD | Gain of function |
| *CHRNB1* | AD | Gain of function |
| *CACNA1C* | AD | Gain of function |

### Supplemental Table 7

| Top 20 genes by carrier frequency (gnomAD) | ACMG T3 | Schmitz (2024) | Hotakainen (2025) | Ba | In | FuP | FuT3 | My | Qu | La | Bl | Na |
|---|---|---|---|---|---|---|---|---|---|---|---|---|
| *CFTR* (AR) | • | • | • | • | • | • | • | • | • | • | • | • |
| *CYP21A2* (AR) | • | • | • | • | | • | • | • | • | • | • | • |
| *F2* (AD;AR) | | | | • | | | • | • | | | | |
| *DHCR7* (AR) | • | • | • | • | | • | • | • | • | • | • | • |
| *GAA (AR)* | • | • | • | • | • | • | • | • | • | • | • | • |
| *ACADM* (AR) | • | • | • | • | • | • | • | • | • | • | • | • |
| *ALDOB* (AR) | • | • | • | • | | • | • | • | • | • | • | • |
| *RBM8A* (AR) | | • | | | | | | | | | | |
| *ASL (AR)* | • | • | • | • | • | • | • | • | • | • | • | • |
| *GBA1* (AR) | • | • | • | • | | • | • | • | • | • | • | • |
| *ALPL* (AR) | • | • | • | • | | • | • | • | • | • | • | • |
| *ACADVL* (AR) | • | • | • | • | • | • | • | • | • | • | • | • |
| *RMRP* (AR) | | • | • | • | | • | • | • | • | • | • | • |
| *CPT2* (AR) | • | • | • | • | • | • | • | • | • | • | • | • |
| *IDUA* (AR) | • | | • | • | • | • | • | • | • | • | • | • |
| *RAPSN* (AR) | | | • | • | | • | • | • | • | • | • | • |
| *PRRT2* (AD) | | | | | | | | | | | | |
| *MMACHC* (AR) | • | • | • | • | • | • | • | • | • | • | • | • |
| *F11* (AD;AR) | | | | • | | • | • | • | • | | | • |
| *AGXT* (AR) | • | • | • | • | | • | • | • | • | • | • | • |

### Supplemental Table 8

| Top 20 genes by carrier frequency (AoU) | ACMG T3 | Schmitz 2024 | Hotakainen 2025 | Ba | In | FuP | FuT3 | My | Qu | La | Bl | Na |
|---|---|---|---|---|---|---|---|---|---|---|---|---|
| *CFTR* (AR) | ● | ● | ● | ● | ● | ● | ● | ● | ● | ● | ● | ● |
| *F2* (AD;AR) | | | | ● | | | ● | ● | | | | |
| *DHCR7* (AR) | ● | ● | ● | ● | | ● | ● | ● | ● | ● | ● | ● |
| *GAA* (AR) | ● | ● | ● | ● | ● | ● | ● | ● | ● | ● | ● | ● |
| *ACADM* (AR) | ● | ● | ● | ● | ● | ● | ● | ● | ● | ● | ● | ● |
| *ALDOB* (AR) | ● | ● | ● | ● | | ● | ● | ● | ● | ● | ● | ● |
| *RBM8A* (AR) | | ● | | | | | | | | | | |
| *GBA1* (AR) | ● | ● | ● | ● | | ● | ● | ● | ● | ● | ● | ● |
| *MMACHC* (AR) | ● | ● | ● | ● | ● | ● | ● | ● | ● | ● | ● | ● |
| *F11* (AD;AR) | | | | ● | | ● | ● | ● | ● | | | ● |
| *CPT2* (AR) | ● | ● | ● | ● | ● | ● | ● | ● | ● | ● | ● | ● |
| *ASL* (AR) | ● | ● | ● | ● | ● | ● | ● | ● | ● | ● | ● | ● |
| *ACADVL* (AR) | ● | ● | ● | ● | ● | ● | ● | ● | ● | ● | ● | ● |
| *F7* (AR) | | | ● | | | | ● | | | | | |
| *IDUA* (AR) | ● | | ● | ● | ● | ● | ● | ● | ● | ● | ● | ● |
| *RMRP* (AR) | | ● | ● | ● | | ● | ● | ● | ● | ● | ● | ● |
| *ALPL* (AR) | ● | ● | ● | ● | | ● | ● | ● | ● | ● | ● | ● |
| *RAPSN* (AR) | | | ● | ● | | ● | ● | ● | ● | ● | ● | ● |
| *GALT* (AR) | ● | ● | ● | ● | ● | ● | ● | ● | ● | ● | ● | ● |
| *MMUT* (AR) | ● | ● | ● | ● | | | | ● | ● | ● | | |

Supplemental Table 9

| Gene | Inheritance | HI=3 | TS=3 | Reason |
|---|---|---|---|---|
| *ATP7A* | XL | Yes | No | ClinGen_HI |
| *AVPR2* | XL | Yes | No | ClinGen_HI |
| *BTK* | XL | Yes | No | ClinGen_HI |
| *CD40LG* | XL | Yes | No | ClinGen_HI |
| *COL1A1* | AD | Yes | No | ClinGen_HI |
| *CYBB* | XL | Yes | No | ClinGen_HI |
| *EDA* | XL | Yes | No | ClinGen_HI |
| *F8* | XL | Yes | No | ClinGen_HI |
| *F9* | XL | Yes | No | ClinGen_HI |
| *GATA4* | AD | Yes | No | ClinGen_HI |
| *GATA6* | AD | Yes | No | ClinGen_HI |
| *GCK* | AR | Yes | No | ClinGen_HI |
| *HNF1A* | AD | Yes | No | ClinGen_HI |
| *HNF4A* | AD | Yes | No | ClinGen_HI |
| *IDS* | XL | Yes | No | ClinGen_HI |
| *IKBKG* | XL | Yes | No | ClinGen_HI |
| *IKZF1* | AD | Yes | No | ClinGen_HI |
| *KCNH2* | AD | Yes | No | ClinGen_HI |
| *KCNQ1* | AD | Yes | No | ClinGen_HI |
| *KCNQ2* | AD | Yes | No | ClinGen_HI |
| *MNX1* | AR | Yes | No | ClinGen_HI |
| *NR0B1* | XL | Yes | No | ClinGen_HI |
| *NR3C2* | AD | Yes | No | ClinGen_HI |
| *NR5A1* | AR | Yes | No | ClinGen_HI |
| *OTC* | XL | Yes | No | ClinGen_HI |
| *OTX2* | AD | Yes | No | ClinGen_HI |
| *PAX6* | AD | Yes | No | ClinGen_HI |
| *PDHA1* | XL | Yes | No | ClinGen_HI |
| *PRRT2* | AD | Yes | No | ClinGen_HI |
| *RPS19* | AD | Yes | No | ClinGen_HI |
| *RPS24* | AD | Yes | No | ClinGen_HI |
| *SCN2A* | AD | Yes | No | ClinGen_HI |

| | | | | |
|---|---|---|---|---|
| *SCN5A* | AD | Yes | No | ClinGen_HI |
| *SH2D1A* | XL | Yes | No | ClinGen_HI |
| *SLC16A2* | XL | Yes | No | ClinGen_HI |
| *SLC2A1* | AR | Yes | No | ClinGen_HI |
| *SLC35A2* | XL | Yes | No | ClinGen_HI |
| *STXBP1* | AR | Yes | No | ClinGen_HI |
| *TSC1* | AD | Yes | No | ClinGen_HI |
| *TSC2* | AD | Yes | No | ClinGen_HI |
| *WAS* | XL | Yes | No | ClinGen_HI |
| *XIAP* | XL | Yes | No | ClinGen_HI |

### Supplemental Table 10

| Geenes with prenatal intervention | gnomAD Carrier Frequency (%) | AoU Carrier Frequency (%) | Difference (%) | AoU/gnomAD CF Ratio |
|---|---|---|---|---|
| *COL1A1* | 0.0167 | 0.0094 | -0.0073 | 0.563 |
| *COL1A2* | 0.0373 | 0.0253 | -0.012 | 0.679 |
| *GAA* | 1.6136 | 1.4356 | -0.178 | 0.89 |
| *LIPA* | 0.2795 | 0.2499 | -0.0296 | 0.894 |
| *IDUA* | 0.5343 | 0.4319 | -0.1025 | 0.808 |
| *IDS* | 0.003 | 0.0016 | -0.0014 | 0.537 |
| *GALNS* | 0.1942 | 0.2012 | 0.007 | 1.036 |
| *ARSB* | 0.129 | 0.1405 | 0.0115 | 1.089 |
| *GUSB* | 0.0502 | 0.0723 | 0.0221 | 1.442 |
| *GBA1* | 0.6388 | 0.783 | 0.1442 | 1.226 |
| *EDA* | 0.0156 | 0.0188 | 0.0032 | 1.206 |
| *KCNH2* | 0.0295 | 0.0214 | -0.0081 | 0.727 |
| *SCN5A* | 0.057 | 0.061 | 0.004 | 1.07 |
| *CACNA1C* | 0.0025 | 0.001 | -0.0016 | 0.381 |
| *TPO* | 0.3282 | 0.279 | -0.0491 | 0.85 |
| *SLC16A2* | no qualifying variants | 0.0004 | N/A | N/A |
| *RPS19* | no qualifying variants | 0.0002 | N/A | N/A |
| *RPL5* | no qualifying variants | no qualifying variants | N/A | N/A |
| *RPS10* | no qualifying variants | no qualifying variants | N/A | N/A |
| *RPS24* | no qualifying variants | no qualifying variants | N/A | N/A |
| *FGB* | 0.0162 | 0.013 | -0.0032 | 0.802 |
| *FGA* | 0.0594 | 0.0615 | 0.0021 | 1.035 |
| *FGG* | 0.0022 | 0.0034 | 0.0011 | 1.512 |
| *F2* | 2.1003 | 2.239 | 0.1387 | 1.066 |
| *F5* | 0.0468 | 0.0606 | 0.0137 | 1.293 |
| *F7* | 0.2415 | 0.4474 | 0.2058 | 1.852 |
| *F8* | 0.1146 | 0.0975 | -0.0171 | 0.851 |
| *F9* | 0.0109 | 0.0184 | 0.0075 | 1.695 |
| *F10* | 0.0042 | 0.0089 | 0.0047 | 2.114 |
| *F11* | 0.4126 | 0.5732 | 0.1606 | 1.389 |
| *F12* | 0.0049 | 0.0077 | 0.0028 | 1.583 |
| *F13A1* | 0.0466 | 0.0252 | -0.0215 | 0.54 |

| | | | | |
|---|---|---|---|---|
| *F13B* | 0.0007 | 0.0017 | 0.0009 | 2.268 |
| *HBA1* | 0.0607 | 0.0655 | 0.0048 | 1.079 |
| *HBA2* | 0.0877 | 0.0344 | -0.0533 | 0.393 |
| *IL2RG* | 0.0002 | 0.0004 | 0.0002 | 1.62 |
| *MMACHC* | 0.4529 | 0.5956 | 0.1428 | 1.315 |
| *HLCS* | 0.1023 | 0.0832 | -0.0191 | 0.813 |
| *MMAA* | 0.1385 | 0.1111 | -0.0274 | 0.802 |
| *MMBB* | 0.1486 | 0.1033 | -0.0453 | 0.695 |
| *MMADHC* | 0.0195 | 0.0147 | -0.0048 | 0.755 |
| *NANS* | 0.0016 | 0.0012 | -0.0004 | 0.748 |
| *PHGDH* | 0.0347 | 0.0386 | 0.0039 | 1.111 |
| *DHCR7* | 1.9738 | 1.5405 | -0.4333 | 0.78 |
| *TRMU* | 0.0979 | 0.0757 | -0.0222 | 0.773 |
| *ALDH7A1* | 0.2407 | 0.1684 | -0.0723 | 0.7 |
| *SCN8A* | 0.0009 | 0.0029 | 0.002 | 3.309 |
| *TSC1* | 0.002 | 0.0022 | 0.0001 | 1.061 |
| *TSC2* | 0.0046 | 0.0029 | -0.0017 | 0.626 |
| *MAGED2* | no qualifying variants | no qualifying variants | N/A | N/A |
| *ACE* | 0.0991 | 0.0721 | -0.027 | 0.727 |
| *CFTR* | 5.6373 | 5.2362 | -0.4011 | 0.929 |

### Supplemental Table 11

| Gene with carrier frequency>1/200 | ACMG T3 | Schmitz 2024 | Hotakainen 2025 | Ba | In | FuP | FuT3 | My | Qu | La | Bl | Na |
|---|---|---|---|---|---|---|---|---|---|---|---|---|
| *CFTR* (AR) | ● | ● | ● | ● | ● | ● | ● | ● | ● | ● | ● | ● |
| *F2* (AD;AR) | | | | ● | | | ● | ● | | | | |
| DHCR7 (AR) | ● | ● | ● | ● | | ● | ● | ● | ● | ● | ● | ● |
| *GAA* (AR) | ● | ● | ● | ● | ● | ● | ● | ● | ● | ● | ● | ● |
| *ACADM* (AR) | ● | ● | ● | ● | ● | ● | ● | ● | ● | ● | ● | ● |
| *ALDOB* (AR) | ● | ● | ● | ● | | ● | ● | ● | ● | ● | ● | ● |
| *RBM8A* (AR) | | ● | | | | | | | | | | |
| *GBA1* (AR) | ● | ● | ● | ● | | ● | ● | ● | ● | ● | ● | ● |
| *ASL (AR)* | ● | ● | ● | ● | ● | ● | ● | ● | ● | ● | ● | ● |
| *ACADVL* (AR) | ● | ● | ● | ● | ● | ● | ● | ● | ● | ● | ● | ● |
| *CPT2* (AR) | ● | ● | ● | ● | ● | ● | ● | ● | ● | ● | ● | ● |

### Supplemental Table 12

| Gene with carrier frequency>1/200 | **AoU** | **gnomAD** |
|---|---|---|
| *CFTR* | T | T |
| *CYP21A2* | F | T |
| *F2* | T | T |
| *DHCR7* | T | T |
| *GAA* | T | T |
| *ACADM* | T | T |
| *ALDOB* | T | T |
| *RBM8A* | T | T |
| *GBA1* | T | T |
| *ALPL* | F | T |
| *ASL* | T | T |
| *MMACHC* | T | F |
| *RMRP* | F | T |
| *F11* | T | F |
| *ACADVL* | T | T |
| *CPT2* | T | T |
| *IDUA* | F | T |

### Supplemental Table 13

| Scenario | Dataset | Panel | Ancestry | Genes in panel (*n*) | Genes with data (*n*) | Combined cf | NNS | NNS (95 CI lower) | NNS (95 CI upper) | 1 inN |
|---|---|---|---|---|---|---|---|---|---|---|
| AoU_PLP_only | All of Us | 1 – 293 genes | ALL | 293 | 242 | 0.27596707392408600 | 3.623620694239350 | 3.6069153950371200 | 3.640150920879620 | 1 in 4 |
| AoU_PLP_only | All of Us | 1 – 293 genes | AFR | 293 | 204 | 0.17751088172163200 | 5.633457455121960 | 5.553281232495210 | 5.715070904076070 | 1 in 6 |
| AoU_PLP_only | All of Us | 1 – 293 genes | AMR | 293 | 208 | 0.2302378749703700 | 4.343334041493790 | 4.289672950946430 | 4.398480528082550 | 1 in 4 |
| AoU_PLP_only | All of Us | 1 – 293 genes | EAS | 293 | 143 | 0.18133593314289500 | 5.514626818127570 | 5.275801314280500 | 5.723245765835590 | 1 in 6 |
| AoU_PLP_only | All of Us | 1 – 293 genes | EUR | 293 | 234 | 0.3243797232371940 | 3.082806748894030 | 3.065738630557740 | 3.09972671938792 | 1 in 3 |
| AoU_PLP_only | All of Us | 1 – 293 genes | MID | 293 | 62 | 0.26019537498720000 | 3.8432658537808100 | 3.3836990032443300 | 4.21874133943299 | 1 in 4 |
| AoU_PLP_only | All of Us | 1 – 293 genes | OTH | 293 | 197 | 0.30100188150762300 | 3.3222383693793400 | 3.2639739031274900 | 3.3772180716991200 | 1 in 3 |
| AoU_PLP_only | All of Us | 1 – 293 genes | SAS | 293 | 126 | 0.1669228689218260 | 5.990790875205490 | 5.536552248549140 | 6.339734680326440 | 1 in 6 |
| AoU_PLP_only | All of Us | 2 – AR+XL | ALL | 247 | 212 | 0.2709359784031370 | 3.690908848259560 | 3.673539229189890 | 3.707582720780190 | 1 in 4 |
| AoU_PLP_only | All of Us | 2 – AR+XL | AFR | 247 | 184 | 0.17151362550857800 | 5.830440567242200 | 5.74333249484064 | 5.9176206964295700 | 1 in 6 |
| AoU_PLP_only | All of Us | 2 – AR+XL | AMR | 247 | 190 | 0.22616056029990200 | 4.421637436138030 | 4.3653218306014300 | 4.476994884067060 | 1 in 4 |
| AoU_PLP_only | All of Us | 2 – AR+XL | EAS | 247 | 133 | 0.17452860288518600 | 5.72971984802889 | 5.476954797376480 | 5.962423510929930 | 1 in 6 |
| AoU_PLP_only | All of Us | 2 – AR+XL | EUR | 247 | 205 | 0.3194144568418860 | 3.1307286773654400 | 3.112960605146810 | 3.1476689924594600 | 1 in 3 |
| AoU_PLP_only | All of Us | 2 – AR+XL | MID | 247 | 61 | 0.2583664417495090 | 3.870471696047580 | 3.4046844695733900 | 4.243787302271190 | 1 in 4 |
| AoU_PLP_only | All of Us | 2 – AR+XL | OTH | 247 | 178 | 0.29623490298498600 | 3.375699453115020 | 3.3167988209132700 | 3.4326765581476600 | 1 in 3 |
| AoU_PLP_only | All of Us | 2 – AR+XL | SAS | 247 | 117 | 0.16195713121632500 | 6.1744734084250100 | 5.710144606121830 | 6.548881455169360 | 1 in 6 |
| AoU_PLP_only | All of Us | 3 – 46 AD | ALL | 46 | 30 | 0.006900759565572790 | 144.91158407965700 | 139.90220818226900 | 150.48789622545200 | 1 in 145 |
| AoU_PLP_only | All of Us | 3 – 46 AD | AFR | 46 | 20 | 0.007238810917965970 | 138.1442354735510 | 127.35075993891200 | 150.3047934429670 | 1 in 138 |

| | | | | | | | | | | |
|---|---|---|---|---|---|---|---|---|---|---|
| AoU_PLP_only | All of Us | 3 – 46 AD | AMR | 46 | 18 | 0.005268941412508890 | 189.79144418382000 | 172.11001340956000 | 210.52974209934200 | 1 in 190 |
| AoU_PLP_only | All of Us | 3 – 46 AD | EAS | 46 | 10 | 0.008246597376363800 | 121.26213447332600 | 97.76483901098650 | 155.28430859064000 | 1 in 121 |
| AoU_PLP_only | All of Us | 3 – 46 AD | EUR | 46 | 29 | 0.007295580173900420 | 137.0692907436550 | 130.44499293190000 | 143.94234022133400 | 1 in 137 |
| AoU_PLP_only | All of Us | 3 – 46 AD | MID | 46 | 1 | 0.0024660874866636600 | 405.5006180469650 | 149.58589079102400 | 4635.443904233590 | 1 in 406 |
| AoU_PLP_only | All of Us | 3 – 46 AD | OTH | 46 | 19 | 0.006773536429777180 | 147.63336853167200 | 127.46701257343600 | 173.11477990135100 | 1 in 148 |
| AoU_PLP_only | All of Us | 3 – 46 AD | SAS | 46 | 9 | 0.005925398199148390 | 168.76502918297100 | 114.36458775076200 | 273.689645291734 | 1 in 169 |
| AoU_PLP_only | All of Us | 4 – 289 genes | ALL | 289 | 240 | 0.2759374002663440 | 3.624010369869280 | 3.6071297266562100 | 3.640459225726180 | 1 in 4 |
| AoU_PLP_only | All of Us | 4 – 289 genes | AFR | 289 | 202 | 0.1774696645356240 | 5.634765821058200 | 5.553835914713570 | 5.715052757017040 | 1 in 6 |
| AoU_PLP_only | All of Us | 4 – 289 genes | AMR | 289 | 207 | 0.23022716168399400 | 4.343536152231180 | 4.289123654320930 | 4.3971405914184700 | 1 in 4 |
| AoU_PLP_only | All of Us | 4 – 289 genes | EAS | 289 | 143 | 0.18133593314289500 | 5.514626818127570 | 5.274659379266980 | 5.731315549729920 | 1 in 6 |
| AoU_PLP_only | All of Us | 4 – 289 genes | EUR | 289 | 232 | 0.3243464459258880 | 3.083123038840070 | 3.0657041132627400 | 3.099520589261120 | 1 in 3 |
| AoU_PLP_only | All of Us | 4 – 289 genes | MID | 289 | 62 | 0.26019537498720000 | 3.8432658537808100 | 3.386284340120350 | 4.21896496548743 | 1 in 4 |
| AoU_PLP_only | All of Us | 4 – 289 genes | OTH | 289 | 196 | 0.30097447357858500 | 3.322540905579160 | 3.265554768352870 | 3.3792730149547900 | 1 in 3 |
| AoU_PLP_only | All of Us | 4 – 289 genes | SAS | 289 | 126 | 0.1669228689218260 | 5.990790875205490 | 5.536593377420840 | 6.350871017457410 | 1 in 6 |
| AoU_PLP_only | All of Us | 5 – 42 AD | ALL | 42 | 28 | 0.00686005853402516 | 145.77135093528800 | 140.54213870853900 | 151.42133105825600 | 1 in 146 |
| AoU_PLP_only | All of Us | 5 – 42 AD | AFR | 42 | 18 | 0.007189060931391270 | 139.1002259604540 | 128.37287104501500 | 151.3772410325550 | 1 in 139 |
| AoU_PLP_only | All of Us | 5 – 42 AD | AMR | 42 | 17 | 0.005255097085343660 | 190.29144157754500 | 172.18821971736300 | 211.84972121897700 | 1 in 190 |
| AoU_PLP_only | All of Us | 5 – 42 AD | EAS | 42 | 10 | 0.008246597376363800 | 121.26213447332600 | 98.02680846848530 | 155.78145560750400 | 1 in 121 |
| AoU_PLP_only | All of Us | 5 – 42 AD | EUR | 42 | 27 | 0.00724668505463022 | 137.9941300693150 | 131.5548132848460 | 145.06639252168300 | 1 in 138 |
| | | | | | | 0.0024660874 | 405.50061804 | 150.42304264 | 4828.4564264 | |

| | | | | | | | | | | |
|---|---|---|---|---|---|---|---|---|---|---|
| AoU_PLP_only | All of Us | 5 – 42 AD | MID | 42 | 1 | 866636600 | 69650 | 504500 | 54650 | 1 in 406 |
| AoU_PLP_only | All of Us | 5 – 42 AD | OTH | 42 | 18 | 0.006734591717749710 | 148.4871009127990 | 127.61872922275900 | 173.817932728125 | 1 in 148 |
| AoU_PLP_only | All of Us | 5 – 42 AD | SAS | 42 | 9 | 0.005925398199148390 | 168.76502918297100 | 113.40721616433200 | 278.4736236229060 | 1 in 169 |
| AoU_PLP_plus_LoF | All of Us | 1 – 293 genes | ALL | 293 | 286 | 0.32126392237777600 | 3.1127055680534700 | 3.0979329499613100 | 3.1276577497397300 | 1 in 3 |
| AoU_PLP_plus_LoF | All of Us | 1 – 293 genes | AFR | 293 | 272 | 0.25159679675504800 | 3.9746134008756500 | 3.9177232233162700 | 4.027309088398250 | 1 in 4 |
| AoU_PLP_plus_LoF | All of Us | 1 – 293 genes | AMR | 293 | 274 | 0.27155119321566700 | 3.6825468824428800 | 3.630581587740320 | 3.730227407625180 | 1 in 4 |
| AoU_PLP_plus_LoF | All of Us | 1 – 293 genes | EAS | 293 | 216 | 0.24429144689402100 | 4.093471190720090 | 3.886776921762400 | 4.232369169084160 | 1 in 4 |
| AoU_PLP_plus_LoF | All of Us | 1 – 293 genes | EUR | 293 | 280 | 0.36155294934547800 | 2.7658466119839700 | 2.749986854942700 | 2.7814335093518100 | 1 in 3 |
| AoU_PLP_plus_LoF | All of Us | 1 – 293 genes | MID | 293 | 86 | 0.29629297672797300 | 3.375037812381570 | 2.7748691961260200 | 3.544465330732650 | 1 in 3 |
| AoU_PLP_plus_LoF | All of Us | 1 – 293 genes | OTH | 293 | 259 | 0.3406940907887920 | 2.9351844573668700 | 2.878725639538310 | 2.9855523858102500 | 1 in 3 |
| AoU_PLP_plus_LoF | All of Us | 1 – 293 genes | SAS | 293 | 189 | 0.2420661052259800 | 4.131102944241020 | 3.722769465477410 | 4.270901316620710 | 1 in 4 |
| AoU_PLP_plus_LoF | All of Us | 2 – AR+XL | ALL | 247 | 246 | 0.30762479969453500 | 3.250713209705390 | 3.234706703868100 | 3.2668729754166600 | 1 in 3 |
| AoU_PLP_plus_LoF | All of Us | 2 – AR+XL | AFR | 247 | 238 | 0.23096776299175500 | 4.329608543836910 | 4.261529798296840 | 4.3916582563692700 | 1 in 4 |
| AoU_PLP_plus_LoF | All of Us | 2 – AR+XL | AMR | 247 | 241 | 0.26167101194825500 | 3.821592588932800 | 3.7660396076433700 | 3.8712245991685400 | 1 in 4 |
| AoU_PLP_plus_LoF | All of Us | 2 – AR+XL | EAS | 247 | 196 | 0.23413803527269900 | 4.270984843770930 | 4.049892247336080 | 4.430313790411850 | 1 in 4 |
| AoU_PLP_plus_LoF | All of Us | 2 – AR+XL | EUR | 247 | 243 | 0.34851397268023900 | 2.869325417025680 | 2.8526271266036300 | 2.8860859914085800 | 1 in 3 |
| AoU_PLP_plus_LoF | All of Us | 2 – AR+XL | MID | 247 | 82 | 0.29094840691984500 | 3.4370354888229200 | 2.8293278413491200 | 3.6200473673946500 | 1 in 3 |
| AoU_PLP_plus_LoF | All of Us | 2 – AR+XL | OTH | 247 | 230 | 0.3296834666197490 | 3.0332124636185700 | 2.97391231833557 | 3.087684066203920 | 1 in 3 |
| AoU_PLP_plus_LoF | All of Us | 2 – AR+XL | SAS | 247 | 171 | 0.2343140537540660 | 4.2677764477993800 | 3.854739851387890 | 4.440880766060110 | 1 in 4 |
| AoU_PLP_plus_LoF | All of Us | 3 – 46 AD | ALL | 46 | 40 | 0.019699034103507400 | 50.76391028847210 | 49.56176358837020 | 52.05048609346180 | 1 in 51 |

| | | | | | | | | | | |
|---|---|---|---|---|---|---|---|---|---|---|
| AoU_PLP_plus_LoF | All of Us | 3 – 46 AD | AFR | 46 | 34 | 0.026824667121297600 | 37.279120575033900 | 35.543794383985300 | 39.10528297709270 | 1 in 37 |
| AoU_PLP_plus_LoF | All of Us | 3 – 46 AD | AMR | 46 | 33 | 0.013381814106314800 | 74.72828362845870 | 69.2869370233577 | 80.60087514617450 | 1 in 75 |
| AoU_PLP_plus_LoF | All of Us | 3 – 46 AD | EAS | 46 | 20 | 0.013257495591829900 | 75.42902753188660 | 60.51017063175580 | 93.43066623779820 | 1 in 75 |
| AoU_PLP_plus_LoF | All of Us | 3 – 46 AD | EUR | 46 | 37 | 0.020014207701248200 | 49.964505961314400 | 48.39715047271430 | 51.63986446148710 | 1 in 50 |
| AoU_PLP_plus_LoF | All of Us | 3 – 46 AD | MID | 46 | 4 | 0.00753763176091371 | 132.66766429019300 | 55.26919092926590 | 544.7460072478030 | 1 in 133 |
| AoU_PLP_plus_LoF | All of Us | 3 – 46 AD | OTH | 46 | 29 | 0.016426007148473100 | 60.879067624961900 | 54.256459671201500 | 68.1405942211284 | 1 in 61 |
| AoU_PLP_plus_LoF | All of Us | 3 – 46 AD | SAS | 46 | 18 | 0.010124322524033200 | 98.77204105521080 | 62.22533231826230 | 138.0853979791650 | 1 in 99 |
| AoU_PLP_plus_LoF | All of Us | 4 – 289 genes | ALL | 289 | 283 | 0.3210501201482620 | 3.114778463680990 | 3.1000436405557100 | 3.1291983526009500 | 1 in 3 |
| AoU_PLP_plus_LoF | All of Us | 4 – 289 genes | AFR | 289 | 269 | 0.2511221716661800 | 3.98212548643181 | 3.9241485915673600 | 4.034890540356380 | 1 in 4 |
| AoU_PLP_plus_LoF | All of Us | 4 – 289 genes | AMR | 289 | 272 | 0.2714274076726930 | 3.684226322515940 | 3.633647721566830 | 3.730976533166380 | 1 in 4 |
| AoU_PLP_plus_LoF | All of Us | 4 – 289 genes | EAS | 289 | 215 | 0.2430895998375320 | 4.113709515620350 | 3.902604478944260 | 4.251888983214360 | 1 in 4 |
| AoU_PLP_plus_LoF | All of Us | 4 – 289 genes | EUR | 289 | 277 | 0.36142502462259200 | 2.766825570653890 | 2.750843641511420 | 2.7828307846760500 | 1 in 3 |
| AoU_PLP_plus_LoF | All of Us | 4 – 289 genes | MID | 289 | 86 | 0.29629297672797300 | 3.375037812381570 | 2.779297412877640 | 3.547418344653210 | 1 in 3 |
| AoU_PLP_plus_LoF | All of Us | 4 – 289 genes | OTH | 289 | 256 | 0.3404854031581120 | 2.9369834674986800 | 2.8806940916713900 | 2.9875621709056800 | 1 in 3 |
| AoU_PLP_plus_LoF | All of Us | 4 – 289 genes | SAS | 289 | 189 | 0.2420661052259800 | 4.131102944241020 | 3.7231896307910400 | 4.2743281065265500 | 1 in 4 |
| AoU_PLP_plus_LoF | All of Us | 5 – 42 AD | ALL | 42 | 37 | 0.0193902387719892 | 51.57234068951140 | 50.319385422779900 | 52.849849707987600 | 1 in 52 |
| AoU_PLP_plus_LoF | All of Us | 5 – 42 AD | AFR | 42 | 31 | 0.026207495218707200 | 38.15702308270140 | 36.336942010720100 | 40.08144629771900 | 1 in 38 |
| AoU_PLP_plus_LoF | All of Us | 5 – 42 AD | AMR | 42 | 31 | 0.01321415775666800 | 75.67640847146610 | 70.26169416663790 | 81.55227373138830 | 1 in 76 |
| AoU_PLP_plus_LoF | All of Us | 5 – 42 AD | EAS | 42 | 19 | 0.011688221869094800 | 85.5562130150979 | 66.97161702712760 | 107.06955175664200 | 1 in 86 |
| AoU_PLP_plus_LoF | All of Us | 5 – 42 AD | EUR | 42 | 34 | 0.019817849348924800 | 50.459562104515300 | 48.88577986326800 | 52.13380884798750 | 1 in 50 |

| | | | | | | | | | | |
|---|---|---|---|---|---|---|---|---|---|---|
| AoU_PLP_plus_LoF | All of Us | 5 – 42 AD | MID | 42 | 4 | 0.00753763176091371 | 132.66766429019300 | 55.530778202079200 | 548.960965571141 | 1 in 133 |
| AoU_PLP_plus_LoF | All of Us | 5 – 42 AD | OTH | 42 | 26 | 0.01611468015549630 | 62.05521861747430 | 55.315503107885400 | 69.54064576959490 | 1 in 62 |
| AoU_PLP_plus_LoF | All of Us | 5 – 42 AD | SAS | 42 | 18 | 0.010124322524033200 | 98.77204105521080 | 61.7368567983407 | 137.79236157592900 | 1 in 99 |
| AoU_PLP_plus_5pctVUS | All of Us | 1 – 293 genes | ALL | 293 | 282 | 0.2786487609713480 | 3.588747340968180 | 3.5723897321085100 | 3.604899401084020 | 1 in 4 |
| AoU_PLP_plus_5pctVUS | All of Us | 1 – 293 genes | AFR | 293 | 265 | 0.18005304533307000 | 5.553918836252710 | 5.466011997561680 | 5.625386480872280 | 1 in 6 |
| AoU_PLP_plus_5pctVUS | All of Us | 1 – 293 genes | AMR | 293 | 273 | 0.23318294778272300 | 4.288478250698630 | 4.229441630545510 | 4.339833910521930 | 1 in 4 |
| AoU_PLP_plus_5pctVUS | All of Us | 1 – 293 genes | EAS | 293 | 226 | 0.1879094920070680 | 5.321710943491800 | 5.053880093496510 | 5.481457079706520 | 1 in 5 |
| AoU_PLP_plus_5pctVUS | All of Us | 1 – 293 genes | EUR | 293 | 281 | 0.3267347344298220 | 3.0605867531809200 | 3.043572966627060 | 3.077127305335030 | 1 in 3 |
| AoU_PLP_plus_5pctVUS | All of Us | 1 – 293 genes | MID | 293 | 127 | 0.2693105775554660 | 3.7131850114355300 | 2.8138453489390300 | 3.7595780047621400 | 1 in 4 |
| AoU_PLP_plus_5pctVUS | All of Us | 1 – 293 genes | OTH | 293 | 258 | 0.3039316509769230 | 3.2902134305055600 | 3.224951409559840 | 3.340123275643900 | 1 in 3 |
| AoU_PLP_plus_5pctVUS | All of Us | 1 – 293 genes | SAS | 293 | 198 | 0.17598985748738800 | 5.682145632009870 | 5.121444782105130 | 5.869128388315230 | 1 in 6 |
| AoU_PLP_plus_5pctVUS | All of Us | 2 – AR+XL | ALL | 247 | 236 | 0.2727943953610460 | 3.665764462193180 | 3.6485291378339500 | 3.682583816539900 | 1 in 4 |
| AoU_PLP_plus_5pctVUS | All of Us | 2 – AR+XL | AFR | 247 | 223 | 0.17315728236038100 | 5.775096411589350 | 5.679720050997260 | 5.853664914979270 | 1 in 6 |
| AoU_PLP_plus_5pctVUS | All of Us | 2 – AR+XL | AMR | 247 | 228 | 0.22826518379170400 | 4.380869580673840 | 4.320641503836980 | 4.4332976062774000 | 1 in 4 |
| AoU_PLP_plus_5pctVUS | All of Us | 2 – AR+XL | EAS | 247 | 192 | 0.17899020275537600 | 5.586897967631740 | 5.302867580453790 | 5.769307013211430 | 1 in 6 |
| AoU_PLP_plus_5pctVUS | All of Us | 2 – AR+XL | EUR | 247 | 235 | 0.32103789014522000 | 3.1148971217934800 | 3.0974905429011200 | 3.131988823763510 | 1 in 3 |
| AoU_PLP_plus_5pctVUS | All of Us | 2 – AR+XL | MID | 247 | 110 | 0.26582874143220800 | 3.7618204661102100 | 2.870647620245820 | 3.856729212988120 | 1 in 4 |
| AoU_PLP_plus_5pctVUS | All of Us | 2 – AR+XL | OTH | 247 | 221 | 0.2983641648903900 | 3.3516089318815100 | 3.2844204650174700 | 3.4038448049858800 | 1 in 3 |
| AoU_PLP_plus_5pctVUS | All of Us | 2 – AR+XL | SAS | 247 | 170 | 0.16922097693910300 | 5.909432849804830 | 5.3292088617425400 | 6.130848563265410 | 1 in 6 |
| AoU_PLP_plus_5pctVUS | All of Us | 3 – 46 AD | ALL | 46 | 46 | 0.008050495723571190 | 124.21595319553800 | 120.27524141812600 | 128.48810624191600 | 1 in 124 |

| | | | | | | | | | | |
|---|---|---|---|---|---|---|---|---|---|---|
| AoU_PLP_plus_5pctVUS | All of Us | 3 – 46 AD | AFR | 46 | 42 | 0.008339872657249450 | 119.90590757171200 | 111.2898490363420 | 129.3364898048600 | 1 in 120 |
| AoU_PLP_plus_5pctVUS | All of Us | 3 – 46 AD | AMR | 46 | 45 | 0.006372349526978520 | 156.92798955335300 | 142.98564650175800 | 171.56926345615600 | 1 in 157 |
| AoU_PLP_plus_5pctVUS | All of Us | 3 – 46 AD | EAS | 46 | 34 | 0.010863803673996300 | 92.04879156584990 | 75.1803696826323 | 110.23702730225600 | 1 in 92 |
| AoU_PLP_plus_5pctVUS | All of Us | 3 – 46 AD | EUR | 46 | 46 | 0.008390518707767410 | 119.18214294359000 | 114.03023453457400 | 124.72891738172000 | 1 in 119 |
| AoU_PLP_plus_5pctVUS | All of Us | 3 – 46 AD | MID | 46 | 17 | 0.004742539404292030 | 210.85749948540100 | 60.560824023058200 | 254.0649555804250 | 1 in 211 |
| AoU_PLP_plus_5pctVUS | All of Us | 3 – 46 AD | OTH | 46 | 37 | 0.007935008173667100 | 126.0238147351340 | 109.39106193761500 | 144.64582574027900 | 1 in 126 |
| AoU_PLP_plus_5pctVUS | All of Us | 3 – 46 AD | SAS | 46 | 28 | 0.008147630549633740 | 122.73506928280500 | 81.54294188705820 | 160.93582978350600 | 1 in 123 |
| AoU_PLP_plus_5pctVUS | All of Us | 4 – 289 genes | ALL | 289 | 278 | 0.27856130686602800 | 3.5898740254006000 | 3.57349325815565 | 3.606108982734870 | 1 in 4 |
| AoU_PLP_plus_5pctVUS | All of Us | 4 – 289 genes | AFR | 289 | 262 | 0.1799543493445960 | 5.556964883827790 | 5.468535267197450 | 5.629839226377370 | 1 in 6 |
| AoU_PLP_plus_5pctVUS | All of Us | 4 – 289 genes | AMR | 289 | 269 | 0.2331039095429430 | 4.289932339447870 | 4.23039013117131 | 4.34029339228564 | 1 in 4 |
| AoU_PLP_plus_5pctVUS | All of Us | 4 – 289 genes | EAS | 289 | 224 | 0.18777599841995700 | 5.325494250673730 | 5.054246093259210 | 5.486800081131760 | 1 in 5 |
| AoU_PLP_plus_5pctVUS | All of Us | 4 – 289 genes | EUR | 289 | 277 | 0.32665026493588700 | 3.061378199697080 | 3.044020551563710 | 3.0781432317019200 | 1 in 3 |
| AoU_PLP_plus_5pctVUS | All of Us | 4 – 289 genes | MID | 289 | 125 | 0.26913022935611800 | 3.7156732723501800 | 2.8175607847813500 | 3.776708551721460 | 1 in 4 |
| AoU_PLP_plus_5pctVUS | All of Us | 4 – 289 genes | OTH | 289 | 254 | 0.3038496970745080 | 3.291100862130490 | 3.2292373640417500 | 3.3411293430817600 | 1 in 3 |
| AoU_PLP_plus_5pctVUS | All of Us | 4 – 289 genes | SAS | 289 | 196 | 0.1758879329542760 | 5.685438353863420 | 5.123308906373870 | 5.87127638492018 | 1 in 6 |
| AoU_PLP_plus_5pctVUS | All of Us | 5 – 42 AD | ALL | 42 | 42 | 0.007930235229479440 | 126.09966426754300 | 122.02743032929700 | 130.63241523809800 | 1 in 126 |
| AoU_PLP_plus_5pctVUS | All of Us | 5 – 42 AD | AFR | 42 | 39 | 0.00822050776914307 | 121.6469867899950 | 112.80606920835000 | 131.39912777037500 | 1 in 122 |
| AoU_PLP_plus_5pctVUS | All of Us | 5 – 42 AD | AMR | 42 | 41 | 0.006269933207124100 | 159.49133219852600 | 145.43899448422500 | 174.658133254915 | 1 in 159 |
| AoU_PLP_plus_5pctVUS | All of Us | 5 – 42 AD | EAS | 42 | 32 | 0.010701206848039700 | 93.44740403585310 | 76.35301219679100 | 112.33533725181100 | 1 in 93 |
| AoU_PLP_plus_5pctVUS | All of Us | 5 – 42 AD | EUR | 42 | 42 | 0.008266108976046180 | 120.97590328143900 | 115.7087054937190 | 126.70124271099100 | 1 in 121 |

| | | | | | | | | | | |
|---|---|---|---|---|---|---|---|---|---|---|
| AoU_PLP_plus_5pctVUS | All of Us | 5 – 42 AD | MID | 42 | 15 | 0.0044968907259452800 | 222.37587278481000 | 64.87011189631180 | 330.0600497762100 | 1 in 222 |
| AoU_PLP_plus_5pctVUS | All of Us | 5 – 42 AD | OTH | 42 | 33 | 0.007818204130439430 | 127.90661171234900 | 111.22762227202100 | 147.3558163078670 | 1 in 128 |
| AoU_PLP_plus_5pctVUS | All of Us | 5 – 42 AD | SAS | 42 | 26 | 0.00802494505772422 | 124.61144503880100 | 83.41625291850600 | 166.66738043226600 | 1 in 125 |
| gnomAD_PLP_only | gnomAD | 1 – 293 genes | TOTAL | 293 | 245 | 0.3122311933787240 | 3.202754949557660 | 3.193102893558720 | 3.2122677146008300 | 1 in 3 |
| gnomAD_PLP_only | gnomAD | 1 – 293 genes | AFR | 293 | 191 | 0.1968764449168940 | 5.079327800855630 | 4.976919506542530 | 5.176654357668440 | 1 in 5 |
| gnomAD_PLP_only | gnomAD | 1 – 293 genes | AMR | 293 | 189 | 0.27297828528193800 | 3.663295045491180 | 3.5995724134533400 | 3.7255398188489600 | 1 in 4 |
| gnomAD_PLP_only | gnomAD | 1 – 293 genes | ASJ | 293 | 125 | 0.5129022361536970 | 1.9496892965394300 | 1.9241724079164500 | 1.9745795013621100 | 1 in 2 |
| gnomAD_PLP_only | gnomAD | 1 – 293 genes | EAS | 293 | 165 | 0.19040896749608600 | 5.251853487523140 | 5.112443198642470 | 5.385633484331520 | 1 in 5 |
| gnomAD_PLP_only | gnomAD | 1 – 293 genes | FIN | 293 | 159 | 0.2611844027839900 | 3.8287125469243300 | 3.759219054422730 | 3.8935206747871900 | 1 in 4 |
| gnomAD_PLP_only | gnomAD | 1 – 293 genes | NFE | 293 | 239 | 0.32993802054552500 | 3.030872278213290 | 3.020812009238710 | 3.0411205215852600 | 1 in 3 |
| gnomAD_PLP_only | gnomAD | 1 – 293 genes | REMAINING | 293 | 194 | 0.3175088988557230 | 3.1495180248614100 | 3.1024658507027400 | 3.195952422494700 | 1 in 3 |
| gnomAD_PLP_only | gnomAD | 2 – AR+XL | TOTAL | 247 | 213 | 0.3035629409050730 | 3.2942097510931300 | 3.284165257564730 | 3.304407939094730 | 1 in 3 |
| gnomAD_PLP_only | gnomAD | 2 – AR+XL | AFR | 247 | 172 | 0.18822632624345400 | 5.312753109289220 | 5.203396626539840 | 5.417632704944610 | 1 in 5 |
| gnomAD_PLP_only | gnomAD | 2 – AR+XL | AMR | 247 | 172 | 0.26345315100225600 | 3.795741277702300 | 3.727891039433900 | 3.8609849141039100 | 1 in 4 |
| gnomAD_PLP_only | gnomAD | 2 – AR+XL | ASJ | 247 | 109 | 0.5089036251821040 | 1.9650085998938700 | 1.9394954067490600 | 1.9899465541987000 | 1 in 2 |
| gnomAD_PLP_only | gnomAD | 2 – AR+XL | EAS | 247 | 148 | 0.17910135249809900 | 5.583430756116790 | 5.426338799135690 | 5.728077979363180 | 1 in 6 |
| gnomAD_PLP_only | gnomAD | 2 – AR+XL | FIN | 247 | 142 | 0.24846755640704100 | 4.024670320988690 | 3.950421223187620 | 4.0964125466329800 | 1 in 4 |
| gnomAD_PLP_only | gnomAD | 2 – AR+XL | NFE | 247 | 209 | 0.32146258869620700 | 3.1107818923994100 | 3.100323187277950 | 3.121332322206200 | 1 in 3 |
| gnomAD_PLP_only | gnomAD | 2 – AR+XL | REMAINING | 247 | 180 | 0.3093455067772540 | 3.2326314043412200 | 3.182808359005450 | 3.281343591465000 | 1 in 3 |
| gnomAD_PLP_only | gnomAD | 3 – 46 AD | TOTAL | 46 | 32 | 0.012446569809072200 | 80.34342114653200 | 78.82472233043120 | 81.94439156707920 | 1 in 80 |

| | | | | | | | | | | |
|---|---|---|---|---|---|---|---|---|---|---|
| gnomAD_PLP_only | gnomAD | 3 – 46 AD | AFR | 46 | 19 | 0.01065582557439950 | 93.84537997717330 | 85.15431044579140 | 103.99095567130500 | 1 in 94 |
| gnomAD_PLP_only | gnomAD | 3 – 46 AD | AMR | 46 | 17 | 0.01293214992724980 | 77.32666305490830 | 69.97946345323500 | 86.0743703605671 | 1 in 77 |
| gnomAD_PLP_only | gnomAD | 3 – 46 AD | ASJ | 46 | 16 | 0.008142212357146740 | 122.81674269061000 | 102.13086744572100 | 148.80912380619400 | 1 in 123 |
| gnomAD_PLP_only | gnomAD | 3 – 46 AD | EAS | 46 | 17 | 0.013774678557941700 | 72.59697536995940 | 65.11475788378770 | 81.51320759433760 | 1 in 73 |
| gnomAD_PLP_only | gnomAD | 3 – 46 AD | FIN | 46 | 17 | 0.016921220747506800 | 59.097391076074800 | 54.205204403796300 | 64.4232991763474 | 1 in 59 |
| gnomAD_PLP_only | gnomAD | 3 – 46 AD | NFE | 46 | 30 | 0.012490736263212000 | 80.05933188624170 | 78.26156156032130 | 81.958749909227 | 1 in 80 |
| gnomAD_PLP_only | gnomAD | 3 – 46 AD | REMAINING | 46 | 14 | 0.011819791456619900 | 84.60386155459010 | 76.77002897731300 | 94.20279572390610 | 1 in 85 |
| gnomAD_PLP_only | gnomAD | 4 – 289 genes | TOTAL | 289 | 243 | 0.3122182561885680 | 3.202887660086220 | 3.193097208878420 | 3.212417585108250 | 1 in 3 |
| gnomAD_PLP_only | gnomAD | 4 – 289 genes | AFR | 289 | 190 | 0.19685486493483700 | 5.079884616166440 | 4.979305319532350 | 5.179260091637130 | 1 in 5 |
| gnomAD_PLP_only | gnomAD | 4 – 289 genes | AMR | 289 | 188 | 0.2729533816568680 | 3.663629275921950 | 3.5986104777459700 | 3.725530750552210 | 1 in 4 |
| gnomAD_PLP_only | gnomAD | 4 – 289 genes | ASJ | 289 | 125 | 0.5129022361536970 | 1.9496892965394300 | 1.9239658958371200 | 1.9745226149203000 | 1 in 2 |
| gnomAD_PLP_only | gnomAD | 4 – 289 genes | EAS | 289 | 165 | 0.19040896749608600 | 5.251853487523140 | 5.110625663974320 | 5.382854516704820 | 1 in 5 |
| gnomAD_PLP_only | gnomAD | 4 – 289 genes | FIN | 289 | 157 | 0.2611370280276000 | 3.82940714135074 | 3.7603385284467300 | 3.894293916205800 | 1 in 4 |
| gnomAD_PLP_only | gnomAD | 4 – 289 genes | NFE | 289 | 237 | 0.3299265634752560 | 3.0309775286554000 | 3.020723720020320 | 3.0410515858516200 | 1 in 3 |
| gnomAD_PLP_only | gnomAD | 4 – 289 genes | REMAINING | 289 | 194 | 0.3175088988557230 | 3.1495180248614100 | 3.100985277847660 | 3.196989066086250 | 1 in 3 |
| gnomAD_PLP_only | gnomAD | 5 – 42 AD | TOTAL | 42 | 30 | 0.012427993557295500 | 80.46351129728240 | 78.8996090730386 | 82.07472368254680 | 1 in 80 |
| gnomAD_PLP_only | gnomAD | 5 – 42 AD | AFR | 42 | 18 | 0.010629241832213500 | 94.0800873463386 | 85.37706116270290 | 104.14821928637900 | 1 in 94 |
| gnomAD_PLP_only | gnomAD | 5 – 42 AD | AMR | 42 | 16 | 0.012898338602004700 | 77.52936489390770 | 70.20139292048700 | 86.22649712258110 | 1 in 78 |
| gnomAD_PLP_only | gnomAD | 5 – 42 AD | ASJ | 42 | 16 | 0.008142212357146740 | 122.81674269061000 | 102.45864133927600 | 148.56841651687900 | 1 in 123 |
| gnomAD_PLP_only | gnomAD | 5 – 42 AD | EAS | 42 | 17 | 0.013774678557941700 | 72.59697536995940 | 64.99771525732650 | 81.57217848852660 | 1 in 73 |

| | | | | | | | | | | |
|---|---|---|---|---|---|---|---|---|---|---|
| gnomAD_PLP_only | gnomAD | 5 – 42 AD | FIN | 42 | 15 | 0.016858183207619200 | 59.318373023021800 | 54.50761305080000 | 64.83881978148000 | 1 in 59 |
| gnomAD_PLP_only | gnomAD | 5 – 42 AD | NFE | 42 | 28 | 0.012473851313201300 | 80.16770241133780 | 78.33392256005300 | 82.05264864786880 | 1 in 80 |
| gnomAD_PLP_only | gnomAD | 5 – 42 AD | REMAINING | 42 | 14 | 0.011819791456619900 | 84.60386155459010 | 76.72681607989 | 94.41086888017370 | 1 in 85 |
| gnomAD_PLP_plus_5pctVUS | gnomAD | 1 – 293 genes | TOTAL | 293 | 285 | 0.3144761767757390 | 3.1798911137015200 | 3.170177079412550 | 3.189437727171900 | 1 in 3 |
| gnomAD_PLP_plus_5pctVUS | gnomAD | 1 – 293 genes | AFR | 293 | 266 | 0.20386655063380700 | 4.905169567499280 | 4.803960831130350 | 4.9939458631841900 | 1 in 5 |
| gnomAD_PLP_plus_5pctVUS | gnomAD | 1 – 293 genes | AMR | 293 | 269 | 0.2803691954550990 | 3.566725646791490 | 3.5034163326508400 | 3.624311220680310 | 1 in 4 |
| gnomAD_PLP_plus_5pctVUS | gnomAD | 1 – 293 genes | ASJ | 293 | 157 | 0.513610441606852 | 1.9470009154632000 | 1.920054594595940 | 1.9706361621179700 | 1 in 2 |
| gnomAD_PLP_plus_5pctVUS | gnomAD | 1 – 293 genes | EAS | 293 | 242 | 0.19577339135732800 | 5.1079464531254200 | 4.960357858616230 | 5.227303146280760 | 1 in 5 |
| gnomAD_PLP_plus_5pctVUS | gnomAD | 1 – 293 genes | FIN | 293 | 197 | 0.2617445122681990 | 3.820519449803560 | 3.747157412980280 | 3.881452918543120 | 1 in 4 |
| gnomAD_PLP_plus_5pctVUS | gnomAD | 1 – 293 genes | NFE | 293 | 285 | 0.3315563310121740 | 3.016078736748000 | 3.0059926675960800 | 3.0262781076381000 | 1 in 3 |
| gnomAD_PLP_plus_5pctVUS | gnomAD | 1 – 293 genes | REMAINING | 293 | 267 | 0.3209532726301900 | 3.115718346801920 | 3.067022486271310 | 3.160603715664830 | 1 in 3 |
| gnomAD_PLP_plus_5pctVUS | gnomAD | 2 – AR+XL | TOTAL | 247 | 239 | 0.3051122376368380 | 3.2774824364477200 | 3.2674035465149300 | 3.2876033253052700 | 1 in 3 |
| gnomAD_PLP_plus_5pctVUS | gnomAD | 2 – AR+XL | AFR | 247 | 221 | 0.19333170537765000 | 5.172457347575860 | 5.06528672190884 | 5.27081367613617 | 1 in 5 |
| gnomAD_PLP_plus_5pctVUS | gnomAD | 2 – AR+XL | AMR | 247 | 226 | 0.2692713175823820 | 3.7137263967747200 | 3.6464196418594300 | 3.776232485381050 | 1 in 4 |
| gnomAD_PLP_plus_5pctVUS | gnomAD | 2 – AR+XL | ASJ | 247 | 128 | 0.5093751704661120 | 1.963189527053180 | 1.935897877761490 | 1.9879833397649000 | 1 in 2 |
| gnomAD_PLP_plus_5pctVUS | gnomAD | 2 – AR+XL | EAS | 247 | 202 | 0.18300083214184100 | 5.464456026215870 | 5.3058004524627700 | 5.59818945491775 | 1 in 5 |
| gnomAD_PLP_plus_5pctVUS | gnomAD | 2 – AR+XL | FIN | 247 | 169 | 0.24881721894349500 | 4.019014456660630 | 3.939326219880860 | 4.087257053460190 | 1 in 4 |
| gnomAD_PLP_plus_5pctVUS | gnomAD | 2 – AR+XL | NFE | 247 | 239 | 0.32251582990504900 | 3.1006229997901400 | 3.090020809900100 | 3.1111590218199200 | 1 in 3 |
| gnomAD_PLP_plus_5pctVUS | gnomAD | 2 – AR+XL | REMAINING | 247 | 226 | 0.311864581582886 | 3.2065199418428500 | 3.154882685214220 | 3.2538476465588300 | 1 in 3 |
| gnomAD_PLP_plus_5pctVUS | gnomAD | 3 – 46 AD | TOTAL | 46 | 46 | 0.013475469919137300 | 74.2089148653616 | 72.87740083044320 | 75.61873855305920 | 1 in 74 |

| | | | | | | | | | | |
|---|---|---|---|---|---|---|---|---|---|---|
| gnomAD_PLP_plus_5pctVUS | gnomAD | 3 – 46 AD | AFR | 46 | 45 | 0.013059699168030300 | 76.57144220044260 | 69.9262610047878 | 83.47579005267250 | 1 in 77 |
| gnomAD_PLP_plus_5pctVUS | gnomAD | 3 – 46 AD | AMR | 46 | 43 | 0.015187412427824300 | 65.84400106024230 | 59.885725161633500 | 72.00384233956520 | 1 in 66 |
| gnomAD_PLP_plus_5pctVUS | gnomAD | 3 – 46 AD | ASJ | 46 | 29 | 0.008632402776606370 | 115.84260209799000 | 95.13815500793030 | 135.5325176352560 | 1 in 116 |
| gnomAD_PLP_plus_5pctVUS | gnomAD | 3 – 46 AD | EAS | 46 | 40 | 0.015633503335103400 | 63.96518928387640 | 57.37462364437740 | 70.74590445213390 | 1 in 64 |
| gnomAD_PLP_plus_5pctVUS | gnomAD | 3 – 46 AD | FIN | 46 | 28 | 0.017209251397538800 | 58.108280069812600 | 53.25435943130970 | 63.108208780868600 | 1 in 58 |
| gnomAD_PLP_plus_5pctVUS | gnomAD | 3 – 46 AD | NFE | 46 | 46 | 0.013344224863376600 | 74.93878514776180 | 73.309350422695 | 76.63320087740390 | 1 in 75 |
| gnomAD_PLP_plus_5pctVUS | gnomAD | 3 – 46 AD | REMAINING | 46 | 41 | 0.013207707093772900 | 75.71336893679840 | 68.29406829754530 | 83.3348675869628 | 1 in 76 |
| gnomAD_PLP_plus_5pctVUS | gnomAD | 4 – 289 genes | TOTAL | 289 | 281 | 0.31441832375979200 | 3.1804762141152300 | 3.1709759862293300 | 3.189952291801600 | 1 in 3 |
| gnomAD_PLP_plus_5pctVUS | gnomAD | 4 – 289 genes | AFR | 289 | 263 | 0.20376918862311300 | 4.90751328381436 | 4.805981570452600 | 4.998709865562040 | 1 in 5 |
| gnomAD_PLP_plus_5pctVUS | gnomAD | 4 – 289 genes | AMR | 289 | 266 | 0.2802218002463140 | 3.5686017259221200 | 3.504806849686400 | 3.6274054263298300 | 1 in 4 |
| gnomAD_PLP_plus_5pctVUS | gnomAD | 4 – 289 genes | ASJ | 289 | 154 | 0.5135739049213160 | 1.947139429043240 | 1.9201118449069100 | 1.970906010259700 | 1 in 2 |
| gnomAD_PLP_plus_5pctVUS | gnomAD | 4 – 289 genes | EAS | 289 | 239 | 0.19570783722769300 | 5.109657406497060 | 4.964741350022310 | 5.2294303931180000 | 1 in 5 |
| gnomAD_PLP_plus_5pctVUS | gnomAD | 4 – 289 genes | FIN | 289 | 194 | 0.2616806074093620 | 3.821452456488840 | 3.7481900493480500 | 3.8833021997169600 | 1 in 4 |
| gnomAD_PLP_plus_5pctVUS | gnomAD | 4 – 289 genes | NFE | 289 | 281 | 0.33150677204816300 | 3.016529628706090 | 3.006255538656990 | 3.026435924993780 | 1 in 3 |
| gnomAD_PLP_plus_5pctVUS | gnomAD | 4 – 289 genes | REMAINING | 289 | 263 | 0.32088874521575300 | 3.116344885600900 | 3.067820814260320 | 3.1598513872010700 | 1 in 3 |
| gnomAD_PLP_plus_5pctVUS | gnomAD | 5 – 42 AD | TOTAL | 42 | 42 | 0.013392214724441600 | 74.67024839251850 | 73.27414829547610 | 76.09820271397830 | 1 in 75 |
| gnomAD_PLP_plus_5pctVUS | gnomAD | 5 – 42 AD | AFR | 42 | 42 | 0.012939002704138900 | 77.28570917448830 | 70.63353406166310 | 84.29558502735400 | 1 in 77 |
| gnomAD_PLP_plus_5pctVUS | gnomAD | 5 – 42 AD | AMR | 42 | 40 | 0.014985702528743000 | 66.73027160935390 | 60.64270827107620 | 72.96882891460440 | 1 in 67 |
| gnomAD_PLP_plus_5pctVUS | gnomAD | 5 – 42 AD | ASJ | 42 | 26 | 0.00855793307320718 | 116.85064506180300 | 96.06380099300530 | 138.06986015616200 | 1 in 117 |
| gnomAD_PLP_plus_5pctVUS | gnomAD | 5 – 42 AD | EAS | 42 | 37 | 0.015553265640606000 | 64.2951791030451 | 57.57990294751140 | 71.33633462418660 | 1 in 64 |

| | | | | | | | | | | |
|---|---|---|---|---|---|---|---|---|---|---|
| gnomAD_PLP_plus_5pctVUS | gnomAD | 5 – 42 AD | FIN | 42 | 25 | 0.017124179081654400 | 58.39695994953280 | 53.51814151944270 | 63.478948877845200 | 1 in 58 |
| gnomAD_PLP_plus_5pctVUS | gnomAD | 5 – 42 AD | NFE | 42 | 42 | 0.013271073391801400 | 75.35185515723060 | 73.70086547834160 | 77.02534032275610 | 1 in 75 |
| gnomAD_PLP_plus_5pctVUS | gnomAD | 5 – 42 AD | REMAINING | 42 | 37 | 0.013113935704144000 | 76.25475849206740 | 68.98778381149490 | 83.915202791947 | 1 in 76 |

#### SUPPLEMENTAL FIGURES

Supplemental Figure 1. Figure showing the top 20 genes by carrier frequency in the gnomAD database.  The inheritance model legend reflects the perinatal-treatable phenotypes used for this analysis, not necessarily all known inheritance mechanisms for the gene. *P/LP, pathogenic/likely pathogenic*.

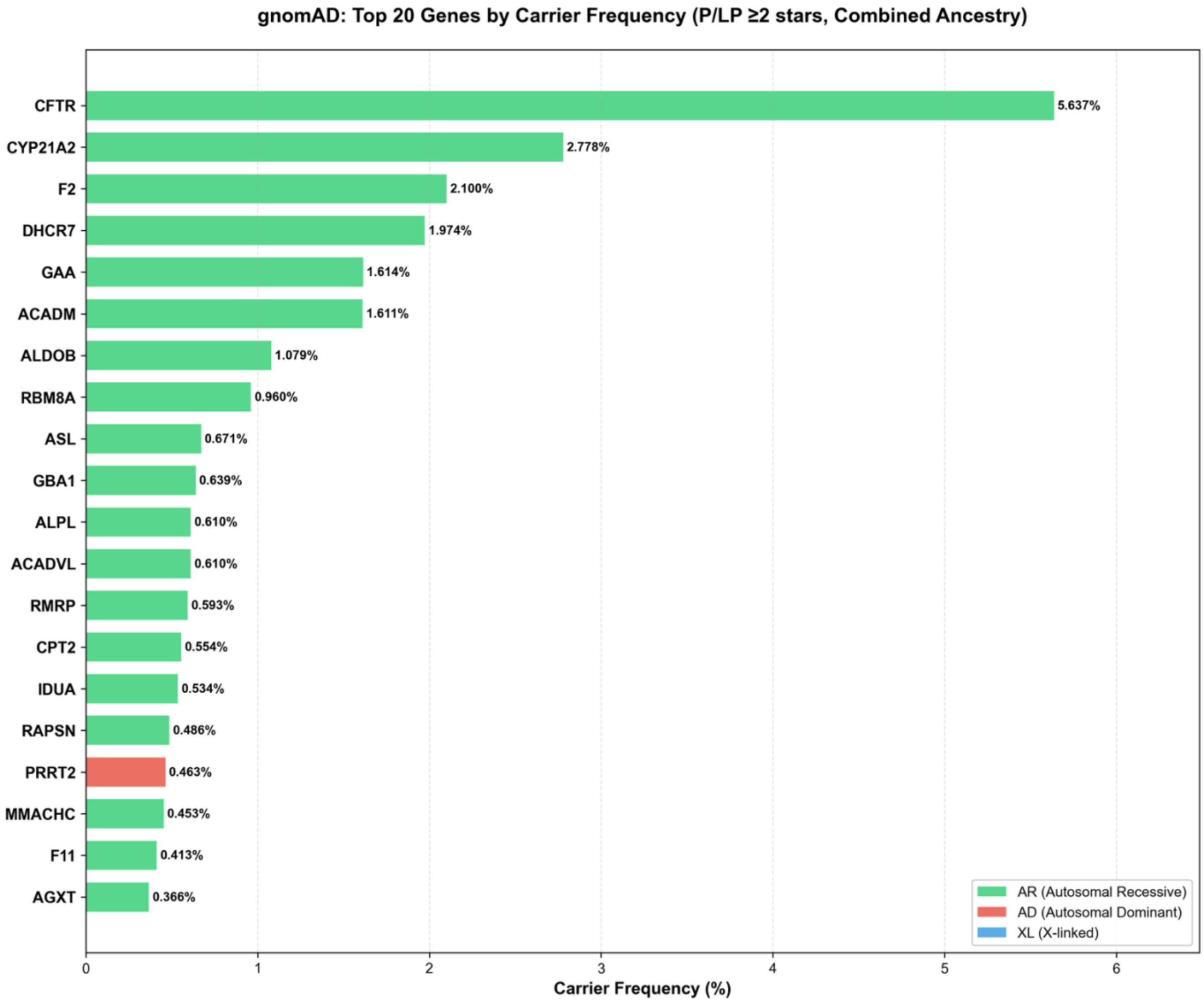

Supplemental Figure 2. Figure showing the top 20 genes by carrier frequency in the All of Us database. The inheritance model legend reflects the perinatal-treatable phenotypes used for this analysis, not necessarily all known inheritance mechanisms for the gene. *P/LP, pathogenic/likely pathogenic*.

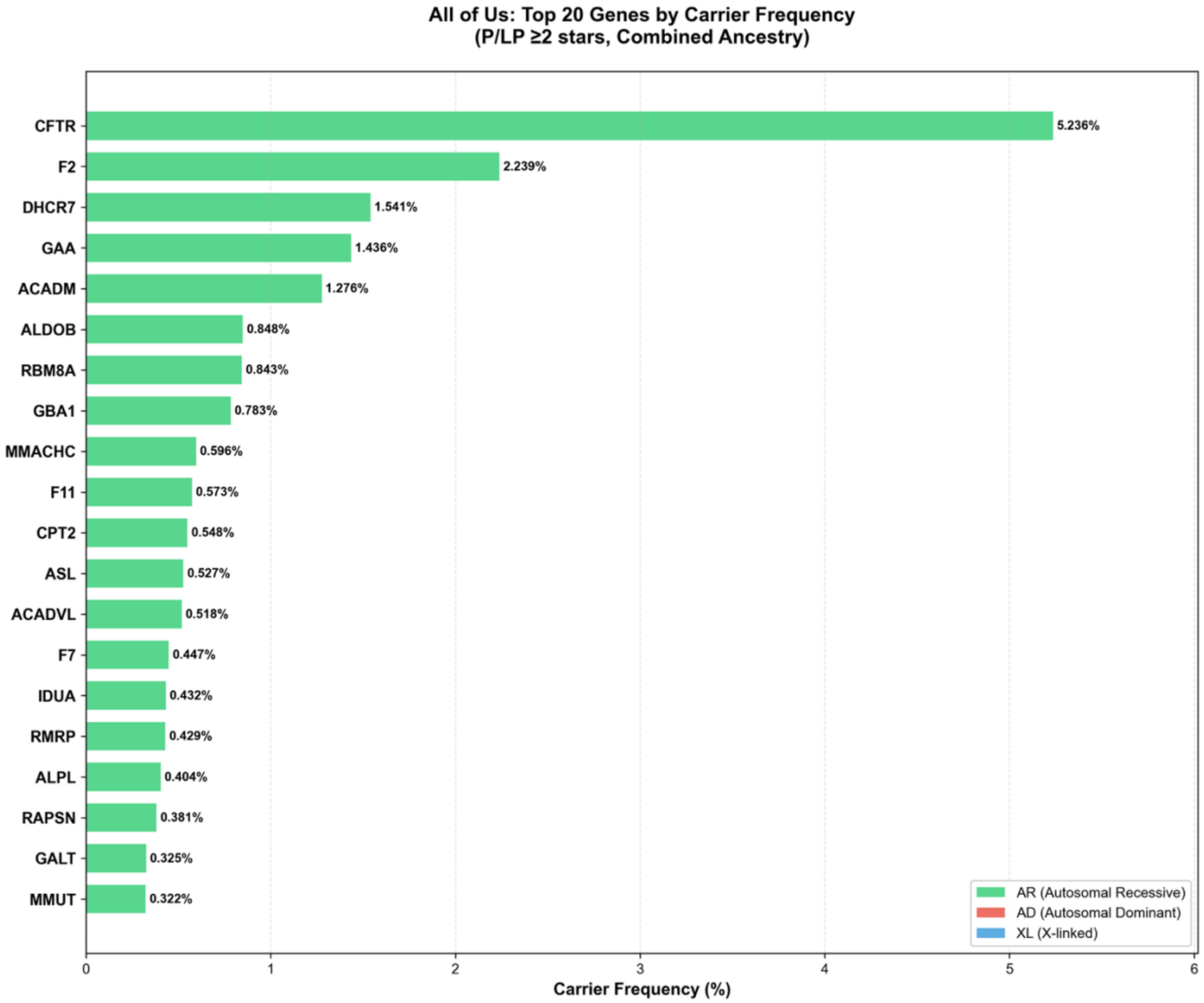

Supplemental Figure 3. Cumulative contribution plot showing the share of total qualifying heterozygote yield explained by the top 1, 3, 5, 10 and 20 genes for each of the two datasets (AoU and gnomAD). *P/LP, pathogenic/likely pathogenic; CF, carrier frequency; AoU, All of Us (v8).*

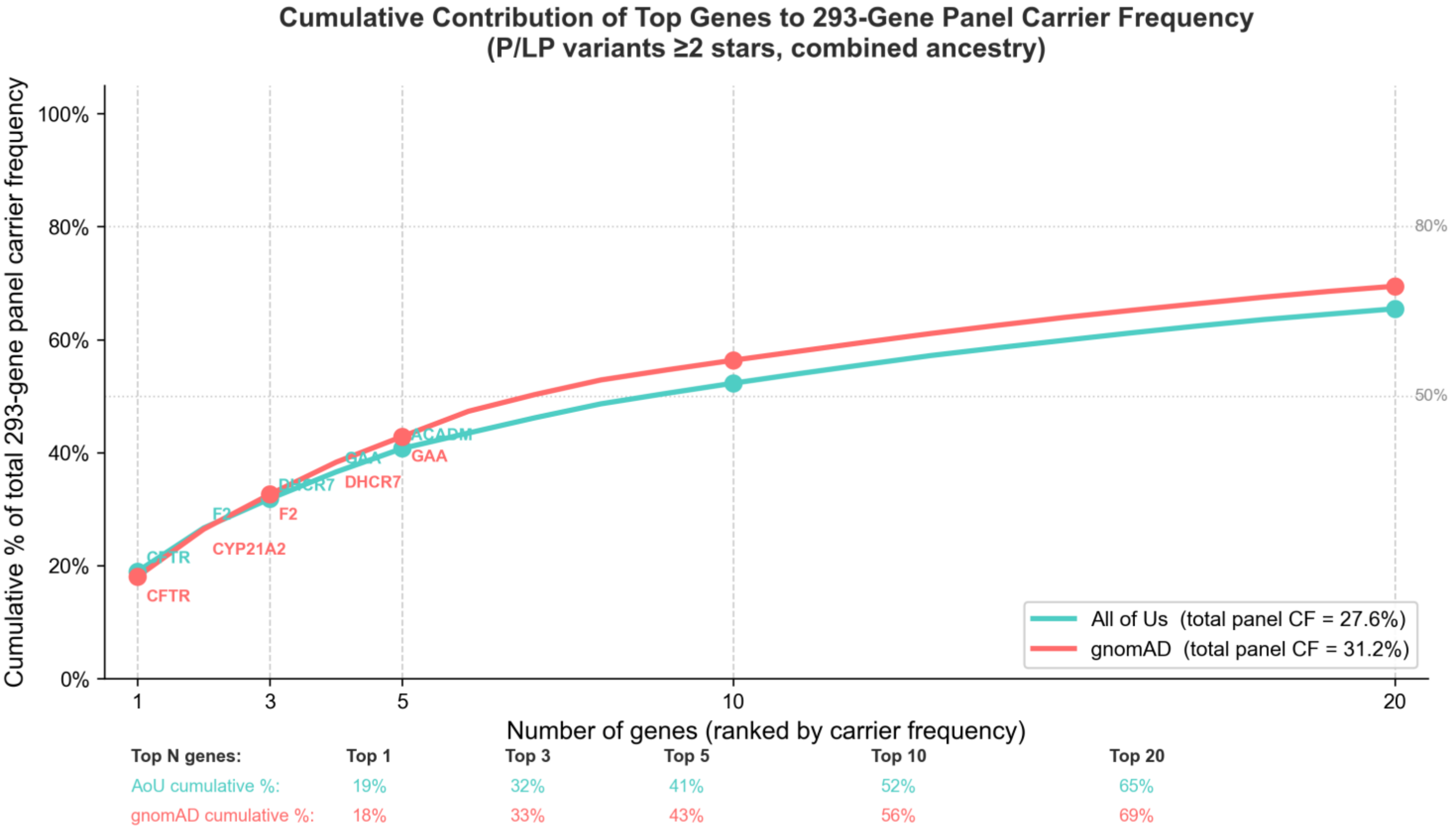

Supplemental Figure 4. NNS for 293-gene panel across ancestral groups and across two variant inclusion datasets. NNS refers to the individual level heterozygote yield (i.e., NNS to ascertain a single individual with a heterozygous pathogenic or likely pathogenic (P/LP) variant in a gene on the list). *P/LP, pathogenic/likely pathogenic; VUS, variant of uncertain significance; NNS, number needed to screen.*

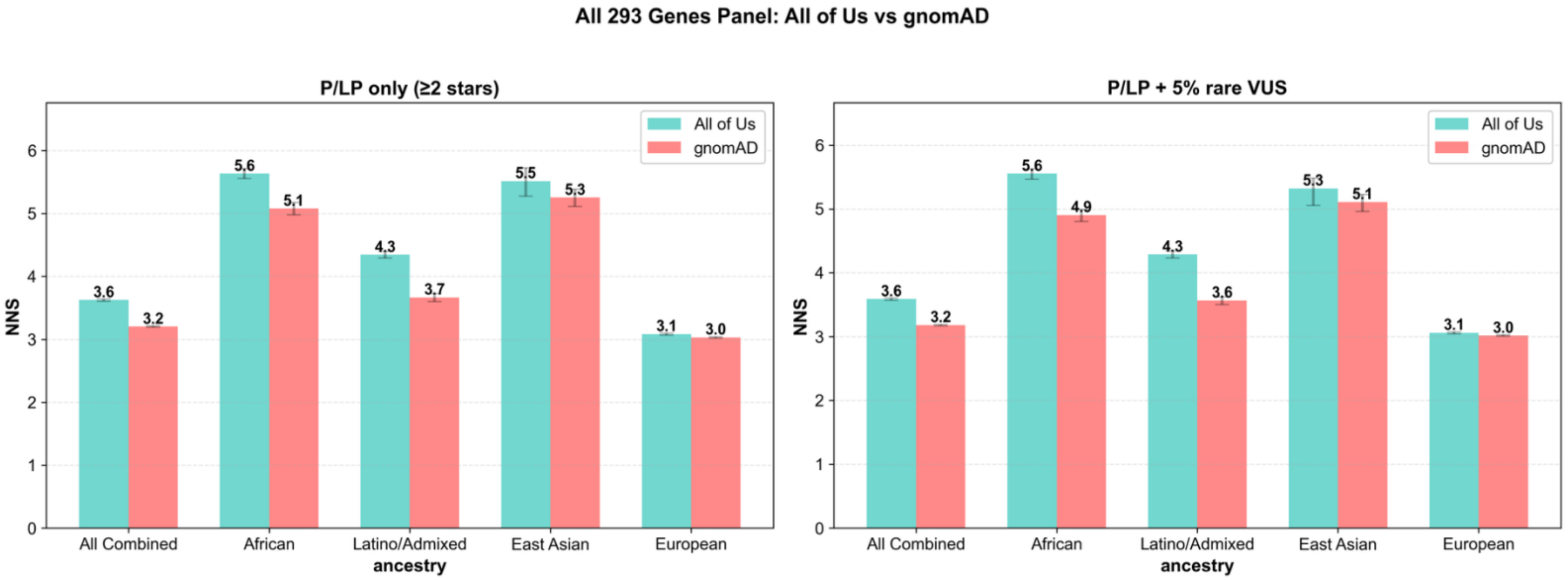

Supplemental Figure 5. Carrier frequency of AD genes compared between All of Us and gnomAD (excluding 4 highly penetrant early onset genes: *FAM111A, SNAP25, SCN8A,* and *KCNQ3*). Estimates of heterozygotes for AD conditions do not imply that all heterozygous individuals are clinically affected. *AD, autosomal dominant; P/LP, pathogenic/likely pathogenic.*

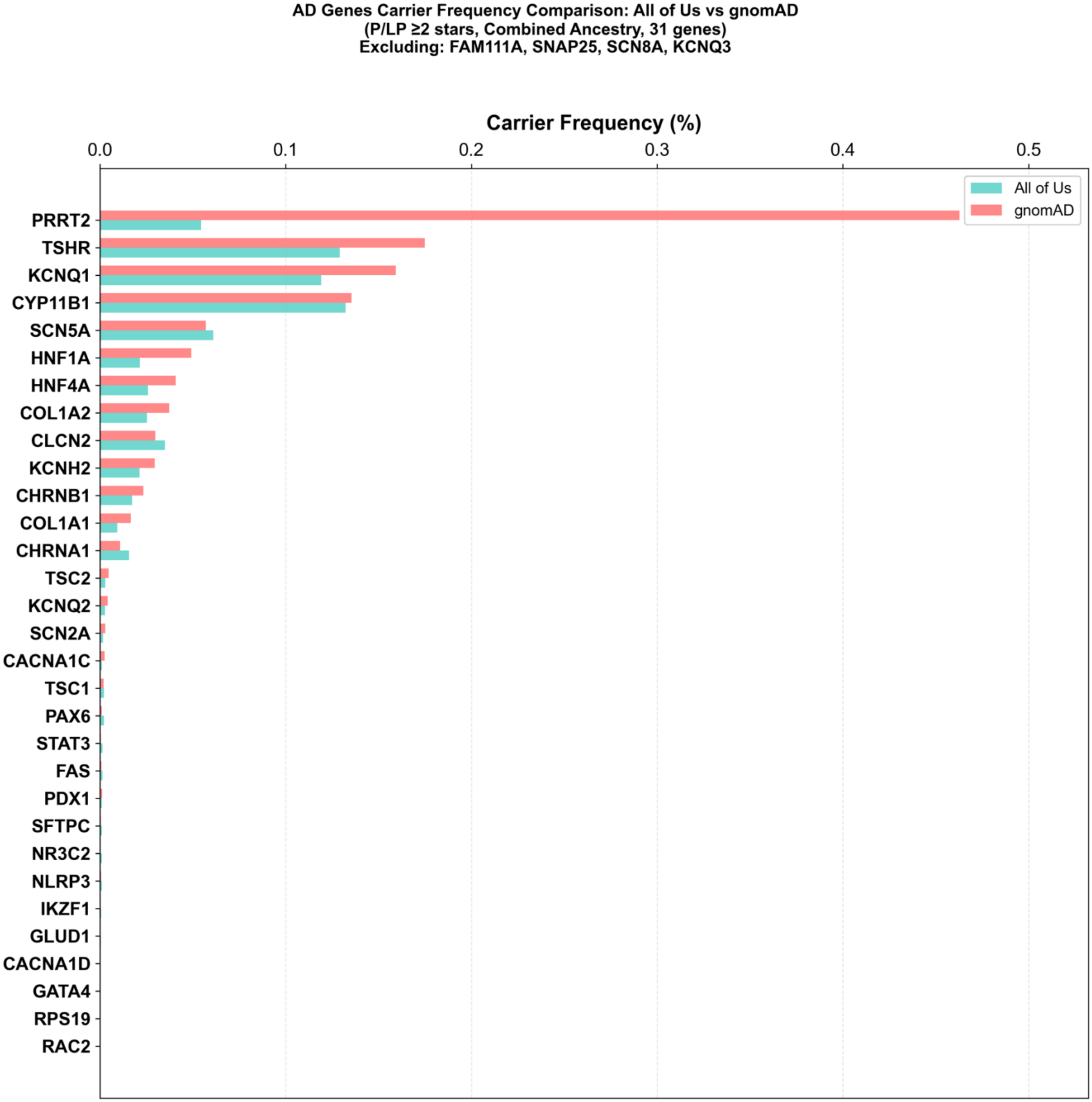
